## Supplemental Figures for "Cost and social distancing dynamics in a mathematical model of COVID-19 with application to Ontario, Canada"

### Supplementary Material: Additional Figures

We plot the active cases (tested and actual cases) along with the cost for each of the 25 scenarios considered in Figure 3 of the main text. These are plotted in Figure 1 for all cases of $M_c^*/4$, Figure 2 for all cases of $M_c^*/2$, Figure 3 for all cases of $M_c^*$, Figure 4 for all cases of $2M_c^*$, and Figure 5 for all cases of $4M_c^*$.

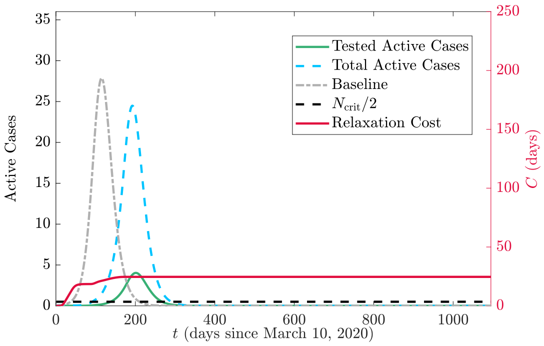


(a) $C_c^*/4$.

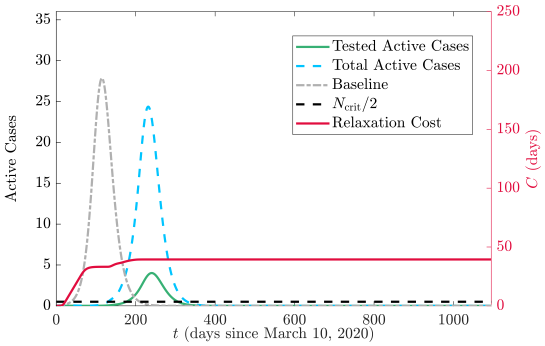


(b) $C_c^*/2$.

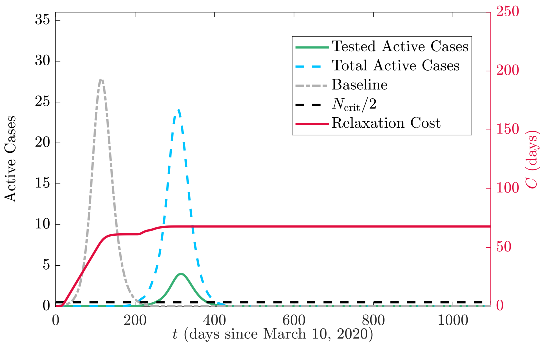


(c) $C_c^*$.

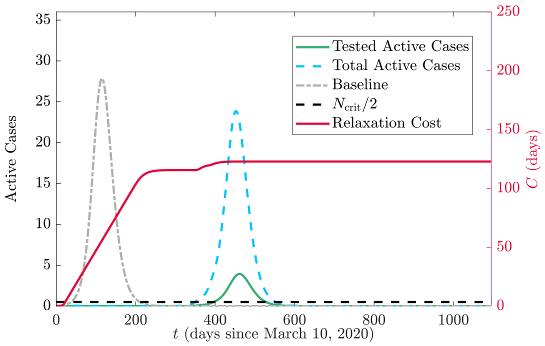


(d) $2C_c^*$.

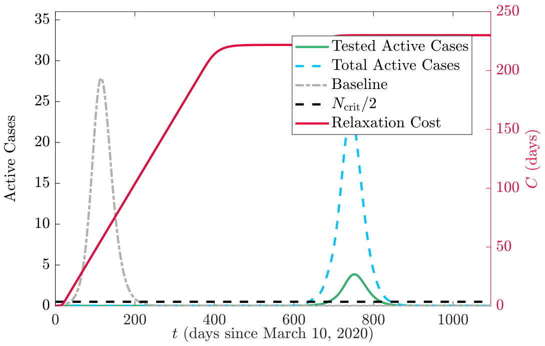


(e) $4C_c^*$.

Figure 1: Tested active cases (green solid), total active cases (blue dashed) and relaxation cost (red solid) for $M_c^*/4$ and varying $C_c$. The grey curve represents the baseline case of no implementation of social distancing (and thus no cost) and the black dashed line is the critical threshold $N_{\text{crit}}/2$. Case values are scaled by $N_{\text{crit}}$.

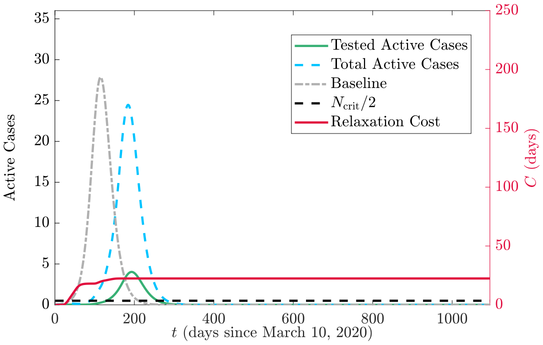


(a) $C_c^*/4$.

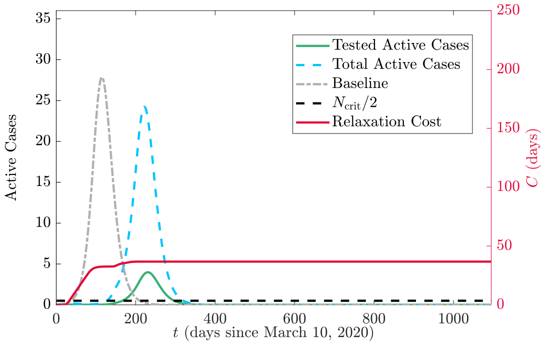


(b) $C_c^*/2$.

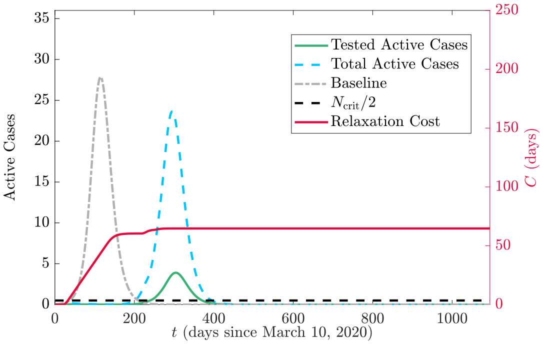


(c) $C_c^*$.

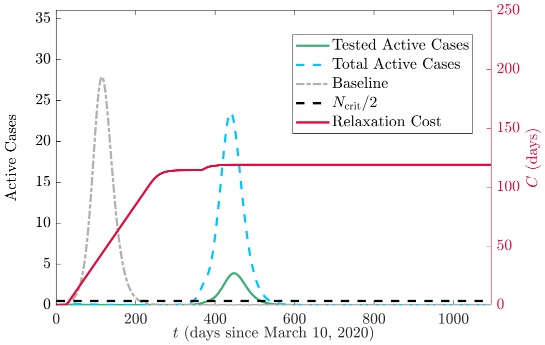


(d) $2C_c^*$.

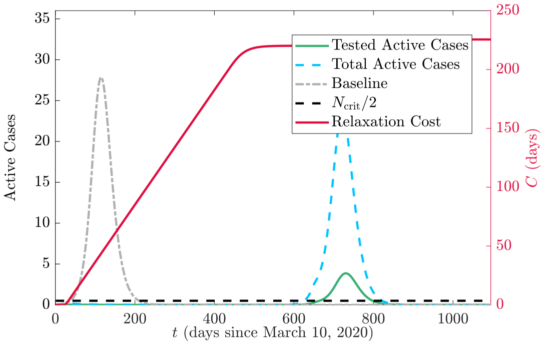


(e) $4C_c^*$.

Figure 2: Tested active cases (green solid), total active cases (blue dashed) and relaxation cost (red solid) for $M_c^*/2$ and varying $C_c$. The grey curve represents the baseline case of no implementation of social distancing (and thus no cost) and the black dashed line is the critical threshold $N_{\rm crit}/2$. Case values are scaled by $N_{\rm crit}$.

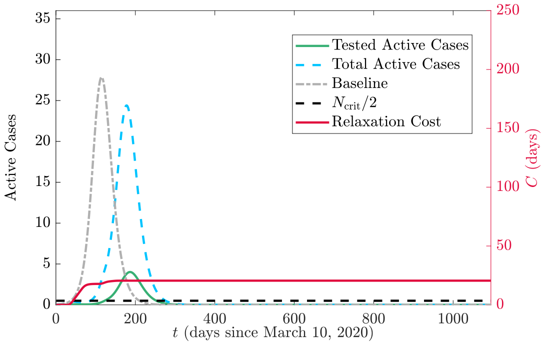


(a) $C_c^*/4$.

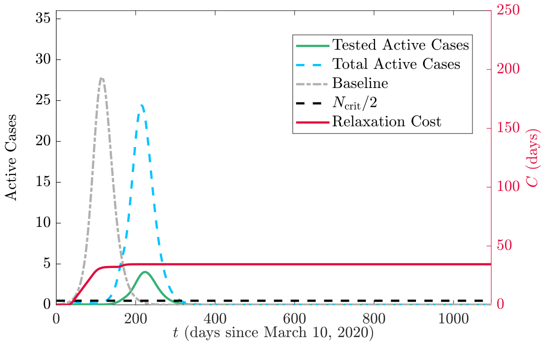


(b) $C_c^*/2$.

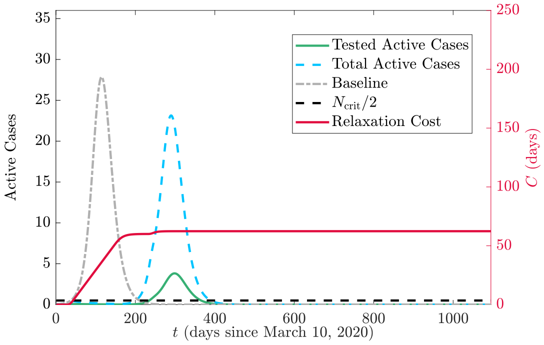


(c) $C_c^*$.

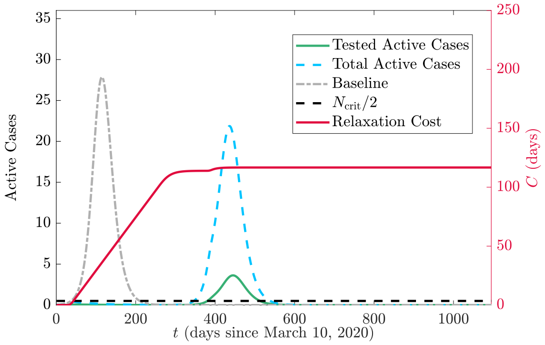


(d) $2C_c^*$.

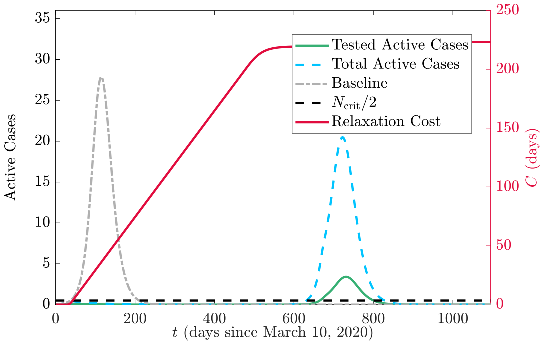


(e) $4C_c^*$.

Figure 3: Tested active cases (green solid), total active cases (blue dashed) and relaxation cost (red solid) for $M_c^*$ and varying $C_c$. The grey curve represents the baseline case of no implementation of social distancing (and thus no cost) and the black dashed line is the critical threshold $N_{\mathrm{crit}}/2$. Case values are scaled by $N_{\mathrm{crit}}$.

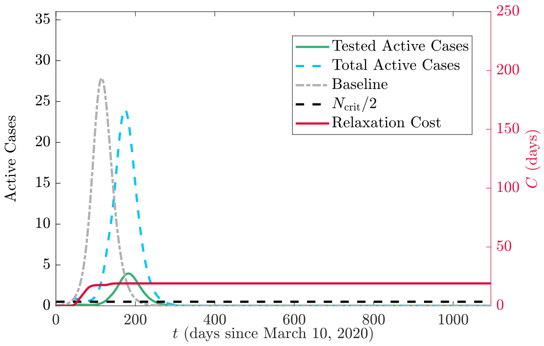


(a) $C_c^*/4$.

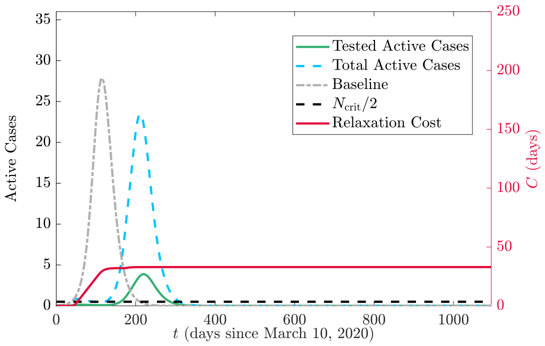


(b) $C_c^*/2$.

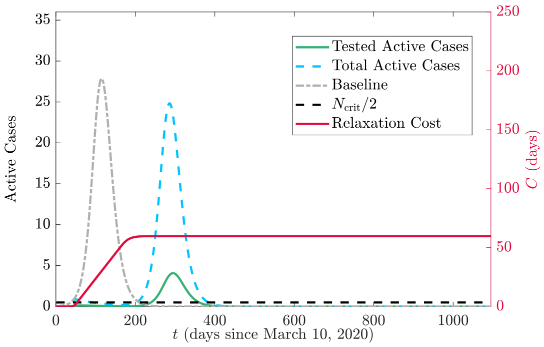


(c) $C_c^*$.

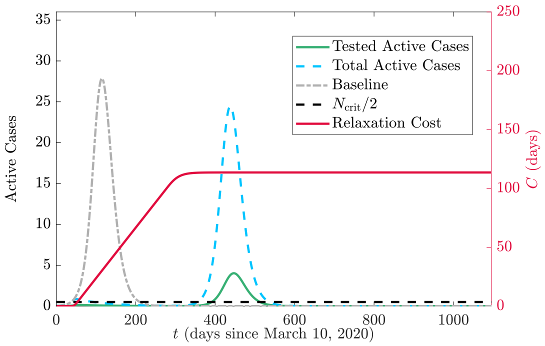


(d) $2C_c^*$.

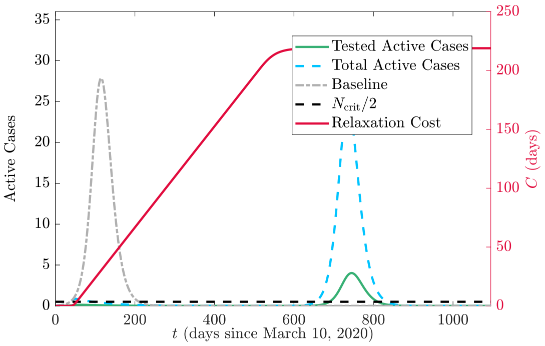


(e) $4C_c^*$.

Figure 4: Tested active cases (green solid), total active cases (blue dashed) and relaxation cost (red solid) for $2M_c^*$ and varying $C_c$. The grey curve represents the baseline case of no implementation of social distancing (and thus no cost) and the black dashed line is the critical threshold $N_{\rm crit}/2$. Case values are scaled by $N_{\rm crit}$.

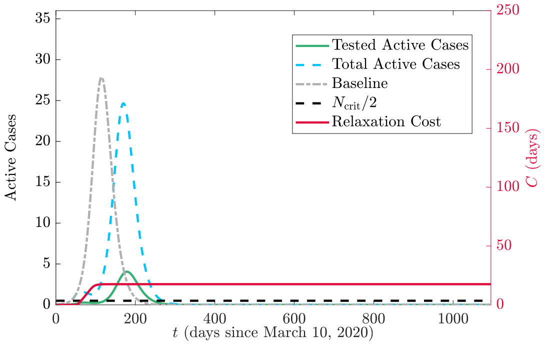


(a) $C_c^*/4$.

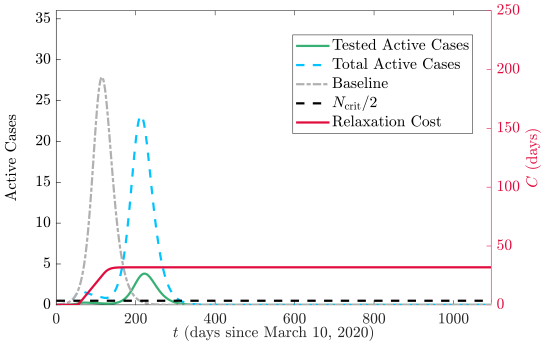


(b) $C_c^*/2$.

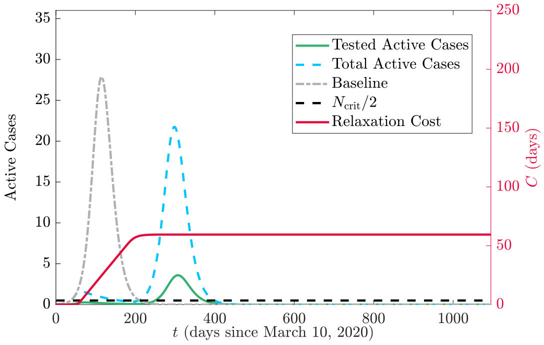


(c) $C_c^*$.

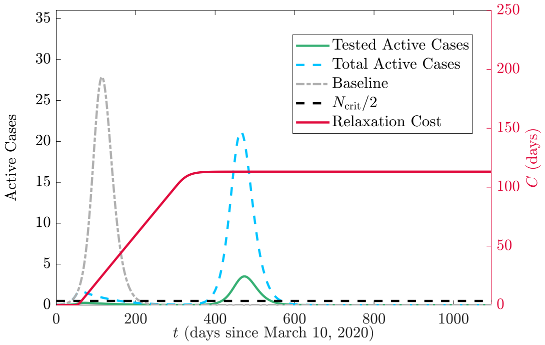


(d) $2C_c^*$.

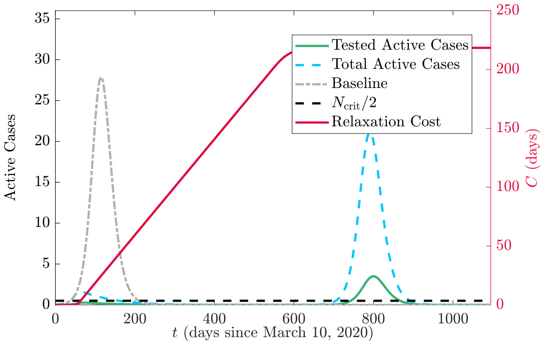


(e) $4C_c^*$.

Figure 5: Tested active cases (green solid), total active cases (blue dashed) and relaxation cost (red solid) for $4M_c^*$ and varying $C_c$. The grey curve represents the baseline case of no implementation of social distancing (and thus no cost) and the black dashed line is the critical threshold $N_{\rm crit}/2$. Case values are scaled by $N_{\rm crit}$.

We also plot the populations in each of the two social distancing classes for each of the 25 scenarios considered in Figure 3 of the main text. These are plotted in Figure 6 for social distancing class 1 and Figure 7 for social distancing class 2.

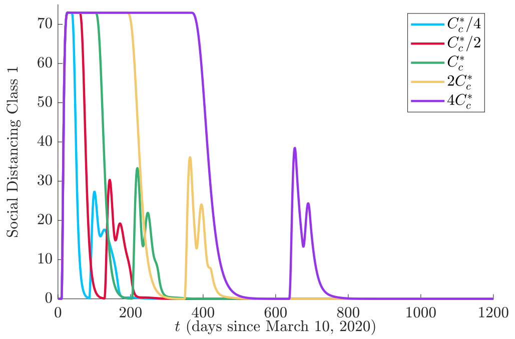


(a) $M_c^*/4$.

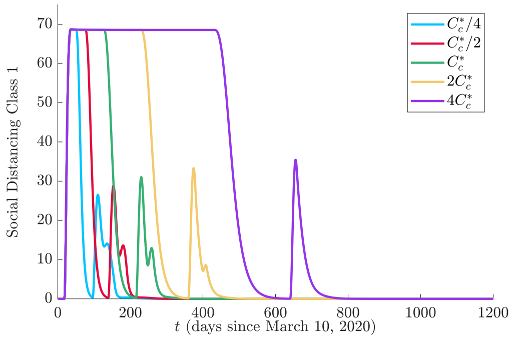


(b) $M_c^*/2$.

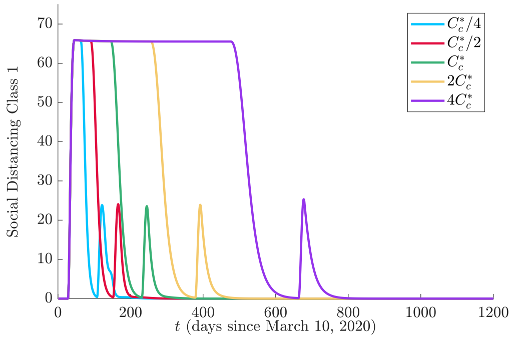


(c) $M_c^*$.

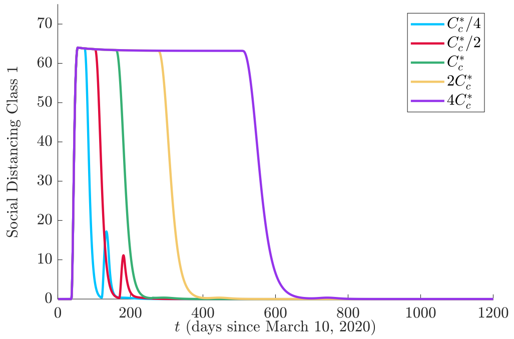


(d) $2M_c^*$.

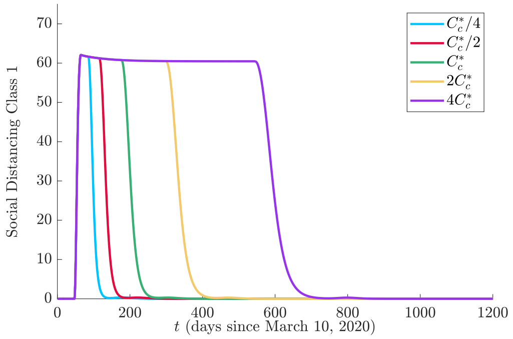


(e) $4M_c^*$.

Figure 6: Total people in social distancing class 1 ($S_1$, $E_1$, $P_1$, $I_{S_1}$, $I_{A_1}$) for different values of $M_c$ and $C_c$.

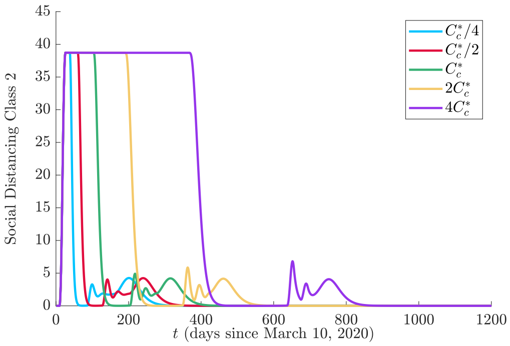


(a) $M_c^*/4$.

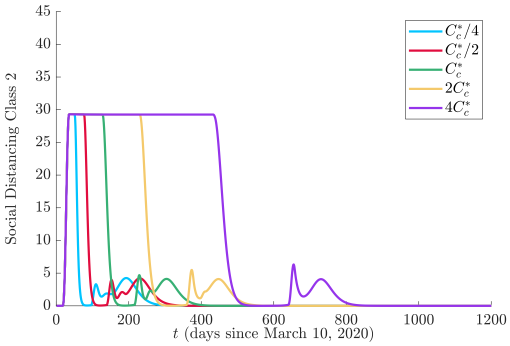


(b) $M_c^*/2$.

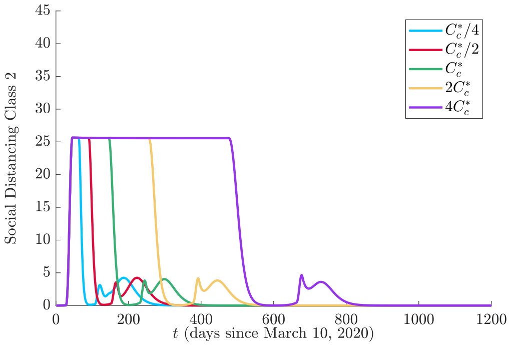


(c) $M_c^*$.

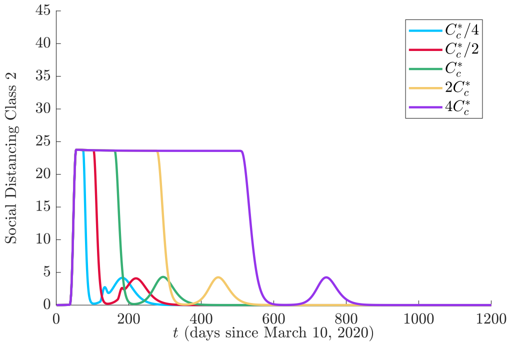


(d) $2M_c^*$.

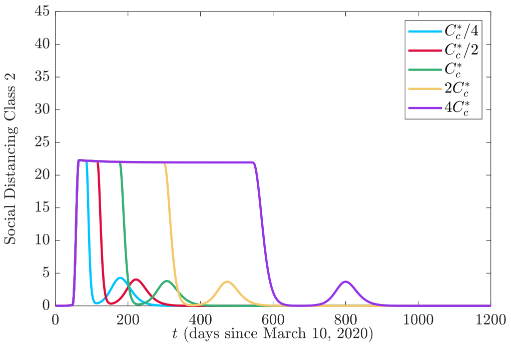


(e) $4M_c^*$.

Figure 7: Total people in social distancing class 2 ($S_2$, $E_2$, $P_2$, $P_M$, $I_{S_2}$, $I_{S_M}$, $I_{A_2}$, $I_{A_M}$) for different values of $M_c$ and $C_c$.

Next, we plot the active cases (tested and actual cases) along with the cost for each of the 25 scenarios considered in Figure 8 using the modified relaxation cost (3.1). These are plotted in Figure 8 for all cases of $\eta = 1/4$, Figure 9 for all cases of $\eta = 1/2$, Figure 10 for all cases of $\eta = 1$, Figure 11 for all cases of $\eta = 2$, and Figure 12 for all cases of $\eta = 4$.

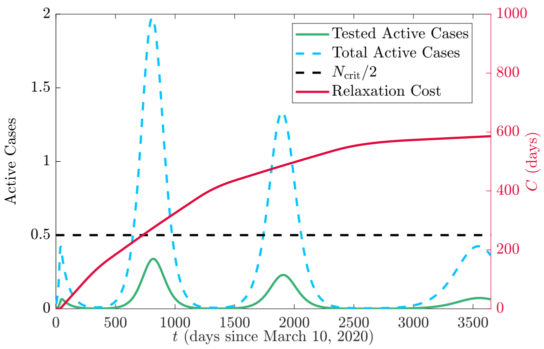


(a) $C_c^*/4$.

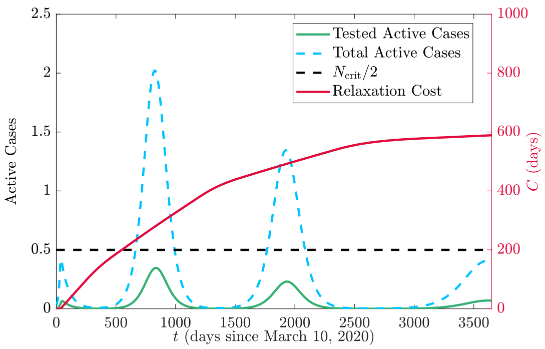


(b) $C_c^*/2$.

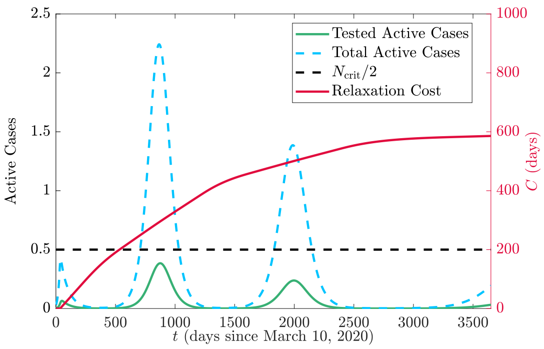


(c) $C_c^*$.

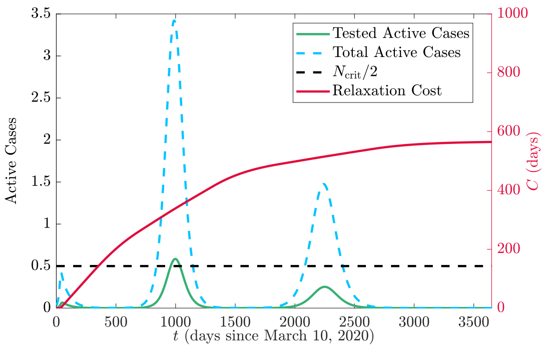


(d) $2C_c^*$.

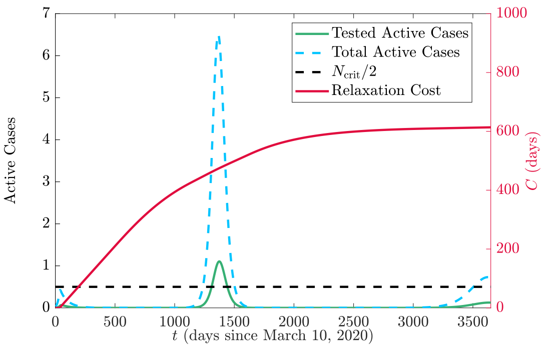


(e) $4C_c^*$.

Figure 8: Tested active cases (green solid), total active cases (blue dashed) and relaxation cost (red solid) for $\eta = 1/4$ and varying $C_c$. The grey curve represents the baseline case of no implementation of social distancing (and thus no cost) and the black dashed line is the critical threshold $N_{\text{crit}}/2$. Case values are scaled by $N_{\text{crit}}$.

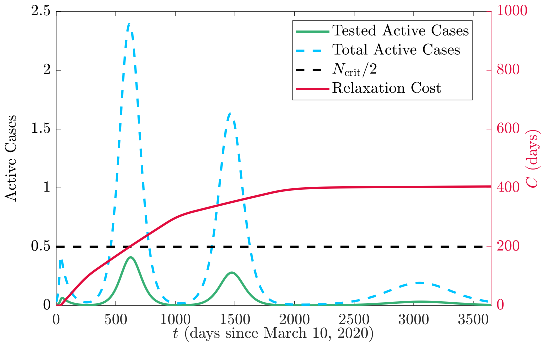


(a) $C_c^*/4$.

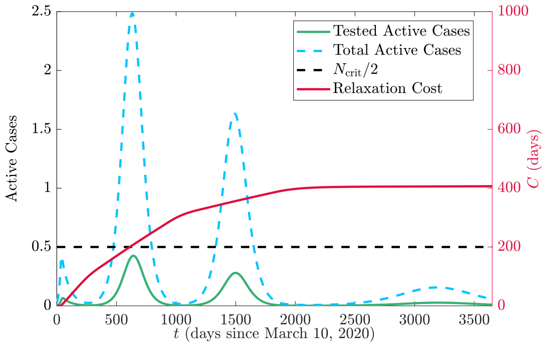


(b) $C_c^*/2$.

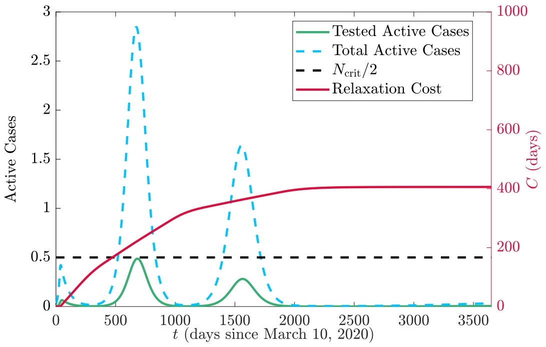


(c) $C_c^*$.

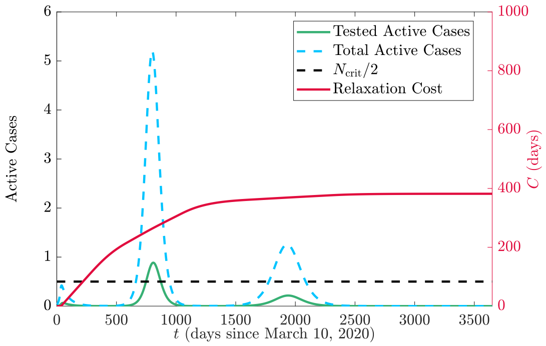


(d) $2C_c^*$.

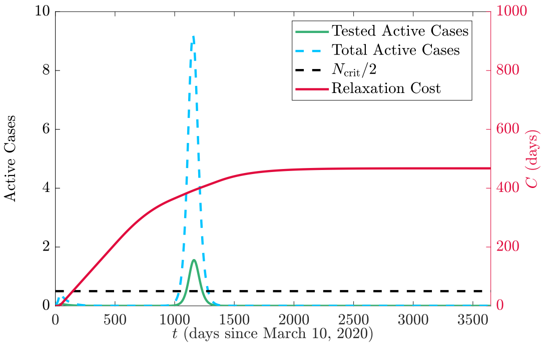


(e) $4C_c^*$.

Figure 9: Tested active cases (green solid), total active cases (blue dashed) and relaxation cost (red solid) for $\eta = 1/2$ and varying $C_c$. The grey curve represents the baseline case of no implementation of social distancing (and thus no cost) and the black dashed line is the critical threshold $N_{\rm crit}/2$. Case values are scaled by $N_{\rm crit}$.

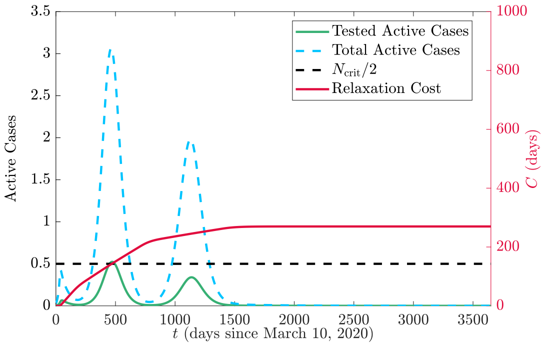


(a) $C_c^*/4$.

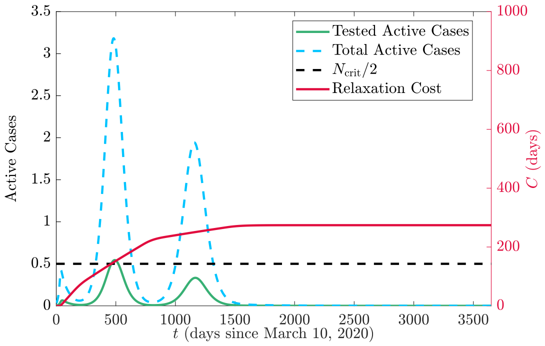


(b) $C_c^*/2$.

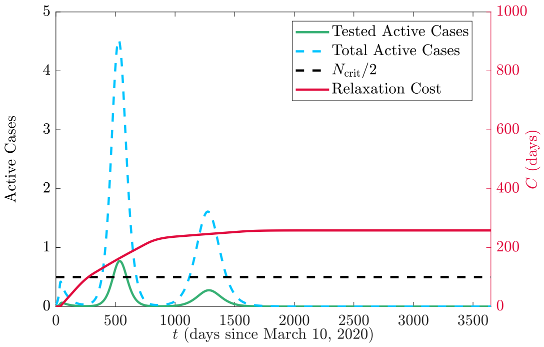


(c) $C_c^*$.

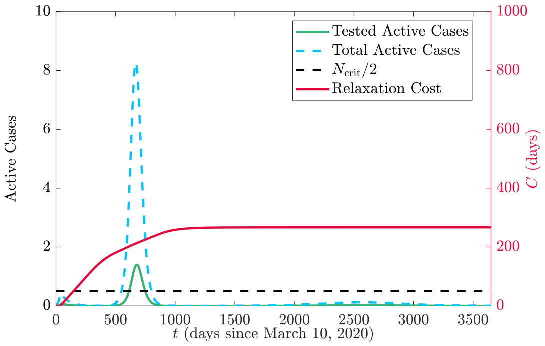


(d) $2C_c^*$.

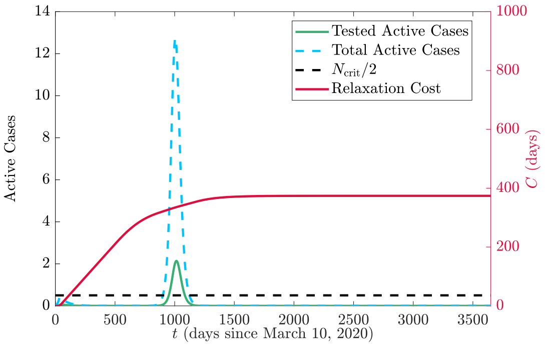


(e) $4C_c^*$.

Figure 10: Tested active cases (green solid), total active cases (blue dashed) and relaxation cost (red solid) for $\eta = 1$ and varying $C_c$. The grey curve represents the baseline case of no implementation of social distancing (and thus no cost) and the black dashed line is the critical threshold $N_{\rm crit}/2$. Case values are scaled by $N_{\rm crit}$.

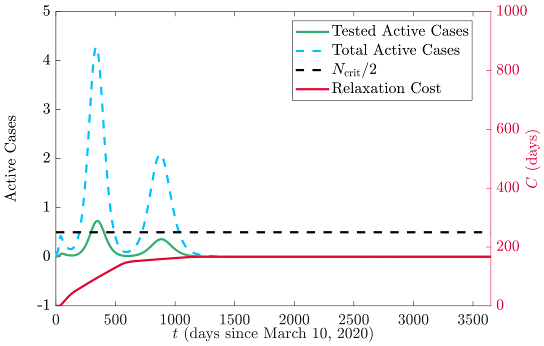


(a) $C_c^*/4$.

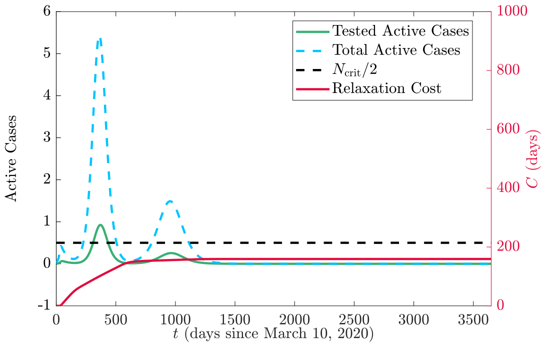


(b) $C_c^*/2$.

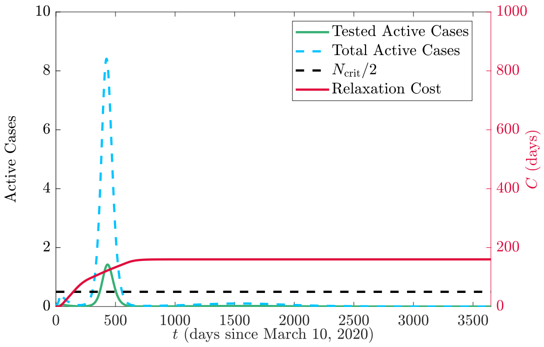


(c) $C_c^*$.

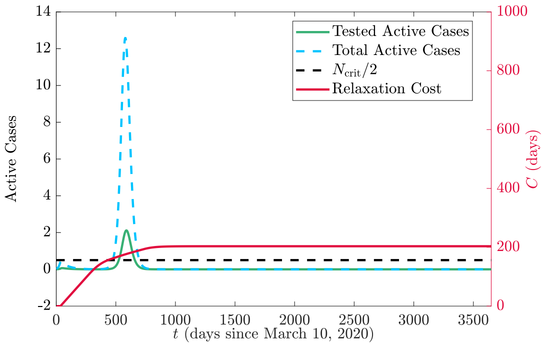


(d) $2C_c^*$.

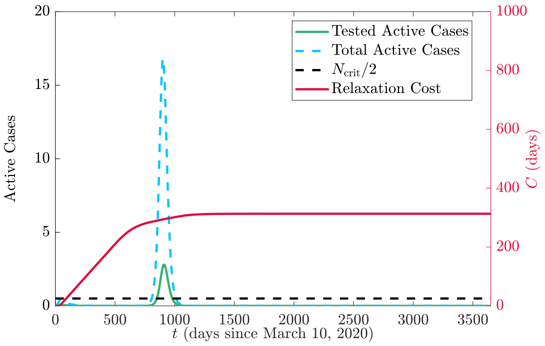


(e) $4C_c^*$.

Figure 11: Tested active cases (green solid), total active cases (blue dashed) and relaxation cost (red solid) for $\eta = 2$ and varying $C_c$. The grey curve represents the baseline case of no implementation of social distancing (and thus no cost) and the black dashed line is the critical threshold $N_{\text{crit}}/2$. Case values are scaled by $N_{\text{crit}}$.

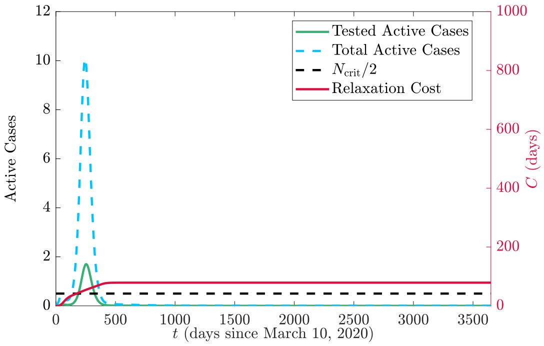


(a) $C_c^*/4$.

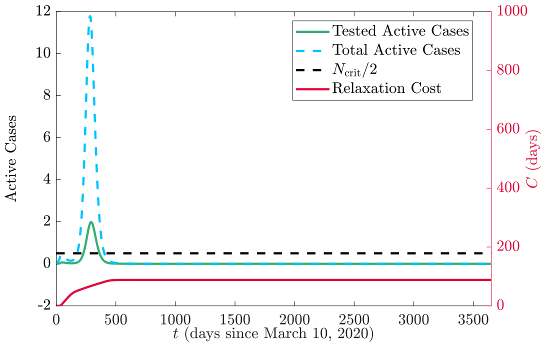


(b) $C_c^*/2$.

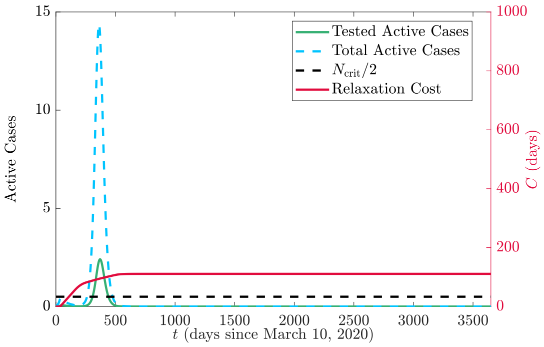


(c) $C_c^*$.

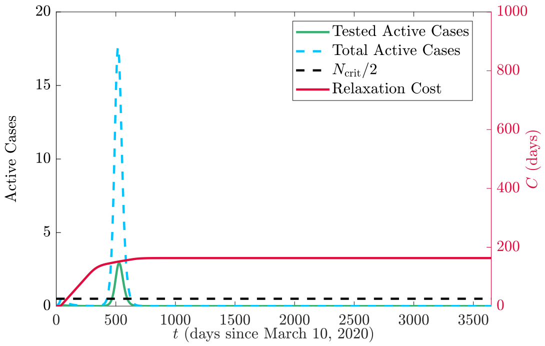


(d) $2C_c^*$.

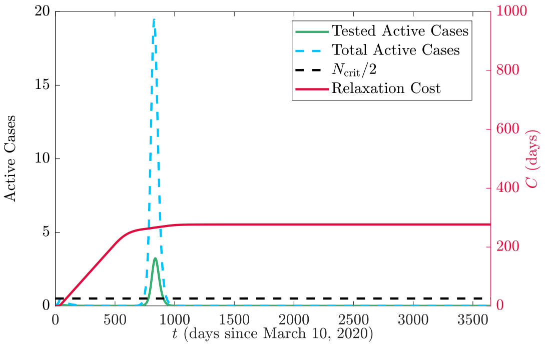


(e) $4C_c^*$.

Figure 12: Tested active cases (green solid), total active cases (blue dashed) and relaxation cost (red solid) for $\eta = 4$ and varying $C_c$. The grey curve represents the baseline case of no implementation of social distancing (and thus no cost) and the black dashed line is the critical threshold $N_{\text{crit}}/2$. Case values are scaled by $N_{\text{crit}}$.

We also plot the populations in each of the two social distancing classes for each of the 25 scenarios considered in Figure 8 of the main text. These are plotted in Figure 13 for social distancing class 1 and Figure 14 for social distancing class 2.

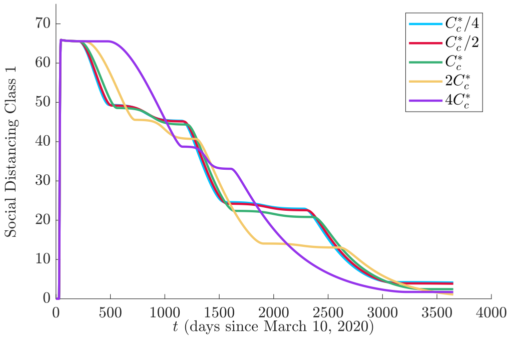


(a) $\eta/4$.

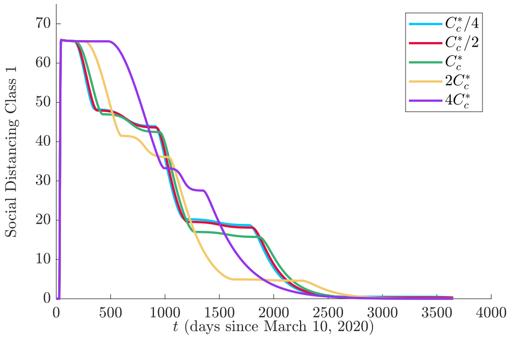


(b) $\eta/2$.

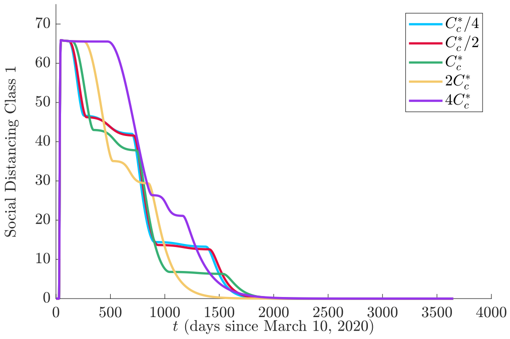


(c) $\eta$.

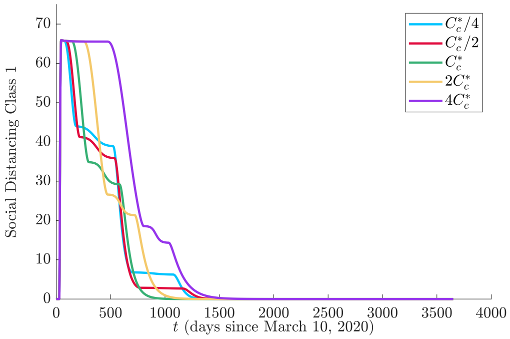


(d) $2\eta$.

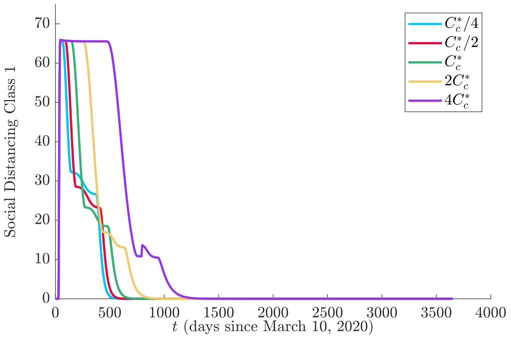


(e) $4\eta$.

Figure 13: Total people in social distancing class 1 ($S_1$, $E_1$, $P_1$, $I_{S_1}$, $I_{A_1}$) for different values of $\eta$ and $C_c$.

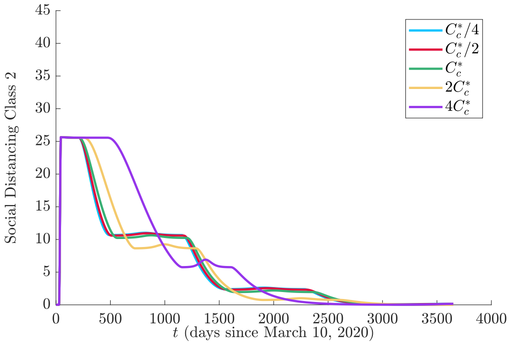


(a) $\eta/4$.

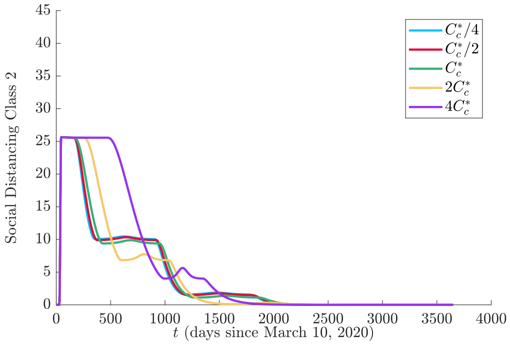


(b) $\eta/2$.

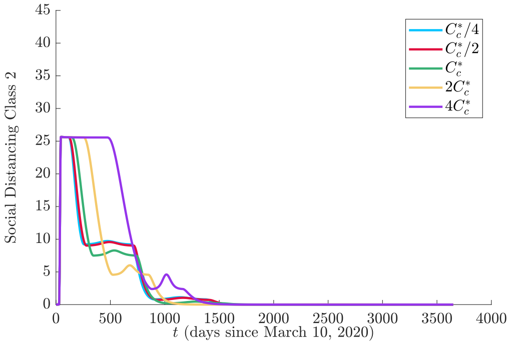


(c) $\eta$.

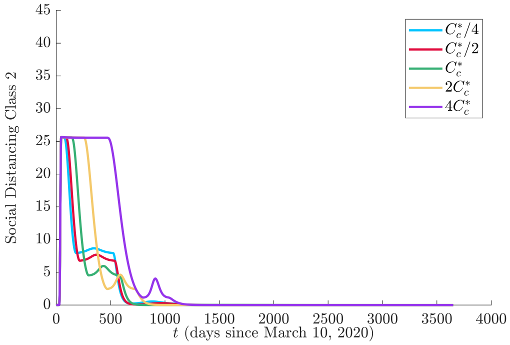


(d) $2\eta$.

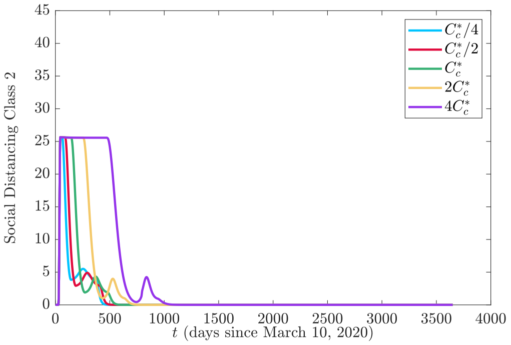


(e) $4\eta$.

Figure 14: Total people in social distancing class 2 ($S_2$, $E_2$, $P_2$, $P_M$, $I_{S_2}$, $I_{S_M}$, $I_{A_2}$, $I_{A_M}$) for different values of $\eta$ and $C_c$.
