## Supplementary material for "Cost and social distancing dynamics in a mathematical model of COVID-19 with application to Ontario, Canada": Code: covid_lsq2.m

```
function Y=covid_lsq2(v,t,Y0)


flags.model = 3;
flags.cost_control = 0;
run_params;

params.Kc = v(1);
params.K0 = v(2);
% params.Mc = v(1);
% params.M0 = v(2);

params.beta = params.R0*params.phi*params.gamma/(params.alpha*(params.gamma+params.phi)+(1-params.alpha)*params.q*params.phi);

odeopts = odeset('NonNegative',1,'RelTol',1e-8,'AbsTol',1e-9);
% odeopts = odeset('NonNegative',1);
[T,y] = ode23t(@ODEf,t,Y0,odeopts,params,flags);

PM = y(:,10);
IsM = y(:,14);
IaM = y(:,18);
M = y(:,27);

AM = PM + IsM + IaM;

Y = [AM M];
% Y = M;
end
```
