## Supplementary material for "Cost and social distancing dynamics in a mathematical model of COVID-19 with application to Ontario, Canada": Code: covid_lsq.m

```
function Y=covid_lsq(v,t)

% persistent x;
% if isempty(x)
%     x = 1;
% else
%     x = x+1;
% end
% fprintf('Computing iteration %i...\n v=%s\n',x,num2str(v'));

flags.model = 3;
flags.cost_control = 0;
run_params;

params.Kc = v(1);
params.Mc = v(2);
params.rho0 = v(3);

params.rhoI = 4*v(3);

params.K0 = 4*params.Kc;
params.M0 = 2*params.Mc;

% y0 = [params.N/params.N0-v(4);0;0;0;0;0;0;0;0;0;v(4);0;0;0;0;0;0;0;0;0;0;0;0;0;0;0;0;0];


params.beta = params.R0*params.phi*params.gamma/(params.alpha*(params.gamma+params.phi)+(1-params.alpha)*params.q*params.phi);

odeopts = odeset('NonNegative',1,'RelTol',1e-8,'AbsTol',1e-9);
% odeopts = odeset('NonNegative',1);
[T,y] = ode23t(@ODEf,t,y0,odeopts,params,flags);

PM = y(:,10);
IsM = y(:,14);
IaM = y(:,18);
M = y(:,27);

AM = PM + IsM + IaM;

Y = [AM M];
% Y = M;
end
```
