## Supplementary material for "Cost and social distancing dynamics in a mathematical model of COVID-19 with application to Ontario, Canada": Code: data_compare_plot.m

```
load('DATA-Aug18.mat');
% new_dat=load('DATA-Nov21.mat');
new_dat=load('DATA-Jan06.mat');

T = t;%t*365/12;
figure
plot(T,AM,'linewidth',2);
hold on
% plot(T,It,'--','linewidth',2);
plot(DATA_T,(DATA_pos)/params.N0,'o');
plot(new_dat.DATA_T(163:end),(new_dat.DATA_pos(163:end))/params.N0,'ko');
hold off
% xlim([0,170]);
xlim([0,500]);
ylim([0,0.35]);

figure
plot(T,M,'linewidth',2);
hold on
plot(DATA_T,(DATA_tot)/params.N0,'o');
plot(new_dat.DATA_T(163:end),(new_dat.DATA_tot(163:end))/params.N0,'ko');
hold off
xlim([0,500]);

% figure
% Tspot=find(abs(T-204)==min(abs(T-204)));
% plot(T(1:Tspot),AM(1:Tspot),'linewidth',2);
% hold on
% plot(T(Tspot:end)+30,AM(Tspot:end),'linewidth',2);
% plot(DATA_T,(DATA_pos)/params.N0,'o');
% plot(new_dat.DATA_T(163:end),(new_dat.DATA_pos(163:end))/params.N0,'ko');
% hold off
% % xlim([0,170]);
% xlim([0,500]);
% ylim([0,0.2]);
```
