## Supplementary material for "Cost and social distancing dynamics in a mathematical model of COVID-19 with application to Ontario, Canada": Code: data_fit2.m

```
close all
clear variables

run_params;
flags.cost_control=0;

%Use the extended Ontario data to fit Kc, K0

load('DATA-Jan06.mat');

dat_start = 180;
dat_end = 220;%DATA_T(end);%221;

temp_p = params;
% v0 = [params.Kc;params.K0];
v0 = [1/16.24/100 150*1/16.24/100];
% v0 = [1/16.24/100 150*1/16.24/100 params.Mc params.M0];
% v0 = [params.Kc 4*params.Kc params.Mc params.M0];
% v0 = [0.031673619726661   0.236213223621245   0.020050482377453   0.057420688812194];
v0 = [0 0.070122974212569];
% v0 = [params.Kc;params.K0];

LB = [0;0];

UB =[inf;inf];

%Setup initial simulation
flags.model = 3;
flags.cost_control = 0;
odeopts = odeset('NonNegative',1);
[t1,y1] = ode23t(@ODEf,DATA_T(1:dat_start),y0,odeopts,params,flags);

PM1 = y1(:,10);
IsM1 = y1(:,14);
IaM1 = y1(:,18);
M1 = y1(:,27);

AM1 = PM1 + IsM1 + IaM1;

options = optimoptions('lsqcurvefit','UseParallel',true);
[V,RESNORM,RESIDUAL,EXITFLAG] = lsqcurvefit(@covid_lsq2,v0,DATA_T(dat_start:dat_end),[DATA_pos(dat_start:dat_end)/params.N_crit,DATA_tot(dat_start:dat_end)/params.N_crit],LB,UB,options,y1(end,:));

%%%%%%%%%%%%%%%%%%%%%%%%%%%%%%%%%%%%%%%%%%%%%%%%%%%%%%%%%%%%%%%%%%%%%%%%%%
flags.model = 3;
params.Kc = V(1);
params.K0 = V(2);
% params.Mc = V(1);
% params.M0 = V(2);

params.beta = params.R0*params.phi*params.gamma/(params.alpha*(params.gamma+params.phi)+(1-params.alpha)*params.q*params.phi);

var_names = {'S';'S_1';'S_2';'E';'E_1';'E_2';'P';'P_1';'P_2';'P_M';'I_S';'I_{S1}';'I_{S2}';'I_{SM}';'I_A';'I_{A1}';'I_{A2}';'I_{AM}';'R_S';'R_{S1}';'R_{S2}';'R_{SM}';'R_A';'R_{A1}';'R_{A2}';'R_{AM}';'M';'C'};

odeopts = odeset('NonNegative',1,'RelTol',1e-8,'AbsTol',1e-9);
% odeopts = odeset('NonNegative',1);
[t,y] = ode23t(@ODEf,DATA_T(dat_start:dat_end),y1(end,:),odeopts,params,flags);
[t2,y2] = ode23t(@ODEf,DATA_T(dat_start:end),y1(end,:),odeopts,params,flags);

PM = y(:,10);
IsM = y(:,14);
IaM = y(:,18);
M = y(:,27);

AM = PM + IsM + IaM;
AT = y(:,7)+y(:,8)+y(:,9)+y(:,10)+y(:,11)+y(:,12)+y(:,13)+y(:,14)+y(:,15)+y(:,16)+y(:,17)+y(:,18);

PM2 = y2(:,10);
IsM2 = y2(:,14);
IaM2 = y2(:,18);
M2 = y2(:,27);

AM2 = PM2 + IsM2 + IaM2;

% for k=1:2
%     figure
%     hold on
%     for j=1:3
%         plot(t,y(:,(k-1)*3+j),'linewidth',2);
%     end
%     legend(var_names{(k-1)*3+1:(k-1)*3+3},'location','best');
%     hold off
% end
% for k = 3:5
%     figure
%     hold on
%     for j=1:4
%         plot(t,y(:,6+(k-3)*4+j),'linewidth',2);
%     end
%     legend(var_names{6+(k-3)*4+1:6+(k-3)*4+4},'location','best');
%     if k==4
%         plot(t,ones(length(t),1)*params.N_crit/params.N0,'k--','linewidth',2);
%         plot(t,y(:,11)+y(:,12)+y(:,13)+y(:,14),'k','linewidth',2);
%     end
%     hold off
% end
% figure
% yyaxis left
% plot(t,y(:,27),'linewidth',2);
% hold on
% yyaxis right
% plot(t,y(:,10)+y(:,14)+y(:,18),'linewidth',2);
% legend({var_names{27},'M_A'},'location','best');
% 
% figure
% plot(t,y(:,28),'linewidth',2);
% legend(var_names{28},'location','best');

figure
plot(t,AM,'linewidth',2);
hold on
plot(DATA_T(dat_start:dat_end),DATA_pos(dat_start:dat_end)/params.N0,'o');
hold off

figure
plot(t,M,'linewidth',2);
hold on
plot(DATA_T(dat_start:dat_end),DATA_tot(dat_start:dat_end)/params.N0,'o');
hold off

figure
plot([t1;t2],[AM1;AM2],'linewidth',2);
hold on
plot(DATA_T,DATA_pos/params.N0,'o');
hold off
```
