## Supplementary material for "Cost and social distancing dynamics in a mathematical model of COVID-19 with application to Ontario, Canada": Code: data_fit.m

```
close all
clear variables

run_params;
flags.cost_control=0;

%Use the Ontario data to fit Kc, K0, Mc, M0, and initial sick

load('DATA-Aug18.mat');

DATA_pos = DATA_pos - DATA_pos(1);
DATA_tot = DATA_tot - DATA_tot(1);

% v0 = [params.Kc;params.Mc;params.rho0];
v0 = [0.0607 0.0263 0.0095];

LB = [0;0;0];

UB =[inf;inf;inf];

options = optimoptions('lsqcurvefit','UseParallel',true);
[V,RESNORM,RESIDUAL,EXITFLAG] = lsqcurvefit(@covid_lsq,v0,DATA_T,[DATA_pos/params.N_crit,DATA_tot/params.N_crit],LB,UB,options);

%%%%%%%%%%%%%%%%%%%%%%%%%%%%%%%%%%%%%%%%%%%%%%%%%%%%%%%%%%%%%%%%%%%%%%%%%%
flags.model = 3;
params.Kc = V(1);
params.Mc = V(2);
params.rho0 = V(3);

params.rhoI = 4*V(3);
params.K0 = 4*params.Kc;
params.M0 = 2*params.Mc;

% y0 = [params.N/params.N0-V(4);0;0;0;0;0;0;0;0;0;V(4);0;0;0;0;0;0;0;0;0;0;0;0;0;0;0;0;0];


params.beta = params.R0*params.phi*params.gamma/(params.alpha*(params.gamma+params.phi)+(1-params.alpha)*params.q*params.phi);

var_names = {'S';'S_1';'S_2';'E';'E_1';'E_2';'P';'P_1';'P_2';'P_M';'I_S';'I_{S1}';'I_{S2}';'I_{SM}';'I_A';'I_{A1}';'I_{A2}';'I_{AM}';'R_S';'R_{S1}';'R_{S2}';'R_{SM}';'R_A';'R_{A1}';'R_{A2}';'R_{AM}';'M';'C'};

odeopts = odeset('NonNegative',1,'RelTol',1e-8,'AbsTol',1e-9);
% odeopts = odeset('NonNegative',1);
[t,y] = ode23t(@ODEf,DATA_T,y0,odeopts,params,flags);

PM = y(:,10);
IsM = y(:,14);
IaM = y(:,18);
M = y(:,27);

AM = PM + IsM + IaM;
AT = y(:,7)+y(:,8)+y(:,9)+y(:,10)+y(:,11)+y(:,12)+y(:,13)+y(:,14)+y(:,15)+y(:,16)+y(:,17)+y(:,18);

for k=1:2
    figure
    hold on
    for j=1:3
        plot(t,y(:,(k-1)*3+j),'linewidth',2);
    end
    legend(var_names{(k-1)*3+1:(k-1)*3+3},'location','best');
    hold off
end
for k = 3:5
    figure
    hold on
    for j=1:4
        plot(t,y(:,6+(k-3)*4+j),'linewidth',2);
    end
    legend(var_names{6+(k-3)*4+1:6+(k-3)*4+4},'location','best');
    if k==4
        plot(t,ones(length(t),1)*params.N_crit/params.N0,'k--','linewidth',2);
        plot(t,y(:,11)+y(:,12)+y(:,13)+y(:,14),'k','linewidth',2);
    end
    hold off
end
figure
yyaxis left
plot(t,y(:,27),'linewidth',2);
hold on
yyaxis right
plot(t,y(:,10)+y(:,14)+y(:,18),'linewidth',2);
legend({var_names{27},'M_A'},'location','best');

figure
plot(t,y(:,28),'linewidth',2);
legend(var_names{28},'location','best');

figure
plot(t,AM,'linewidth',2);
hold on
plot(DATA_T,(DATA_pos)/params.N0,'o');
hold off

figure
plot(t,M,'linewidth',2);
hold on
plot(DATA_T,(DATA_tot)/params.N0,'o');
hold off
```
