## Supplementary material for "Cost and social distancing dynamics in a mathematical model of COVID-19 with application to Ontario, Canada": Code: figure_revise.m

```
function figure_revise(pos,plotpath,fig,figprop)


for k=1:length(fig)
    fnt_size = figprop(k).fnt_size;
    if ~isempty(figprop(k).ax)
        set(figprop(k).ax,'TickLabelInterpreter','LaTeX','FontSize',fnt_size);
    end
    if ~isempty(figprop(k).title)
        set(figprop(k).title,'Interpreter','LaTeX','FontSize',fnt_size,'Units','normalized');
    end
    if ~isempty(figprop(k).xlab)
        set(figprop(k).xlab,'Interpreter','LaTeX','FontSize',fnt_size,'Units','normalized');
        set(figprop(k).xlab,'position',[0.5;-0.065;0]);
    end
    if ~isempty(figprop(k).ylab)
        set(figprop(k).ylab,'Interpreter','LaTeX','FontSize',fnt_size,'Units','normalized');
        if ~isempty(figprop(k).ypos)
            set(figprop(k).ylab,'position',ypos{k});
        else
            set(figprop(k).ylab,'position',[-0.08;0.5;0]);
        end
    end
    if ~isempty(figprop(k).ylab2)
        set(figprop(k).ylab2,'Interpreter','LaTeX','FontSize',fnt_size,'Units','normalized');
        if ~isempty(figprop(k).ypos2)
            set(figprop(k).ylab2,'position',ypos2{k});
        else
            set(figprop(k).ylab2,'position',[1.075;0.5;0]);
        end
    end
    if ~isempty(figprop(k).leg)
        fnt_size=14;
        set(figprop(k).leg,'Interpreter','LaTeX','FontSize',fnt_size);
        fnt_size=16;
    end
    if ~isempty(figprop(k).ax2)
        set(figprop(k).ax2,'TickLabelInterpreter','LaTeX','FontSize',fnt_size);
    end
    if ~isempty(figprop(k).cb)
        set(figprop(k).cb,'TickLabelInterpreter','LaTeX','fontsize',fnt_size);
        set(get(figprop(k).cb,'Label'),'Interpreter','LaTeX')
    end
    if strcmp(figprop(k).type,'figure')
        box on
    end
    
    set(fig(k),'paperposition',[0,0,pos(3),pos(4)]);
    set(fig(k),'papersize',[pos(3),pos(4)]);
    hgexport(fig(k), strcat(plotpath,get(fig(k),'name')), hgexport('factorystyle'), 'Format', 'pdf');
end

end
```
