## Supplementary material for "Cost and social distancing dynamics in a mathematical model of COVID-19 with application to Ontario, Canada": Code: generate_media.m

```
close all
clear variables

process_data_2 = 0;
process_data_5 = 0;
process_data_8 = 0;
process_data_9 = 0;
process_data_10 = 0;

loop_flag = 0;

%% BASIC SETUP-DO NOT NEED TO EDIT GENERALLY
pcols = {1/255*[0,196,255],1/255*[225,12,62],1/255*[55,176,116],1/255*[244,201,107],1/255*[149,52,235],1/255*[176,176,176]};

curpath = pwd;
newpath = strcat(curpath(1:length(curpath)-6),'\data_figs\');
plotpath = strcat(curpath(1:length(curpath)-6),'\plots\');
runpath = strcat(curpath,'\runs\');

pos = [2,4.5,6,4.5];

exp_len = 10;
sub_len = [0;0;4;0;0;3;0;0;4;3;0];
%% 

%0 - mu and nu graphs
%1 - comparison of nothing, just quarantine, and social distancing
%2 - runs with the parameter changes
%3 - q study with no intervention
%4 - sensitivity analysis graphs
%5 - same as 2 but with varying rho
%6 - data fitting curves
%7 - eta demonstration
%8 - eta runs with heat maps etc.
%9 - same as 2 but with increasing Mc even more
%10 - Plot of health cost with varying Mc


done = 0;
experiment =6;
sub_experiment = 0;
err_bar_flag = 0;

if loop_flag
    experiment =2;
    sub_experiment =0;
end

while ~done
    close all
    clear fig
    clr_flag = 0;
    if ~loop_flag
        done = 1;
    else
        fprintf('Currently running experiment %i sub experiment %i...\n',experiment,sub_experiment);
    end

    switch experiment
        case 0
            M = linspace(0,1,10000);
            fig(1) = new_figure('mu_fig',pos);
            mumax = 2;
            M0 = 0.25;
            M1 = 0.75;
            mu = mumax*tanh(atanh(0.9)/(M1-M0)*max(M-M0,0));
            mplot = plot(M,mu,'Color',pcols{1},'linewidth',2);
            xlab{1} = xlabel('$M$');
            ylab{1} = ylabel('$\mu$');
            ax{1} = gca;

            fig(2) = new_figure('nu_fig',pos);
            numin = 1;
            numax = 2;
            N0 = 0.25;
            N1 = 0.75;
            nu = -(numax-numin)*tanh(atanh(1-0.1*numin/(numax-numin))/(N1-N0)*max(M-N0,0))+numax;
            nplot = plot(M,nu,'Color',pcols{1},'linewidth',2);
            xlab{2} = xlabel('$M$');
            ylab{2} = ylabel('$\nu$');
            ax{2} = gca;
        case 1
            N = 13448494; %population
            N_crit = 655*10; %critical icu capacity
            DATAS1 = load(strcat(runpath,'case1.mat'));
            DATAS2 = load(strcat(runpath,'case2.mat'));
            DATAS3 = load(strcat(runpath,'case3.mat'));

            y_fact = 100;
            fig(1) = new_figure('I_S',pos);
            p1=plot(DATAS1.t,y_fact*DATAS1.y(:,10),'Color',pcols{1},'linewidth',2,'linestyle','-');
            hold on
            p2=plot(DATAS2.t,y_fact*DATAS2.y(:,10),'Color',pcols{2},'linewidth',2,'linestyle','-');
            plot(DATAS2.t,y_fact*DATAS2.y(:,11),'Color',pcols{2},'linewidth',2,'linestyle','--');
            plot(DATAS2.t,y_fact*DATAS2.y(:,12),'Color',pcols{2},'linewidth',2,'linestyle','-.');
            p3=plot(DATAS3.t,y_fact*DATAS3.y(:,10),'Color',pcols{3},'linewidth',2,'linestyle','-');
            plot(DATAS3.t,y_fact*DATAS3.y(:,11),'Color',pcols{3},'linewidth',2,'linestyle','--');
            plot(DATAS3.t,y_fact*DATAS3.y(:,12),'Color',pcols{3},'linewidth',2,'linestyle','-.');
            p4=plot(DATAS1.t,y_fact*ones(length(DATAS1.t),1)*N_crit/N,'k--','linewidth',2);
            hold off
            xlab{1} = xlabel('$t$ (months)');
            ylab{1} = ylabel('$I_{S} \times 10^{-2}$');
            leg{1} = legend([p1,p2,p3,p4],'Case 1','Case 2','Case 3','$n_{\rm crit}$','location','northeast');
            ax{1} = gca;

            y_fact = 100;
            fig(2) = new_figure('I_A',pos);
            p1=plot(DATAS1.t,y_fact*DATAS1.y(:,13),'Color',pcols{1},'linewidth',2,'linestyle','-');
            hold on
            p2=plot(DATAS2.t,y_fact*DATAS2.y(:,13),'Color',pcols{2},'linewidth',2,'linestyle','-');
            p3=plot(DATAS3.t,y_fact*DATAS3.y(:,13),'Color',pcols{3},'linewidth',2,'linestyle','-');
            plot(DATAS3.t,y_fact*DATAS3.y(:,14),'Color',pcols{3},'linewidth',2,'linestyle','--');
            plot(DATAS3.t,y_fact*DATAS3.y(:,15),'Color',pcols{3},'linewidth',2,'linestyle','-.');
            hold off
            xlab{2} = xlabel('$t$ (months)');
            ylab{2} = ylabel('$I_{A} \times 10^{-2}$');
            leg{2} = legend([p1,p2,p3],'Case 1','Case 2','Case 3','location','northeast');
            ax{2} = gca;

            y_fact = 100;
            fig(3) = new_figure('I_S_zoom',pos);
            hold on
            p2=plot(DATAS2.t,y_fact*DATAS2.y(:,10),'Color',pcols{2},'linewidth',2,'linestyle','-');
            plot(DATAS2.t,y_fact*DATAS2.y(:,11),'Color',pcols{2},'linewidth',2,'linestyle','--');
            plot(DATAS2.t,y_fact*DATAS2.y(:,12),'Color',pcols{2},'linewidth',2,'linestyle','-.');
            p3=plot(DATAS3.t,y_fact*DATAS3.y(:,10),'Color',pcols{3},'linewidth',2,'linestyle','-');
            plot(DATAS3.t,y_fact*DATAS3.y(:,11),'Color',pcols{3},'linewidth',2,'linestyle','--');
            plot(DATAS3.t,y_fact*DATAS3.y(:,12),'Color',pcols{3},'linewidth',2,'linestyle','-.');
            p4=plot(DATAS1.t,y_fact*ones(length(DATAS1.t),1)*N_crit/N,'k--','linewidth',2);
            hold off
            xlab{3} = xlabel('$t$ (months)');
            ylab{3} = ylabel('$I_{S} \times 10^{-2}$');
            leg{3} = legend([p2,p3,p4],'Case 2','Case 3','$n_{\rm crit}$','location','northeast');
            ax{3} = gca;

            y_fact = 1;
            fig(4) = new_figure('S',pos);
            p1=plot(DATAS1.t,y_fact*DATAS1.y(:,1),'Color',pcols{1},'linewidth',2,'linestyle','-');
            hold on
            p2=plot(DATAS2.t,y_fact*DATAS2.y(:,1),'Color',pcols{2},'linewidth',2,'linestyle','-');
            p3=plot(DATAS3.t,y_fact*DATAS3.y(:,1),'Color',pcols{3},'linewidth',2,'linestyle','-');
            plot(DATAS3.t,y_fact*DATAS3.y(:,2),'Color',pcols{3},'linewidth',2,'linestyle','--');
            plot(DATAS3.t,y_fact*DATAS3.y(:,3),'Color',pcols{3},'linewidth',2,'linestyle','-.');
            hold off
            xlab{4} = xlabel('$t$ (months)');
            ylab{4} = ylabel('$S$');
            leg{4} = legend([p1,p2,p3],'Case 1','Case 2','Case 3','location','northeast');
            ax{4} = gca;

            fig(5) = new_figure('sick',pos);
            sick1 = sum(DATAS1.y(:,10:21),2);
            sick2 = sum(DATAS2.y(:,10:21),2);
            sick3 = sum(DATAS3.y(:,10:21),2);
            plot(DATAS1.t,sick1,'Color',pcols{1},'linewidth',2);
            hold on
            plot(DATAS2.t,sick2,'Color',pcols{2},'linewidth',2);
            plot(DATAS3.t,sick3,'Color',pcols{3},'linewidth',2);
            hold off
            xlab{5} = xlabel('$t$ (months');
            ylab{5} = ylabel('sick');
            leg{5} = legend('Case 1','Case 2','Case 3','location','southeast');
            ax{5} = gca;
        case 2
            q = [0.9;0.69;0.27];
            multiplier = [1/4;1/2;1;2;4];
            len_q = length(q);
            len_m = length(multiplier);
            fprintf('Loading Data...\n');
            for k=1:len_q
    %             K = (k-1)*(len_m^2+1)+1;
                file_title=strcat('odemodel-3-q-',num2str(floor(100*q(k))),'-case0.mat');
                DATAS = load(strcat(runpath,file_title));
                params{k,1} = DATAS.params;
                t{k,1} = DATAS.t;
                y = DATAS.y;
                S{k,1} = y(:,1);
                S1{k,1} = y(:,2);
                S2{k,1} = y(:,3);
                E{k,1} = y(:,4);
                E1{k,1} = y(:,5);
                E2{k,1} = y(:,6);
                P{k,1} = y(:,7);
                P1{k,1} = y(:,8);
                P2{k,1} = y(:,9);
                PM{k,1} = y(:,10);
                Is{k,1} = y(:,11);
                Is1{k,1} = y(:,12);
                Is2{k,1} = y(:,13);
                IsM{k,1} = y(:,14);
                Ia{k,1} = y(:,15);
                Ia1{k,1} = y(:,16);
                Ia2{k,1} = y(:,17);
                IaM{k,1} = y(:,18);
                Rs{k,1} = y(:,19);
                Rs1{k,1} = y(:,20);
                Rs2{k,1} = y(:,21);
                RsM{k,1} = y(:,22);
                Ra{k,1} = y(:,23);
                Ra1{k,1} = y(:,24);
                Ra2{k,1} = y(:,25);
                RaM{k,1} = y(:,26);
                M{k,1} = y(:,27);
                C{k,1} = y(:,28);
                for j1=1:len_m
                    for j2=1:len_m
                        K2 = 1 + (j1-1)*len_m + j2;
                        file_title=strcat('odemodel-3-q-',num2str(floor(100*q(k))),'-case',num2str((j1-1)*len_m+j2),'.mat');
                        DATAS = load(strcat(runpath,file_title));
                        params{k,K2} = DATAS.params;
                        t{k,K2} = DATAS.t;
                        y = DATAS.y;
                        S{k,K2} = y(:,1);
                        S1{k,K2} = y(:,2);
                        S2{k,K2} = y(:,3);
                        E{k,K2} = y(:,4);
                        E1{k,K2} = y(:,5);
                        E2{k,K2} = y(:,6);
                        P{k,K2} = y(:,7);
                        P1{k,K2} = y(:,8);
                        P2{k,K2} = y(:,9);
                        PM{k,K2} = y(:,10);
                        Is{k,K2} = y(:,11);
                        Is1{k,K2} = y(:,12);
                        Is2{k,K2} = y(:,13);
                        IsM{k,K2} = y(:,14);
                        Ia{k,K2} = y(:,15);
                        Ia1{k,K2} = y(:,16);
                        Ia2{k,K2} = y(:,17);
                        IaM{k,K2} = y(:,18);
                        Rs{k,K2} = y(:,19);
                        Rs1{k,K2} = y(:,20);
                        Rs2{k,K2} = y(:,21);
                        RsM{k,K2} = y(:,22);
                        Ra{k,K2} = y(:,23);
                        Ra1{k,K2} = y(:,24);
                        Ra2{k,K2} = y(:,25);
                        RaM{k,K2} = y(:,26);
                        M{k,K2} = y(:,27);
                        C{k,K2} = y(:,28);
                    end
                end
            end
            fprintf('Data Loaded...\n');

%%%%%%%%%%%%% Now do some processing on the data
            if process_data_2 == 0 %If looping only process the data once
                n_crit = 1/2; %Value for which we surpass health resources
                omega = 0.9; %Cost weighting
                for j=1:len_q
                    for k=1:len_m^2+1
                        It{j,k} = Is{j,k}+Is1{j,k}+Is2{j,k}+IsM{j,k};
                        IAt{j,k} = Ia{j,k}+Ia1{j,k}+Ia2{j,k}+IaM{j,k};
                        SD1{j,k} = S1{j,k}+E1{j,k}+P1{j,k}+Is1{j,k}+Ia1{j,k};
                        SD2{j,k} = S2{j,k}+E2{j,k}+P2{j,k}+PM{j,k}+Is2{j,k}+IsM{j,k}+Ia2{j,k}+IaM{j,k};
                        Pt{j,k} = P{j,k}+P1{j,k}+P2{j,k}+PM{j,k};
                        peak_time(j,k) = t{j,k}(It{j,k}==max(It{j,k}));
                        peak_val(j,k) = max(It{j,k});
                        sickS{j,k} = (Is{j,k}+Is1{j,k}+Is2{j,k}+IsM{j,k}+Rs{j,k}+Rs1{j,k}+Rs2{j,k}+RsM{j,k})*params{j,k}.N0/params{j,k}.N;
                        sickA{j,k} = (Ia{j,k}+Ia1{j,k}+Ia2{j,k}+IaM{j,k}+Ra{j,k}+Ra1{j,k}+Ra2{j,k}+RaM{j,k})*params{j,k}.N0/params{j,k}.N;
                        sick{j,k} = (sickS{j,k} + sickA{j,k});

                        KM{j,k} = 1/log(2)*max((params{j,k}.rho0*(Ia{j,k}+Ia1{j,k}+Ia2{j,k}+P{j,k}+P1{j,k}+P2{j,k}) + params{j,k}.rhoI*(Is{j,k}+Is1{j,k}+Is2{j,k}))./M{j,k},0);
                        AM{j,k} = PM{j,k} + IsM{j,k} + IaM{j,k};
                        Kfun{j,k} = max(KM{j,k}-params{j,k}.Kc,0)./(max(KM{j,k}-params{j,k}.Kc,0)+params{j,k}.K0-params{j,k}.Kc);
                        Afun{j,k} = max(AM{j,k}-params{j,k}.Mc,0)./(max(AM{j,k}-params{j,k}.Mc,0)+params{j,k}.M0-params{j,k}.Mc);
                        mu{j,k} = params{j,k}.mumax*Kfun{j,k}.*Afun{j,k};
                        nu{j,k} = params{j,k}.numax*max(C{j,k}-params{j,k}.Cc,0)./(max(C{j,k}-params{j,k}.Cc,0)+params{j,k}.C0-params{j,k}.Cc);

                        %Find spike times for infection
                        donespikes = 0;
                        Itcount(j,k) = 0;
                        sgnIt = 1;
                        while(~donespikes)
                            sgnIt = sgnIt-1+find(sign(It{j,k}(sgnIt:end)-n_crit)==1,1);
                            if isempty(sgnIt)
                                donespikes=1;
                            else
                                Itcount(j,k) = Itcount(j,k)+1;
                                Itspots0{j,k}(Itcount(j,k)) = sgnIt;
                                tIt0{j,k}(Itcount(j,k))= t{j,k}(sgnIt);
                                sgnIt = sgnIt-1+find(sign(It{j,k}(sgnIt:end)-n_crit)==-1,1);

                                if isempty(sgnIt)
                                    Itspots1{j,k}(Itcount(j,k)) = length(It{j,k});
                                    tIt1{j,k}(Itcount(j,k)) = t{j,k}(end);
                                    t_cut_flag(j,k) = 1;
                                else
                                    Itspots1{j,k}(Itcount(j,k)) = sgnIt;
                                    tIt1{j,k}(Itcount(j,k)) = t{j,k}(sgnIt);
                                    t_cut_flag(j,k) = 0;
                                end

                            end
                        end
                        donespikes = 0;
                        AMcount(j,k) = 0;
                        sgnAM = 1;
                        while(~donespikes)
                            sgnAM = sgnAM-1+find(sign(AM{j,k}(sgnAM:end)-n_crit)==1,1);
                            if isempty(sgnAM)
                                donespikes=1;
                            else
                                AMcount(j,k) = AMcount(j,k)+1;
                                AMspots0{j,k}(AMcount(j,k)) = sgnAM;
                                tAM0{j,k}(AMcount(j,k))= t{j,k}(sgnAM);
                                sgnAM = sgnAM-1+find(sign(AM{j,k}(sgnAM:end)-n_crit)==-1,1);

                                if isempty(sgnAM)
                                    AMspots1{j,k}(AMcount(j,k)) = length(AM{j,k});
                                    tAM1{j,k}(AMcount(j,k)) = t{j,k}(end);
                                    t_AMcut_flag(j,k) = 1;
                                else
                                    AMspots1{j,k}(AMcount(j,k)) = sgnAM;
                                    tAM1{j,k}(AMcount(j,k)) = t{j,k}(sgnAM);
                                    t_AMcut_flag(j,k) = 0;
                                end

                            end
                        end

                        health_cost(j,k) = 0;
                        if AMcount(j,k)~=0
                            for k2=1:length(tAM0{j,k})
                                health_cost(j,k) = health_cost(j,k) + trapz(t{j,k}(AMspots0{j,k}(k2):AMspots1{j,k}(k2)),AM{j,k}(AMspots0{j,k}(k2):AMspots1{j,k}(k2)));
                            end
                        end
                        rel_health_cost(j,k) = health_cost(j,k)/health_cost(j,1);
                        cost(j,k) = C{j,k}(end);
                    end
                    max_cost(j) = max(cost(j,:));
                    rel_cost(j,:) = cost(j,:)/max_cost(j);
                end
%                 max_cost = max(max(cost));
%                 rel_cost = cost/max_cost;
                total_cost = omega*rel_health_cost+(1-omega)*rel_cost;
                for k1=2:len_m^2+1
                    for k2=k1:len_m^2+1
                        rel_total_cost(k1-1,k2-1) = total_cost(2,k2)/total_cost(2,k1);
                    end
                end
                process_data_2 = 1;
            end
%%%%%%%%%%%DONE PROCESSING

            %sub_experiment is instead of all plots, go through the ones we
            %want
            % 0 - It graphs plotted with a critical threshold
            % 1 - heatmap graphs of costs
            % 2 - sick people graphs
            % 3 - peak sick time heat map
            % 4 - Social distancing plots
            

            switch sub_experiment

                case 0
                    pos = [2,4.5,7,4.5];
                    for k=1:(len_m^2+1)
                        [fig(k),figprop(k)] = new_figure(strcat('I_t_case_',num2str(k-1)),pos);
                        temp_It{1} = spline(t{1,k},It{1,k}+IAt{1,k}+Pt{1,k},t{2,k});
                        temp_It{2} = spline(t{3,k},It{3,k}+IAt{3,k}+Pt{3,k},t{2,k});

                        err_bar{k} = [max([temp_It{1} It{2,k}+IAt{2,k}+Pt{2,k} temp_It{2}],[],2)'-(It{2,k}+IAt{2,k}+Pt{2,k})';(It{2,k}+IAt{2,k}+Pt{2,k})'];
                        
                        temp_AM{1} = spline(t{1,k},AM{1,k},t{2,k});
                        temp_AM{2} = spline(t{3,k},AM{3,k},t{2,k});
                        
                        err_barAM{k} = [max([temp_AM{1} AM{2,k} temp_AM{2}],[],2)'-AM{2,k}';AM{2,k}'];
                        
                        temp_It0{1} = spline(t{1,1},It{1,1}+IAt{1,1}+Pt{1,1},t{2,1});
                        temp_It0{2} = spline(t{3,1},It{3,1}+IAt{3,1}+Pt{3,1},t{2,1});
                        
                        err_bar0{k} = [max([temp_It0{1} It{2,1}+IAt{2,1}+Pt{2,1} temp_It0{2}],[],2)'-(It{2,1}+IAt{2,1}+Pt{2,1})';(It{2,1}+IAt{2,1}+Pt{2,1})'];

                        yyaxis left
                        
                        if err_bar_flag
                            shadedErrorBar(t{2,k},AM{2,k},err_barAM{k},'lineprops',{'Color',pcols{3},'linewidth',2});
                            hold on
                            shadedErrorBar(t{2,k},It{2,k}+IAt{2,k}+Pt{2,k},err_bar{k},'lineprops',{'Color',pcols{1},'linewidth',2});
                            shadedErrorBar(t{2,1},It{2,1}+IAt{2,1}+Pt{2,1},err_bar0{k},'lineprops',{'LineStyle','-.','Color',pcols{end},'linewidth',2});
                        else
                            plot(t{2,k},AM{2,k},'linewidth',2,'Color',pcols{3});
                            hold on
                            plot(t{2,k},It{2,k}+IAt{2,k}+Pt{2,k},'--','linewidth',2,'Color',pcols{1});
                            plot(t{2,1},It{2,1}+IAt{2,1}+Pt{2,1},'-.','linewidth',2,'Color',pcols{end});
                        end
                        plot(t{2,k},n_crit*ones(length(t{2,k}),1),'k--','linewidth',2);
                        hold off
                        ylim([0,36]);
                        figprop(k).ylab = ylabel('Active Cases');
                        figprop(k).ax = gca;
                        figprop(k).fnt_size =14;
                        set(gca,'ycolor','k');
                        
                        yyaxis right

                        temp_C{1} = spline(t{1,k},C{1,k},t{2,k});
                        temp_C{2} = spline(t{3,k},C{3,k},t{2,k});

                        err_barC{k} = [max([temp_C{1} C{2,k} temp_C{2}],[],2)'-C{2,k}';C{2,k}'-min([temp_C{1} C{2,k} temp_C{2}],[],2)'];
                        
                        if err_bar_flag
                            shadedErrorBar(t{2,k},C{2,k},err_barC{k},'lineprops',{'Color',pcols{2},'linewidth',2});
                        else
                            plot(t{2,k},C{2,k},'linewidth',2,'Color',pcols{2});
                        end
                        ylim([0,250]);

                        figprop(k).ylab2 = ylabel('$C$ (days)');
                        
                        xlim([0,3*365]);
                        figprop(k).xlab = xlabel('$t$ (days since March 10, 2020)');
                        figprop(k).leg = legend({'Tested Active Cases';'Total Active Cases';'Baseline';'$N_{\rm crit}/2$';'Relaxation Cost'},'location','best');
                        set(gca,'ycolor',pcols{2});
                    end
                case 1
                    fig_num = 0;
                    M_names={'M_c^*/4';'M_c^*/2';'M_c^*';'2M_c^*';'4M_c^*'};
                    c_names={'C_c^*/4';'C_c^*/2';'C_c^*';'2C_c^*';'4C_c^*'};
                    row_names={'M';'c';'health_cost';'cost';'total_cost'};
                    for k=1:len_q
                        for k1=1:len_m
                            heat_health_data{k}(k1,:)=rel_health_cost(k,(k1-1)*len_m+2:(k1-1)*len_m+6);
                            heat_cost_data{k}(k1,:)=rel_cost(k,(k1-1)*len_m+2:(k1-1)*len_m+6);
                            heat_tot_data{k}(k1,:)=total_cost(k,(k1-1)*len_m+2:(k1-1)*len_m+6);

                        end
                        Omega = linspace(0,1,11);
                        for K = 1:length(Omega)
                            fig_num = fig_num + 1;
                            [fig(fig_num),figprop(fig_num)] = new_figure(strcat('heat_tot_j_',num2str(100*q(k)),'-',num2str(Omega(K)*100),'-',num2str((1-Omega(K))*100)),pos);
                            heatmap(c_names,M_names,Omega(K)*heat_health_data{k}+(1-Omega(K))*heat_cost_data{k});
                            cmap = hot;
                            colormap(flipud(cmap));
                            figprop(fig_num).type='heat_map';
                            if K == 1
                                annotation('textarrow', [0.05,0.05], [0.3,0.8],'String' , ['Increase' newline 'Vigilance']);
                                annotation('textarrow', [0.3,0.8], [0.95,0.95],'String' , ['Increase Spending']);
                            end
                        end
                    end
                    fig_num = fig_num + 1;
                    [fig(fig_num),figprop(fig_num)] = new_figure('active_compare',pos);
                    plot(t{2,3},AM{2,3},'linewidth',2,'Color',pcols{1});
                    hold on
                    plot(t{2,4},AM{2,4},'linewidth',2,'Color',pcols{2});
                    line([tAM0{2,3},tAM1{2,3}],[0.5,0.5],'Color',pcols{1},'Linestyle','-','linewidth',2);
                    line([tAM0{2,3},tAM0{2,3}],[0.4,0.6],'Color',pcols{1},'Linestyle','-','linewidth',2);
                    line([tAM1{2,3},tAM1{2,3}],[0.4,0.6],'Color',pcols{1},'Linestyle','-','linewidth',2);
                    line([tAM0{2,4},t{2,4}(AMspots1{2,4}(1)-1)],[0.5,0.5],'Color',pcols{2},'Linestyle','-','linewidth',2);
                    line([tAM0{2,4},tAM0{2,4}],[0.4,0.6],'Color',pcols{2},'Linestyle','-','linewidth',2);
                    line([t{2,4}(AMspots1{2,4}(1)-1),t{2,4}(AMspots1{2,4}(1)-1)],[0.4,0.6],'Color',pcols{2},'Linestyle','-','linewidth',2);
                    
                    AM1maxX=find(AM{2,3}==max(AM{2,3}));
                    AM2maxX=find(AM{2,4}==max(AM{2,4}));
                    plot(t{2,3}(AM1maxX),max(AM{2,3}),'k.');
                    plot(t{2,4}(AM2maxX),max(AM{2,4}),'k.');


                    hold off
                    text(226,0.7,['$',num2str(round(tAM1{2,3}-tAM0{2,3},2)),'$'],'interpreter','latex','Color',pcols{1})
                    text(326,0.7,['$',num2str(round(tAM1{2,4}-tAM0{2,4},2)),'$'],'interpreter','latex','Color',pcols{2})
                    text(t{2,3}(AM1maxX),AM{2,3}(AM1maxX)+0.1,['$',num2str(round(max(AM{2,3}),2)),'$'],'interpreter','latex','color',pcols{1})
                    text(t{2,4}(AM2maxX),AM{2,4}(AM2maxX)+0.1,['$',num2str(round(max(AM{2,4}),2)),'$'],'interpreter','latex','color',pcols{2})
                    ylim([0,4.25]);
                    xlim([175,385]);
                    figprop(fig_num).xlab = xlabel('$t$ (days since March 10)');
                    figprop(fig_num).ylab = ylabel('Active Cases');
                    figprop(fig_num).leg = legend('$C_c^*/2$','$C_c^*$','location','northeast');
                    figprop(fig_num).ax = gca;
                    
                    fig_num = fig_num + 1;
                    [fig(fig_num),figprop(fig_num)] = new_figure('active_compare_2',pos);
                    plot(t{2,2},AM{2,2},'linewidth',2,'Color',pcols{1});
                    hold on
                    plot(t{2,22},AM{2,22},'linewidth',2,'Color',pcols{2});
                    line([tAM0{2,2},tAM1{2,2}],[0.5,0.5],'Color',pcols{1},'Linestyle','-','linewidth',2);
                    line([tAM0{2,2},tAM0{2,2}],[0.4,0.6],'Color',pcols{1},'Linestyle','-','linewidth',2);
                    line([tAM1{2,2},tAM1{2,2}],[0.4,0.6],'Color',pcols{1},'Linestyle','-','linewidth',2);
                    line([tAM0{2,22},t{2,22}(AMspots1{2,22}(1)-1)],[0.5,0.5],'Color',pcols{2},'Linestyle','-','linewidth',2);
                    line([tAM0{2,22},tAM0{2,22}],[0.4,0.6],'Color',pcols{2},'Linestyle','-','linewidth',2);
                    line([t{2,22}(AMspots1{2,22}(1)-1),t{2,22}(AMspots1{2,22}(1)-1)],[0.4,0.6],'Color',pcols{2},'Linestyle','-','linewidth',2);
                    
                    AM1maxX2=find(AM{2,2}==max(AM{2,2}));
                    AM2maxX2=find(AM{2,22}==max(AM{2,22}));
                    plot(t{2,2}(AM1maxX2),max(AM{2,2}),'k.');
                    plot(t{2,22}(AM2maxX2),max(AM{2,22}),'k.');


                    hold off
                    text(262,0.7,['$',num2str(round(tAM1{2,2}-tAM0{2,2},2)),'$'],'interpreter','latex','Color',pcols{1})
                    text(175,0.7,['$',num2str(round(tAM1{2,22}-tAM0{2,22},2)),'$'],'interpreter','latex','Color',pcols{2})
                    text(t{2,2}(AM1maxX2),AM{2,2}(AM1maxX2)+0.1,['$',num2str(round(max(AM{2,2}),2)),'$'],'interpreter','latex','color',pcols{1})
                    text(t{2,22}(AM2maxX2),AM{2,22}(AM2maxX2)+0.1,['$',num2str(round(max(AM{2,22}),2)),'$'],'interpreter','latex','color',pcols{2})
                    ylim([0,4.25]);
                    xlim([0,385]);
                    figprop(fig_num).xlab = xlabel('$t$ (days since March 10)');
                    figprop(fig_num).ylab = ylabel('Active Cases');
                    figprop(fig_num).leg = legend('$M_c^*/4$','$4M_c^*$','location','northeast');
                    figprop(fig_num).ax = gca;
                    
                    fig_num = fig_num + 1;
                    [fig(fig_num),figprop(fig_num)] = new_figure('active_compare_3',pos);
                    plot(t{2,16},AM{2,16},'linewidth',2,'Color',pcols{1});
                    hold on
                    plot(t{2,21},AM{2,21},'linewidth',2,'Color',pcols{2});
                    plot(t{2,11},AM{2,11},'linewidth',2,'Color',pcols{3});
                    line([tAM0{2,16},tAM1{2,16}],[0.5,0.5],'Color',pcols{1},'Linestyle','-','linewidth',2);
                    line([tAM0{2,16},tAM0{2,16}],[0.4,0.6],'Color',pcols{1},'Linestyle','-','linewidth',2);
                    line([tAM1{2,16},tAM1{2,16}],[0.4,0.6],'Color',pcols{1},'Linestyle','-','linewidth',2);
                    line([tAM0{2,21},t{2,21}(AMspots1{2,21}(1)-1)],[0.5,0.5],'Color',pcols{2},'Linestyle','-','linewidth',2);
                    line([tAM0{2,21},tAM0{2,21}],[0.4,0.6],'Color',pcols{2},'Linestyle','-','linewidth',2);
                    line([t{2,21}(AMspots1{2,21}(1)-1),t{2,22}(AMspots1{2,21}(1)-1)],[0.4,0.6],'Color',pcols{2},'Linestyle','-','linewidth',2);
                    line([tAM0{2,11},t{2,11}(AMspots1{2,11}(1)-1)],[0.5,0.5],'Color',pcols{3},'Linestyle','-','linewidth',2);
                    line([tAM0{2,11},tAM0{2,11}],[0.4,0.6],'Color',pcols{3},'Linestyle','-','linewidth',2);
                    line([t{2,11}(AMspots1{2,11}(1)-1),t{2,12}(AMspots1{2,11}(1)-1)],[0.4,0.6],'Color',pcols{3},'Linestyle','-','linewidth',2);
                    
                    AM1maxX3=find(AM{2,16}==max(AM{2,16}));
                    AM2maxX3=find(AM{2,21}==max(AM{2,21}));
                    AM3maxX3=find(AM{2,11}==max(AM{2,11}));
                    plot(t{2,16}(AM1maxX3),max(AM{2,16}),'k.');
                    plot(t{2,21}(AM2maxX3),max(AM{2,21}),'k.');
                    plot(t{2,11}(AM3maxX3),max(AM{2,11}),'k.');


                    hold off
                    text(655,0.7,['$',num2str(round(tAM1{2,16}-tAM0{2,16},2)),'$'],'interpreter','latex','Color',pcols{1})
                    text(750,0.7,['$',num2str(round(tAM1{2,21}-tAM0{2,21},2)),'$'],'interpreter','latex','Color',pcols{2})
                    text(750,0.3,['$',num2str(round(tAM1{2,11}-tAM0{2,11},2)),'$'],'interpreter','latex','Color',pcols{3})
                    text(t{2,16}(AM1maxX3)+2,AM{2,16}(AM1maxX3)+0.1,['$',num2str(round(max(AM{2,16}),2)),'$'],'interpreter','latex','color',pcols{1})
                    text(t{2,21}(AM2maxX3),AM{2,21}(AM2maxX3)+0.1,['$',num2str(round(max(AM{2,21}),2)),'$'],'interpreter','latex','color',pcols{2})
                    text(t{2,11}(AM3maxX3)-4,AM{2,11}(AM3maxX3)+0.1,['$',num2str(round(max(AM{2,11}),2)),'$'],'interpreter','latex','color',pcols{3})
                    ylim([0,4.25]);
                    xlim([600,850]);
                    figprop(fig_num).xlab = xlabel('$t$ (days since March 10)');
                    figprop(fig_num).ylab = ylabel('Active Cases');
                    figprop(fig_num).leg = legend('$M_c^*$','$2M_c^*$','$M_c^*/2$','location','northeast');
                    figprop(fig_num).ax = gca;
                    
                    
                case 2
                    M_names={'M_cd4';'M_cd2';'M_c';'2M_c';'4M_c'};
                    c_names={'$C_c^*/4$';'$C_c^*/2$';'$C_c^*$';'$2C_c^*$';'$4C_c^*$'};
                    for k=1:len_m   


                        [fig(k),figprop(k)] = new_figure(strcat('sick_case_',M_names{k}),pos);
                        temp_sick{1} = spline(t{1,1},sick{1,1},t{2,1});
                        temp_sick{2} = spline(t{3,1},sick{3,1},t{2,1});
                        err_bar_sick0 = [max([temp_sick{1} sick{2,1} temp_sick{2}],[],2)'-sick{2,1}';sick{2,1}'-min([temp_sick{1} sick{2,1} temp_sick{2}],[],2)'];
                        if err_bar_flag
                            shadedErrorBar(t{2,1},sick{2,1},err_bar_sick0,'lineprops',{'Color',pcols{end},'linewidth',2});
                        else
                            plot(t{2,1},sick{2,1},'--','linewidth',2,'Color',pcols{end});
                        end
                        legentry{1} = 'baseline';
                        hold on
                        for j=1:len_m
                            J = (k-1)*len_m+j+1;
                            temp_sick{1} = spline(t{1,J},sick{1,J},t{2,J});
                            temp_sick{2} = spline(t{3,J},sick{3,J},t{2,J});

                            err_bar_sick{j} = [max([temp_sick{1} sick{2,J} temp_sick{2}],[],2)'-sick{2,J}';sick{2,J}'-min([temp_sick{1} sick{2,J} temp_sick{2}],[],2)'];
                            if err_bar_flag
                                shadedErrorBar(t{2,J},sick{2,J},err_bar_sick{j},'lineprops',{'Color',pcols{j},'linewidth',2});
                            else
                                plot(t{2,J},sick{2,J},'linewidth',2,'Color',pcols{j});
                            end
                            legentry{j+1} = c_names{j};
                        end
                        hold off
                        ylim([0,1]);
                        xlim([0,1200]);
                        figprop(k).xlab = xlabel('$t$ (days since March 10, 2020)');
                        figprop(k).ylab = ylabel('\% sick');
                        figprop(k).leg = legend(legentry{:},'location','southeast');
                        figprop(k).ax = gca;
                        figprop(k).fnt_size=14;

                    end
                case 3
                    M_names={'M_c^*/4';'M_c^*/2';'M_c^*';'2M_c^*';'4M_c^*'};
                    c_names={'C_c^*/4';'C_c^*/2';'C_c^*';'2C_c^*';'4C_c^*'};
                    for k=1:len_q
                        [fig(k),figprop(k)] = new_figure(strcat('heat_peak_time_j_',num2str(100*q(k))),pos);
                        heatmap(c_names,M_names,reshape(peak_time(k,2:end)/peak_time(k,1),len_m,len_m)');
                        colormap(jet);
                        figprop(k).type='heat_map';
                    end
                case 4
                    pos = [2,4.5,7,4.5];
                    fig_num = 0;
                    M_names={'M_cd4';'M_cd2';'M_c';'2M_c';'4M_c'};
                    c_names={'$C_c^*/4$';'$C_c^*/2$';'$C_c^*$';'$2C_c^*$';'$4C_c^*$'};
                    for k=1:len_m   
                        fig_num = fig_num + 1;
                        [fig(fig_num),figprop(fig_num)] = new_figure(strcat('SD1_case_',M_names{k}),pos);
                        hold on
                        for j=1:len_m
                            J = (k-1)*len_m+j+1;
                            plot(t{2,J},SD1{2,J},'linewidth',2,'Color',pcols{j});
                            legentry{j+1} = c_names{j};
                        end
                        hold off
                        ylim([0,75]);
                        xlim([0,1200]);
                        figprop(fig_num).xlab = xlabel('$t$ (days since March 10, 2020)');
                        figprop(fig_num).ylab = ylabel('Social Distancing Class 1');
                        figprop(fig_num).leg = legend(legentry{:},'location','northeast');
                        figprop(fig_num).ax = gca;
                        figprop(fig_num).fnt_size=14;
                        
                        fig_num = fig_num + 1;
                        [fig(fig_num),figprop(fig_num)] = new_figure(strcat('SD2_case_',M_names{k}),pos);
                        hold on
                        for j=1:len_m
                            J = (k-1)*len_m+j+1;
                            plot(t{2,J},SD2{2,J},'linewidth',2,'Color',pcols{j});
                            legentry{j+1} = c_names{j};
                        end
                        hold off
                        ylim([0,45]);
                        xlim([0,1200]);
                        figprop(fig_num).xlab = xlabel('$t$ (days since March 10, 2020)');
                        figprop(fig_num).ylab = ylabel('Social Distancing Class 2');
                        figprop(fig_num).leg = legend(legentry{:},'location','northeast');
                        figprop(fig_num).ax = gca;
                        figprop(fig_num).fnt_size=14;

                    end
            end


        case 3
            q = linspace(0,1,100);
            len_q = length(q);
            fprintf('Loading Data...\n');
            for k=1:len_q
                file_title=strcat('qstudy-odemodel-3-q-',num2str(floor(100*q(k))),'.mat');
                DATAS = load(strcat(runpath,file_title));
                params{k,1} = DATAS.params;
                t{k,1} = DATAS.t;
                y = DATAS.y;
                S{k,1} = y(:,1);
                S1{k,1} = y(:,2);
                S2{k,1} = y(:,3);
                E{k,1} = y(:,4);
                E1{k,1} = y(:,5);
                E2{k,1} = y(:,6);
                P{k,1} = y(:,7);
                P1{k,1} = y(:,8);
                P2{k,1} = y(:,9);
                PM{k,1} = y(:,10);
                Is{k,1} = y(:,11);
                Is1{k,1} = y(:,12);
                Is2{k,1} = y(:,13);
                IsM{k,1} = y(:,14);
                Ia{k,1} = y(:,15);
                Ia1{k,1} = y(:,16);
                Ia2{k,1} = y(:,17);
                IaM{k,1} = y(:,18);
                Rs{k,1} = y(:,19);
                Rs1{k,1} = y(:,20);
                Rs2{k,1} = y(:,21);
                RsM{k,1} = y(:,22);
                Ra{k,1} = y(:,23);
                Ra1{k,1} = y(:,24);
                Ra2{k,1} = y(:,25);
                RaM{k,1} = y(:,26);
                M{k,1} = y(:,27);
                C{k,1} = y(:,28);
            end
            fprintf('Data Loaded...\n');

            %Data processing
            peak_val = 0;
            It = {};
            for k=1:len_q
                It{k,1} = Is{k,1}+Is1{k,1}+Is2{k,1}+IsM{k,1};
                peak_val(k) = max(It{k,1});
            end

            [fig,figprop] = new_figure('q_study',pos);
            plot(q,peak_val,'Color',pcols{1},'linewidth',2);
            figprop.xlab = xlabel('$q$');
            figprop.ylab = ylabel('max($I_T$)');
        case 4
            load('prcc_Model_LHS_10000_Aug2120.mat');

            prclab={'sickS','susceptible','peakval','peaktime','sickA'};
            prctitles={'Cumulative Infected (Symptomatic)','Susceptibles','Outbreak Peak Value','Outbreak Peak Time','Cumulative Infected (Asymptomatic)'};
            PRCC_var={'$\alpha$', '$\sigma$', '$\phi$', ...
                        '$\gamma$','$q$', '$q_0$','$q_2$', '$\mu_I$','$q_I$',...
                        '$\rho_A$', '$\rho_S$'};
            for k=1:length(prcc)
                [fig(k),figprop(k)] = new_figure(['prcc_',prclab{k}],pos);
                b=barh(prcc{k}(end,:),'FaceColor','flat');
                b.CData = kron(ones(length(prcc{k}(end,:)),1),pcols{end});
                b.CData(sign_label{k}.index{end},:)=kron(ones(length(sign_label{k}.index{end}),1),pcols{1});
                b.CData(sign_label{k}.index{end}(abs(str2num(sign_label{k}.value{end}))>0.5),:)=kron(ones(sum(abs(str2num(sign_label{k}.value{end}))>0.5),1),pcols{2});
                hold on
                barh(0,'FaceColor',pcols{2});
                barh(0,'FaceColor',pcols{end});
                hold off
                yticklabels(PRCC_var);
                xlim([-1,1]);
                figprop(k).xlab=xlabel('PRCC');
                figprop(k).leg = legend({strcat('$p<0.05$,', 10, '$\rm{PRCC}<0.5$'),strcat('$p<0.05$,',10,'$\rm{PRCC}>0.5$'),'$p>0.05$'},'location','best');
                figprop(k).ax = gca;
                figprop(k).title=title(prctitles{k});
                figprop(k).fnt_size=14;

            end
        case 5
            rho_multiplier = [1/2;1;2];
            rho_label = {'0p5','1','2'};
            multiplier = [1/4;1/2;1;2;4];
            len_rho = length(rho_multiplier);
            len_m = length(multiplier);
            fprintf('Loading Data...\n');
            for k=1:len_rho
                file_title=strcat('odemodel-3-rho_baseline.mat');
                DATAS = load(strcat(runpath,file_title));
                params{k,1} = DATAS.params;
                t{k,1} = DATAS.t;
                y = DATAS.y;
                S{k,1} = y(:,1);
                S1{k,1} = y(:,2);
                S2{k,1} = y(:,3);
                E{k,1} = y(:,4);
                E1{k,1} = y(:,5);
                E2{k,1} = y(:,6);
                P{k,1} = y(:,7);
                P1{k,1} = y(:,8);
                P2{k,1} = y(:,9);
                PM{k,1} = y(:,10);
                Is{k,1} = y(:,11);
                Is1{k,1} = y(:,12);
                Is2{k,1} = y(:,13);
                IsM{k,1} = y(:,14);
                Ia{k,1} = y(:,15);
                Ia1{k,1} = y(:,16);
                Ia2{k,1} = y(:,17);
                IaM{k,1} = y(:,18);
                Rs{k,1} = y(:,19);
                Rs1{k,1} = y(:,20);
                Rs2{k,1} = y(:,21);
                RsM{k,1} = y(:,22);
                Ra{k,1} = y(:,23);
                Ra1{k,1} = y(:,24);
                Ra2{k,1} = y(:,25);
                RaM{k,1} = y(:,26);
                M{k,1} = y(:,27);
                C{k,1} = y(:,28);
                for j1=1:len_m
                    for j2=1:len_m
                        K2 = 1 + (j1-1)*len_m + j2;
                        file_title=strcat('odemodel-3-rho_mult-',rho_label{k},'-case',num2str((j1-1)*len_m+j2),'.mat');
                        DATAS = load(strcat(runpath,file_title));
                        params{k,K2} = DATAS.params;
                        t{k,K2} = DATAS.t;
                        y = DATAS.y;
                        S{k,K2} = y(:,1);
                        S1{k,K2} = y(:,2);
                        S2{k,K2} = y(:,3);
                        E{k,K2} = y(:,4);
                        E1{k,K2} = y(:,5);
                        E2{k,K2} = y(:,6);
                        P{k,K2} = y(:,7);
                        P1{k,K2} = y(:,8);
                        P2{k,K2} = y(:,9);
                        PM{k,K2} = y(:,10);
                        Is{k,K2} = y(:,11);
                        Is1{k,K2} = y(:,12);
                        Is2{k,K2} = y(:,13);
                        IsM{k,K2} = y(:,14);
                        Ia{k,K2} = y(:,15);
                        Ia1{k,K2} = y(:,16);
                        Ia2{k,K2} = y(:,17);
                        IaM{k,K2} = y(:,18);
                        Rs{k,K2} = y(:,19);
                        Rs1{k,K2} = y(:,20);
                        Rs2{k,K2} = y(:,21);
                        RsM{k,K2} = y(:,22);
                        Ra{k,K2} = y(:,23);
                        Ra1{k,K2} = y(:,24);
                        Ra2{k,K2} = y(:,25);
                        RaM{k,K2} = y(:,26);
                        M{k,K2} = y(:,27);
                        C{k,K2} = y(:,28);
                    end
                end
            end
            fprintf('Data Loaded...\n');

%%%%%%%%%%%%%%%% Now do some processing on the data
            if process_data_5 == 0
                n_crit = 1/2; %Value for which we surpass health resources
                for j=1:len_rho
                    for k=1:len_m^2+1
                        It{j,k} = Is{j,k}+Is1{j,k}+Is2{j,k}+IsM{j,k};
                        peak_time(j,k) = t{j,k}(It{j,k}==max(It{j,k}));
                        peak_val(j,k) = max(It{j,k});
                        sickS{j,k} = (Is{j,k}+Is1{j,k}+Is2{j,k}+IsM{j,k}+Rs{j,k}+Rs1{j,k}+Rs2{j,k}+RsM{j,k})*params{j,k}.N0/params{j,k}.N;
                        sickA{j,k} = (Ia{j,k}+Ia1{j,k}+Ia2{j,k}+IaM{j,k}+Ra{j,k}+Ra1{j,k}+Ra2{j,k}+RaM{j,k})*params{j,k}.N0/params{j,k}.N;
                        sick{j,k} = (sickS{j,k} + sickA{j,k});

                        KM{j,k} = 1/log(2)*max((params{j,k}.rho0*(Ia{j,k}+Ia1{j,k}+Ia2{j,k}+P{j,k}+P1{j,k}+P2{j,k}) + params{j,k}.rhoI*(Is{j,k}+Is1{j,k}+Is2{j,k}))./M{j,k},0);
                        AM{j,k} = PM{j,k} + IsM{j,k} + IaM{j,k};
                        Kfun{j,k} = max(KM{j,k}-params{j,k}.Kc,0)./(max(KM{j,k}-params{j,k}.Kc,0)+params{j,k}.K0-params{j,k}.Kc);
                        Afun{j,k} = max(AM{j,k}-params{j,k}.Mc,0)./(max(AM{j,k}-params{j,k}.Mc,0)+params{j,k}.M0-params{j,k}.Mc);
                        mu{j,k} = params{j,k}.mumax*Kfun{j,k}.*Afun{j,k};
                        nu{j,k} = params{j,k}.numax*max(C{j,k}-params{j,k}.Cc,0)./(max(C{j,k}-params{j,k}.Cc,0)+params{j,k}.C0-params{j,k}.Cc);

                        %Find spike times for infection
                        donespikes = 0;
                        Itcount(j,k) = 0;
                        sgnIt = 1;
                        while(~donespikes)
                            sgnIt = sgnIt-1+find(sign(It{j,k}(sgnIt:end)-n_crit)==1,1);
                            if isempty(sgnIt)
                                donespikes=1;
                            else
                                Itcount(j,k) = Itcount(j,k)+1;
                                Itspots0{j,k}(Itcount(j,k)) = sgnIt;
                                tIt0{j,k}(Itcount(j,k))= t{j,k}(sgnIt);
                                sgnIt = sgnIt-1+find(sign(It{j,k}(sgnIt:end)-n_crit)==-1,1);

                                if isempty(sgnIt)
                                    Itspots1{j,k}(Itcount(j,k)) = length(It{j,k});
                                    tIt1{j,k}(Itcount(j,k)) = t{j,k}(end);
                                    t_cut_flag(j,k) = 1;
                                else
                                    Itspots1{j,k}(Itcount(j,k)) = sgnIt;
                                    tIt1{j,k}(Itcount(j,k)) = t{j,k}(sgnIt);
                                    t_cut_flag(j,k) = 0;
                                end

                            end
                        end
                        donespikes = 0;
                        AMcount(j,k) = 0;
                        sgnAM = 1;
                        while(~donespikes)
                            sgnAM = sgnAM-1+find(sign(AM{j,k}(sgnAM:end)-n_crit)==1,1);
                            if isempty(sgnAM)
                                donespikes=1;
                            else
                                AMcount(j,k) = AMcount(j,k)+1;
                                AMspots0{j,k}(AMcount(j,k)) = sgnAM;
                                tAM0{j,k}(AMcount(j,k))= t{j,k}(sgnAM);
                                sgnAM = sgnAM-1+find(sign(AM{j,k}(sgnAM:end)-n_crit)==-1,1);

                                if isempty(sgnAM)
                                    AMspots1{j,k}(AMcount(j,k)) = length(AM{j,k});
                                    tAM1{j,k}(AMcount(j,k)) = t{j,k}(end);
                                    t_AMcut_flag(j,k) = 1;
                                else
                                    AMspots1{j,k}(AMcount(j,k)) = sgnAM;
                                    tAM1{j,k}(AMcount(j,k)) = t{j,k}(sgnAM);
                                    t_AMcut_flag(j,k) = 0;
                                end

                            end
                        end
                        health_cost(j,k) = 0;
                        if AMcount(j,k)~=0
                            for k2=1:length(tAM0{j,k})
                                health_cost(j,k) = health_cost(j,k) + trapz(t{j,k}(AMspots0{j,k}(k2):AMspots1{j,k}(k2)),AM{j,k}(AMspots0{j,k}(k2):AMspots1{j,k}(k2)));
                            end
                        end
                        rel_health_cost(j,k) = health_cost(j,k)/health_cost(j,1);
                        cost(j,k) = C{j,k}(end);
                    end
                end
                max_cost = max(max(cost));
                rel_cost = cost/max_cost;
                total_cost = 1/2*rel_health_cost+1/2*rel_cost;
                process_data_5 = 1;
            end
%%%%%%%%%%%%DONE PROCESSING DATA

            %sub_experiment is instead of all plots, go through the ones we
            %want
            % 0 - It graphs plotted with a critical threshold
            % 1 - heatmap graphs of costs
            % 2 - sick people graphs
            % 3 - peak sick time heat map

            switch sub_experiment

                case 0
                    pos = [2,4.5,7,4.5];
                    for k=2:(len_m^2+1)
                        [fig(k-1),figprop(k-1)] = new_figure(strcat('rho-I_t_case_',num2str(k-1)),pos);
                        temp_It{1} = spline(t{1,k},It{1,k},t{2,k});
                        temp_It{2} = spline(t{3,k},It{3,k},t{2,k});

                        err_bar{k-1} = [max([temp_It{1} It{2,k} temp_It{2}],[],2)'-It{2,k}';It{2,k}'];
                        
                        temp_AM{1} = spline(t{1,k},AM{1,k},t{2,k});
                        temp_AM{2} = spline(t{3,k},AM{3,k},t{2,k});
                        
                        err_barAM{k-1} = [max([temp_AM{1} AM{2,k} temp_AM{2}],[],2)'-AM{2,k}';AM{2,k}'];

                        yyaxis left
                        shadedErrorBar(t{2,k}*12/365,AM{2,k},err_barAM{k-1},'lineprops',{'Color',pcols{3},'linewidth',2});
                        hold on
                        shadedErrorBar(t{2,k}*12/365,It{2,k},err_bar{k-1},'lineprops',{'Color',pcols{1},'linewidth',2});
                        plot(t{k}*12/365,n_crit*ones(length(t{k}),1),'k--','linewidth',2);
                        ylim([0,16]);
                        figprop(k-1).ylab = ylabel('Cases');
                        figprop(k-1).ax = gca;
    %                     hold off
                        yyaxis right

                        temp_C{1} = spline(t{1,k},C{1,k},t{2,k});
                        temp_C{2} = spline(t{3,k},C{3,k},t{2,k});

                        err_barC{k-1} = [max([temp_C{1} C{2,k} temp_C{2}],[],2)'-C{2,k}';C{2,k}'-min([temp_C{1} C{2,k} temp_C{2}],[],2)'];

                        shadedErrorBar(t{2,k}*12/365,C{2,k},err_barC{k-1},'lineprops',{'Color',pcols{2},'linewidth',2});
                        ylim([0,250]);

                        figprop(k-1).ylab2 = ylabel('$C$ (days)');

                        figprop(k-1).xlab = xlabel('$t$ (months)');
                    end
                case 1
                    M_names={'M_c^*/4';'M_c^*/2';'M_c^*';'2M_c^*';'4M_c^*'};
                    c_names={'C_c^*/4';'C_c^*/2';'C_c^*';'2C_c^*';'4C_c^*'};
                    row_names={'k';'c';'health_cost';'cost';'total_cost'};
                    for k=1:len_rho
                        for k1=1:len_m
                            heat_health_data{k}(k1,:)=rel_health_cost(k,(k1-1)*len_m+2:(k1-1)*len_m+6);
                            heat_cost_data{k}(k1,:)=rel_cost(k,(k1-1)*len_m+2:(k1-1)*len_m+6);
                            heat_tot_data{k}(k1,:)=total_cost(k,(k1-1)*len_m+2:(k1-1)*len_m+6);

                        end
                        [fig((k-1)*3+1),figprop((k-1)*3+1)] = new_figure(strcat('rho-heat_health_mult_',rho_label{k}),pos);
                        heatmap(c_names,M_names,heat_health_data{k});
                        colormap(jet);
                        figprop((k-1)*3+1).type='heat_map';
                        [fig((k-1)*3+2),figprop((k-1)*3+2)] = new_figure(strcat('rho-heat_cost_mult_',rho_label{k}),pos);
                        heatmap(c_names,M_names,heat_cost_data{k});
                        colormap(jet);
                        figprop((k-1)*3+2).type='heat_map';
                        [fig((k-1)*3+3),figprop((k-1)*3+3)] = new_figure(strcat('rho-heat_total_mult_',rho_label{k}),pos);
                        heatmap(c_names,M_names,heat_tot_data{k});
                        colormap(jet);
                        figprop((k-1)*3+3).type='heat_map';
                    end
                case 2
                    M_names={'M_cd4';'M_cd2';'M_c';'2M_c';'4M_c'};
                    c_names={'$C_c^*/4$';'$C_c^*/2$';'$C_c^*$';'$2C_c^*$';'$4C_c^*$'};
                    for k=1:len_m   
                        [fig(k),figprop(k)] = new_figure(strcat('rho-sick_case_',M_names{k}),pos);
                        plot(t{2,1}*12/365,sickS{2,1},'Color',pcols{end},'linewidth',2);
                        legentry{1} = 'baseline';
                        hold on
                        for j=1:len_m
                            J = (k-1)*len_m+j+1;
                            temp_sick{1} = spline(t{1,J},sickS{1,J},t{2,J});
                            temp_sick{2} = spline(t{3,J},sickS{3,J},t{2,J});

                            err_bar_sick{j} = [max([temp_sick{1} sickS{2,J} temp_sick{2}],[],2)'-sickS{2,J}';sickS{2,J}'-min([temp_sick{1} sickS{2,J} temp_sick{2}],[],2)'];
                            shadedErrorBar(t{2,J}*12/365,sickS{2,J},err_bar_sick{j},'lineprops',{'Color',pcols{j},'linewidth',2});
                            legentry{j+1} = c_names{j};
                        end
                        hold off
                        ylim([0,1]);
                        figprop(k).xlab = xlabel('$t$ (months)');
                        figprop(k).ylab = ylabel('\% sick');
                        figprop(k).leg = legend(legentry{:},'location','northwest');
                        figprop(k).ax = gca;

                    end
                case 3
                    M_names={'M_c^*/4';'M_c^*/2';'M_c^*';'2M_c^*';'4M_c^*'};
                    c_names={'C_c^*/4';'C_c^*/2';'C_c^*';'2C_c^*';'4C_c^*'};
                    for k=1:len_rho
                        [fig(k),figprop(k)] = new_figure(strcat('rho-heat_peak_time_mult_',rho_label{k}),pos);
                        heatmap(c_names,M_names,reshape(peak_time(k,2:end)/peak_time(k,1),len_m,len_m)');
                        colormap(jet);
                        figprop(k).type='heat_map';
                    end
            end
            
        case 6
            DATA_sim = load(strcat(runpath,'data_fit_case.mat'));
            DATA_data = load('DATA-Aug18.mat');
            DATA_projectdat = load('DATA-Jan06.mat');
            
            DATA_mob = load('retail-data-Jan621.mat');
            
            DATA_altsim = load(strcat(runpath,'alt_fit.mat'));
            
            y = DATA_sim.y;
            t = DATA_sim.t;
            params = DATA_sim.params;
            
            S = y(:,1);
            S1 = y(:,2);
            S2 = y(:,3);
            E = y(:,4);
            E1 = y(:,5);
            E2 = y(:,6);
            P = y(:,7);
            P1 = y(:,8);
            P2 = y(:,9);
            PM = y(:,10);
            Is = y(:,11);
            Is1 = y(:,12);
            Is2 = y(:,13);
            IsM = y(:,14);
            Ia = y(:,15);
            Ia1 = y(:,16);
            Ia2 = y(:,17);
            IaM = y(:,18);
            M = y(:,27);
            C = y(:,28);
            
            AM = PM + IsM + IaM;
            
            SD = S1+S2+E1+E2+P1+P2+PM+Is1+Is2+IsM+Ia1+Ia2+IaM;
            
            y2 = DATA_altsim.y;
            t2 = DATA_altsim.t;
            params2 = DATA_altsim.params;
            
            S2 = y2(:,1);
            S12 = y2(:,2);
            S22 = y2(:,3);
            E2 = y2(:,4);
            E12 = y2(:,5);
            E22 = y2(:,6);
            P2 = y2(:,7);
            P12 = y2(:,8);
            P22 = y2(:,9);
            PM2 = y2(:,10);
            Is2 = y2(:,11);
            Is12 = y2(:,12);
            Is22 = y2(:,13);
            IsM2 = y2(:,14);
            Ia2 = y2(:,15);
            Ia12 = y2(:,16);
            Ia22 = y2(:,17);
            IaM2 = y2(:,18);
            M2 = y2(:,27);
            C2 = y2(:,28);
            
            AM2 = PM2 + IsM2 + IaM2;
            
            t_spot = find(abs(t-DATA_data.DATA_T(end))==min(abs(t-DATA_data.DATA_T(end))));
            
            max_dat_spot = find(DATA_data.DATA_pos==max(DATA_data.DATA_pos));
            max_sim_spot = find(AM(1:t_spot)==max(AM(1:t_spot)));
            
            pos_dat_max = DATA_data.DATA_pos(max_dat_spot);
            pos_sim_max = AM(max_sim_spot)*params.N0;
            
            t_dat_max = DATA_data.DATA_T(max_dat_spot);
            t_sim_max = t(max_sim_spot);
            
            diff_pos = abs(pos_dat_max-pos_sim_max);
            diff_t = abs(t_dat_max-t_sim_max);
            
            
            
            [fig(1),figprop(1)] = new_figure('AM_data_compare',pos);
            
            plot(DATA_data.DATA_T,DATA_data.DATA_pos,'o','Color',pcols{2});
            hold on
            plot(t(1:t_spot),AM(1:t_spot)*params.N0,'linewidth',2,'Color',pcols{1});
            hold off
            figprop(1).xlab = xlabel('$t$ (days since Mar 10, 2020)');
            figprop(1).ylab = ylabel('Active Cases');
            figprop(1).leg = legend('Data','Simulation','location','northeast');
            
            
            [fig(2),figprop(2)] = new_figure('tot_data_compare',pos);
            
            plot(DATA_data.DATA_T,DATA_data.DATA_tot,'o','Color',pcols{2});
            hold on
            plot(t(1:t_spot),M(1:t_spot)*params.N0,'linewidth',2,'Color',pcols{1});
            hold off
            figprop(2).xlab = xlabel('$t$ (days since Mar 10, 2020)');
            figprop(2).ylab = ylabel('Total Cases');
            figprop(2).leg = legend('Data','Simulation','location','northwest');
            
            [fig(3),figprop(3)] = new_figure('AM_data_project',pos);
            
            plot(DATA_data.DATA_T,DATA_data.DATA_pos,'o','Color',pcols{2});
            hold on
            plot(DATA_projectdat.DATA_T(163:end),DATA_projectdat.DATA_pos(163:end),'o','Color',pcols{end});
            plot(t,AM*params.N0,'linewidth',2,'Color',pcols{1});
            plot(t,1/2*params.N0*ones(length(t),1),'k--','linewidth',2);
            hold off
            xlim([0,DATA_projectdat.DATA_T(end)]);
            ylim([0,3E4]);
            figprop(3).xlab = xlabel('$t$ (days since Mar 10, 2020)');
            figprop(3).ylab = ylabel('Active Cases');
            figprop(3).leg = legend('Fitted Data','Unfitted Data','Simulation','location','northwest');
            
            [fig(4),figprop(4)] = new_figure('tot_data_project',pos);
            
            plot(DATA_data.DATA_T,DATA_data.DATA_tot,'o','Color',pcols{2});
            hold on
            plot(DATA_projectdat.DATA_T(163:end),DATA_projectdat.DATA_tot(163:end),'o','Color',pcols{end});
            plot(t,M*params.N0,'linewidth',2,'Color',pcols{1});
            hold off
            xlim([0,DATA_projectdat.DATA_T(end)]);
            ylim([0,2.2E5]);
            figprop(4).xlab = xlabel('$t$ (days since Mar 10, 2020)');
            figprop(4).ylab = ylabel('Total Cases');
            figprop(4).leg = legend('Fitted Data','Unfitted Data','Simulation','location','northwest');
            
            [fig(5),figprop(5)] = new_figure('mobility_compare',pos);
            
            plot(DATA_mob.retail_data_t,DATA_mob.retail_data_dat,'o','Color',pcols{2});
            hold on
            plot(t,-SD*params.N0/params.N*100,'linewidth',2,'Color',pcols{1});
            hold off
            xlim([0,DATA_mob.retail_data_t(end)]);
            figprop(5).xlab = xlabel('$t$ (days since Mar 10, 2020)');
            figprop(5).ylab = ylabel('\% change');
            figprop(5).leg = legend('Mobility Data','Simulation','location','best');
            
            [fig(6),figprop(6)] = new_figure('alt_behaviour_active',pos);
            
            plot(DATA_data.DATA_T,DATA_data.DATA_pos,'o','Color',pcols{2});
            hold on
            plot(DATA_projectdat.DATA_T(163:220),DATA_projectdat.DATA_pos(163:220),'o','Color',pcols{3});
            plot(DATA_projectdat.DATA_T(221:end),DATA_projectdat.DATA_pos(221:end),'o','Color',pcols{end});
            plot(t2,AM2*params2.N0,'linewidth',2,'Color',pcols{1});
            hold off
            xlim([0,DATA_projectdat.DATA_T(end)]);
            ylim([0,3E4]);
            figprop(3).xlab = xlabel('$t$ (days since Mar 10, 2020)');
            figprop(3).ylab = ylabel('Active Cases');
            figprop(3).leg = legend('Original Fitted Data','New Fitted Data','Unfitted Data','Simulation','location','northwest');
            
            
            
            [fig(7),figprop(7)] = new_figure('alt_behaviour_total',pos);
            
             plot(DATA_data.DATA_T,DATA_data.DATA_tot,'o','Color',pcols{2});
            hold on
            plot(DATA_projectdat.DATA_T(163:220),DATA_projectdat.DATA_tot(163:220),'o','Color',pcols{3});
            plot(DATA_projectdat.DATA_T(221:end),DATA_projectdat.DATA_tot(221:end),'o','Color',pcols{end});
            plot(t2,M2*params2.N0,'linewidth',2,'Color',pcols{1});
            hold off
            xlim([0,DATA_projectdat.DATA_T(end)]);
            ylim([0,2.2E5]);
            figprop(4).xlab = xlabel('$t$ (days since Mar 10, 2020)');
            figprop(4).ylab = ylabel('Total Cases');
            figprop(4).leg = legend('Original Fitted Data','New Fitted Data','Unfitted Data','Simulation','location','northwest');
        case 7
            files = {'case-eta1d2.mat';'case-eta1d5.mat'};
            titles = {'eta1d2';'eta1d5'};
            for k=1:2
                DATAS = load([runpath,files{k}]);
                params{k} = DATAS.params;
                t{k} = DATAS.t*365/12;
                y = DATAS.y;
                S{k} = y(:,1);
                S1{k} = y(:,2);
                S2{k} = y(:,3);
                E{k} = y(:,4);
                E1{k} = y(:,5);
                E2{k} = y(:,6);
                P{k} = y(:,7);
                P1{k} = y(:,8);
                P2{k} = y(:,9);
                PM{k} = y(:,10);
                Is{k} = y(:,11);
                Is1{k} = y(:,12);
                Is2{k} = y(:,13);
                IsM{k} = y(:,14);
                Ia{k} = y(:,15);
                Ia1{k} = y(:,16);
                Ia2{k} = y(:,17);
                IaM{k} = y(:,18);
                Rs{k} = y(:,19);
                Rs1{k} = y(:,20);
                Rs2{k} = y(:,21);
                RsM{k} = y(:,22);
                Ra{k} = y(:,23);
                Ra1{k} = y(:,24);
                Ra2{k} = y(:,25);
                RaM{k} = y(:,26);
                M{k} = y(:,27);
                C{k} = y(:,28);
                
                It{k} = Is{k}+Is1{k}+Is2{k}+IsM{k};
                IAt{k} = Ia{k}+Ia1{k}+Ia2{k}+IaM{k};
                Pt{k} = P{k}+P1{k}+P2{k}+PM{k};
                AM{k} = PM{k}+IsM{k}+IaM{k};
                
                [fig(k),figprop(k)]=new_figure(titles{k},pos);
                yyaxis left
                plot(t{k},(It{k}+IAt{k}+Pt{k})*params{k}.N0,'linewidth',2,'Color',pcols{1},'Linestyle','--');
                hold on
                plot(t{k},AM{k}*params{k}.N0,'linewidth',2,'Color',pcols{3},'Linestyle','-');
                plot(t{k},1/2*ones(length(t{k}),1)*params{k}.N0,'k--','linewidth',2);
%                 plot(t{k},ones(length(t{k}),1)*params{k}.N0,'--','linewidth',2,'Color',pcols{end});
                hold off
                figprop(k).ylab = ylabel('Active Cases');
                ylim([0,35E4]);
                set(gca,'ycolor','k');
                
                yyaxis right
                plot(t{k},C{k},'linewidth',2,'Color',pcols{2});
                ylim([0,600]);
                
                xlim([0,7*365]);
                figprop(k).xlab = xlabel('$t$ (days since March 10, 2020)');
                figprop(k).ylab2 = ylabel('$C$ (days)');
                figprop(k).leg = legend({'Total Active Cases';'Tested Active Cases';'$N_{\rm crit}/2$';'Relaxation Cost'},'location','northwest');
                figprop(k).ax = gca;
                figprop(k).fnt_size = 14;
                set(gca,'ycolor',pcols{2});
                
            end
            
            
        case 8
            multiplier = [1/16;1/8;1/4;1/2;1];
            len_m = length(multiplier);
            fprintf('Loading Data...\n');
            file_title=strcat('odemodel-3-eta-case0.mat');
            DATAS = load(strcat(runpath,file_title));
            params{1,1} = DATAS.params;
            t{1,1} = DATAS.t;
            y = DATAS.y;
            S{1,1} = y(:,1);
            S1{1,1} = y(:,2);
            S2{1,1} = y(:,3);
            E{1,1} = y(:,4);
            E1{1,1} = y(:,5);
            E2{1,1} = y(:,6);
            P{1,1} = y(:,7);
            P1{1,1} = y(:,8);
            P2{1,1} = y(:,9);
            PM{1,1} = y(:,10);
            Is{1,1} = y(:,11);
            Is1{1,1} = y(:,12);
            Is2{1,1} = y(:,13);
            IsM{1,1} = y(:,14);
            Ia{1,1} = y(:,15);
            Ia1{1,1} = y(:,16);
            Ia2{1,1} = y(:,17);
            IaM{1,1} = y(:,18);
            Rs{1,1} = y(:,19);
            Rs1{1,1} = y(:,20);
            Rs2{1,1} = y(:,21);
            RsM{1,1} = y(:,22);
            Ra{1,1} = y(:,23);
            Ra1{1,1} = y(:,24);
            Ra2{1,1} = y(:,25);
            RaM{1,1} = y(:,26);
            M{1,1} = y(:,27);
            C{1,1} = y(:,28);
            for j1=1:len_m
                for j2=1:len_m
                    K2 = 1 + (j1-1)*len_m + j2;
                    file_title=strcat('odemodel-3-eta-case',num2str((j1-1)*len_m+j2),'.mat');
                    DATAS = load(strcat(runpath,file_title));
                    params{1,K2} = DATAS.params;
                    t{1,K2} = DATAS.t;
                    y = DATAS.y;
                    S{1,K2} = y(:,1);
                    S1{1,K2} = y(:,2);
                    S2{1,K2} = y(:,3);
                    E{1,K2} = y(:,4);
                    E1{1,K2} = y(:,5);
                    E2{1,K2} = y(:,6);
                    P{1,K2} = y(:,7);
                    P1{1,K2} = y(:,8);
                    P2{1,K2} = y(:,9);
                    PM{1,K2} = y(:,10);
                    Is{1,K2} = y(:,11);
                    Is1{1,K2} = y(:,12);
                    Is2{1,K2} = y(:,13);
                    IsM{1,K2} = y(:,14);
                    Ia{1,K2} = y(:,15);
                    Ia1{1,K2} = y(:,16);
                    Ia2{1,K2} = y(:,17);
                    IaM{1,K2} = y(:,18);
                    Rs{1,K2} = y(:,19);
                    Rs1{1,K2} = y(:,20);
                    Rs2{1,K2} = y(:,21);
                    RsM{1,K2} = y(:,22);
                    Ra{1,K2} = y(:,23);
                    Ra1{1,K2} = y(:,24);
                    Ra2{1,K2} = y(:,25);
                    RaM{1,K2} = y(:,26);
                    M{1,K2} = y(:,27);
                    C{1,K2} = y(:,28);
                end
            end
            fprintf('Data Loaded...\n');

%%%%%%%%%%%%% Now do some processing on the data
            omega = 0.9; % decide on weighting
            if process_data_8 == 0 %If looping only process the data once
                n_crit = 1/2; %Value for which we surpass health resources
                j = 1;
                for k=1:len_m^2+1
                    It{j,k} = Is{j,k}+Is1{j,k}+Is2{j,k}+IsM{j,k};
                    IAt{j,k} = Ia{j,k}+Ia1{j,k}+Ia2{j,k}+IaM{j,k};
                    Pt{j,k} = P{j,k}+P1{j,k}+P2{j,k}+PM{j,k};
                    SD1{j,k} = S1{j,k}+E1{j,k}+P1{j,k}+Is1{j,k}+Ia1{j,k};
                    SD2{j,k} = S2{j,k}+E2{j,k}+P2{j,k}+PM{j,k}+Is2{j,k}+IsM{j,k}+Ia2{j,k}+IaM{j,k};
                    peak_time(j,k) = t{j,k}(It{j,k}==max(It{j,k}));
                    peak_val(j,k) = max(It{j,k});
                    sickS{j,k} = (Is{j,k}+Is1{j,k}+Is2{j,k}+IsM{j,k}+Rs{j,k}+Rs1{j,k}+Rs2{j,k}+RsM{j,k})*params{j,k}.N0/params{j,k}.N;
                    sickA{j,k} = (Ia{j,k}+Ia1{j,k}+Ia2{j,k}+IaM{j,k}+Ra{j,k}+Ra1{j,k}+Ra2{j,k}+RaM{j,k})*params{j,k}.N0/params{j,k}.N;
                    sick{j,k} = (sickS{j,k} + sickA{j,k});

                    KM{j,k} = 1/log(2)*max((params{j,k}.rho0*(Ia{j,k}+Ia1{j,k}+Ia2{j,k}+P{j,k}+P1{j,k}+P2{j,k}) + params{j,k}.rhoI*(Is{j,k}+Is1{j,k}+Is2{j,k}))./M{j,k},0);
                    AM{j,k} = PM{j,k} + IsM{j,k} + IaM{j,k};
                    Kfun{j,k} = max(KM{j,k}-params{j,k}.Kc,0)./(max(KM{j,k}-params{j,k}.Kc,0)+params{j,k}.K0-params{j,k}.Kc);
                    Afun{j,k} = max(AM{j,k}-params{j,k}.Mc,0)./(max(AM{j,k}-params{j,k}.Mc,0)+params{j,k}.M0-params{j,k}.Mc);
                    mu{j,k} = params{j,k}.mumax*Kfun{j,k}.*Afun{j,k};
                    nu{j,k} = params{j,k}.numax*max(C{j,k}-params{j,k}.Cc,0)./(max(C{j,k}-params{j,k}.Cc,0)+params{j,k}.C0-params{j,k}.Cc).*max(params{j,k}.eta*params{j,k}.Mc-AM{j,k},0);

                    %Find spike times for infection
                    donespikes = 0;
                    Itcount(j,k) = 0;
                    sgnIt = 1;
                    while(~donespikes)
                        sgnIt = sgnIt-1+find(sign(It{j,k}(sgnIt:end)-n_crit)==1,1);
                        if isempty(sgnIt)
                            donespikes=1;
                        else
                            Itcount(j,k) = Itcount(j,k)+1;
                            Itspots0{j,k}(Itcount(j,k)) = sgnIt;
                            tIt0{j,k}(Itcount(j,k))= t{j,k}(sgnIt);
                            sgnIt = sgnIt-1+find(sign(It{j,k}(sgnIt:end)-n_crit)==-1,1);

                            if isempty(sgnIt)
                                Itspots1{j,k}(Itcount(j,k)) = length(It{j,k});
                                tIt1{j,k}(Itcount(j,k)) = t{j,k}(end);
                                t_cut_flag(j,k) = 1;
                            else
                                Itspots1{j,k}(Itcount(j,k)) = sgnIt;
                                tIt1{j,k}(Itcount(j,k)) = t{j,k}(sgnIt);
                                t_cut_flag(j,k) = 0;
                            end

                        end
                    end
                    donespikes = 0;
                    AMcount(j,k) = 0;
                    sgnAM = 1;
                    while(~donespikes)
                        sgnAM = sgnAM-1+find(sign(AM{j,k}(sgnAM:end)-n_crit)==1,1);
                        if isempty(sgnAM)
                            donespikes=1;
                        else
                            AMcount(j,k) = AMcount(j,k)+1;
                            AMspots0{j,k}(AMcount(j,k)) = sgnAM;
                            tAM0{j,k}(AMcount(j,k))= t{j,k}(sgnAM);
                            sgnAM = sgnAM-1+find(sign(AM{j,k}(sgnAM:end)-n_crit)==-1,1);

                            if isempty(sgnAM)
                                AMspots1{j,k}(AMcount(j,k)) = length(AM{j,k});
                                tAM1{j,k}(AMcount(j,k)) = t{j,k}(end);
                                t_AMcut_flag(j,k) = 1;
                            else
                                AMspots1{j,k}(AMcount(j,k)) = sgnAM;
                                tAM1{j,k}(AMcount(j,k)) = t{j,k}(sgnAM);
                                t_AMcut_flag(j,k) = 0;
                            end

                        end
                    end

                    health_cost(j,k) = 0;
                    if AMcount(j,k)~=0
                        for k2=1:length(tAM0{j,k})
                            health_cost(j,k) = health_cost(j,k) + trapz(t{j,k}(AMspots0{j,k}(k2):AMspots1{j,k}(k2)),AM{j,k}(AMspots0{j,k}(k2):AMspots1{j,k}(k2)));
                        end
                    end
                    rel_health_cost(j,k) = health_cost(j,k)/health_cost(j,1);
                    cost(j,k) = C{j,k}(end);
                end
                max_cost(j) = max(cost(j,:));
                rel_cost(j,:) = cost(j,:)/max_cost(j);
                total_cost = omega*rel_health_cost+(1-omega)*rel_cost;
                for k1=2:len_m^2+1
                    for k2=k1:len_m^2+1
                        rel_total_cost(k1-1,k2-1) = total_cost(1,k2)/total_cost(1,k1);
                    end
                end
                process_data_8 = 1;
            end
%%%%%%%%%%%DONE PROCESSING

            %sub_experiment is instead of all plots, go through the ones we
            %want
            % 0 - It graphs plotted with a critical threshold
            % 1 - heatmap graphs of costs
            % 2 - sick people graphs
            % 3 - peak sick time heat map
            % 4 - social distancing plots
            

            switch sub_experiment

                case 0
                    pos = [2,4.5,7,4.5];
                    for k=1:(len_m^2+1)
                        [fig(k),figprop(k)] = new_figure(strcat('eta-I_t_case_',num2str(k-1)),pos);
                        
                        yyaxis left
                        
                        plot(t{1,k},AM{1,k},'linewidth',2,'Color',pcols{3});
                        hold on
                        plot(t{1,k},It{1,k}+IAt{1,k}+Pt{1,k},'--','linewidth',2,'Color',pcols{1});
                        plot(t{1,k},n_crit*ones(length(t{k}),1),'k--','linewidth',2);
%                         plot(t{1,1},It{1,1}+IAt{1,1}+Pt{1,1},'-.','linewidth',2,'Color',pcols{end});
                        hold off
%                         ylim([0,30]);
                        figprop(k).ylab = ylabel('Active Cases');
                        figprop(k).ax = gca;
                        figprop(k).fnt_size =14;
                        
                        set(gca,'ycolor','k')
 
                        yyaxis right
                        
                        plot(t{1,k},C{1,k},'linewidth',2,'Color',pcols{2});
                        
                        ylim([0,1000]);

                        figprop(k).ylab2 = ylabel('$C$ (days)');
                        
                        xlim([0,10*365]);
                        figprop(k).xlab = xlabel('$t$ (days since March 10, 2020)');
                        figprop(k).leg = legend({'Tested Active Cases';'Total Active Cases';'$N_{\rm crit}/2$';'Relaxation Cost'},'location','best');
                        set(gca,'ycolor',pcols{2})
                    end
                case 1
                    fig_num = 0;
                    eta_names={'1/4';'1/2';'1';'2';'4'};
                    c_names={'C_c^*/4';'C_c^*/2';'C_c^*';'2C_c^*';'4C_c^*'};
                    row_names={'eta';'c';'health_cost';'cost';'total_cost'};
                    for k1=1:len_m
                        heat_health_data{1}(k1,:)=rel_health_cost(1,(k1-1)*len_m+2:(k1-1)*len_m+6);
                        heat_cost_data{1}(k1,:)=rel_cost(1,(k1-1)*len_m+2:(k1-1)*len_m+6);
                        heat_tot_data{1}(k1,:)=total_cost(1,(k1-1)*len_m+2:(k1-1)*len_m+6);

                    end
                    Omega = linspace(0,1,11);
                    for K = 1:length(Omega)
                        fig_num = fig_num + 1;
                        [fig(fig_num),figprop(fig_num)] = new_figure(strcat('heat_tot_eta','-',num2str(Omega(K)*100),'-',num2str((1-Omega(K))*100)),pos);
                        heatmap(c_names,eta_names,Omega(K)*heat_health_data{1}+(1-Omega(K))*heat_cost_data{1});
                        cmap = hot;
                        colormap(flipud(cmap));
                        figprop(fig_num).type='heat_map';
                        if K == 1
                            annotation('textarrow', [0.05,0.05], [0.3,0.8],'String' , ['Increase' newline 'Concern']);
                            annotation('textarrow', [0.3,0.8], [0.95,0.95],'String' , ['Increase Spending']);
                        end
                        annotation('textbox','string','$\eta$','interpreter','latex','linestyle','none','position',[0.05,0.5,0.1,0.1],'Fontsize',14);
                    end
                    
                    fig_num = fig_num + 1;
                    [fig(fig_num),figprop(fig_num)] = new_figure('eta_active_compare',pos);
                    j = 1;
                    k1 = 13;
                    k2 = 14;
                    k3 = 15;
                    plot(t{j,k1},AM{j,k1},'linewidth',2,'Color',pcols{1});
                    hold on
                    plot(t{j,k2},AM{j,k2},'linewidth',2,'Color',pcols{2});
                    plot(t{j,k3},AM{j,k3},'linewidth',2,'Color',pcols{3});
                    if ~isempty(tAM0{j,k1})
                        line([tAM0{j,k1},tAM1{j,k1}],[0.5,0.5],'Color',pcols{1},'Linestyle','-','linewidth',2);
                        line([tAM0{j,k1},tAM0{j,k1}],[0.4,0.6],'Color',pcols{1},'Linestyle','-','linewidth',2);
                        line([tAM1{j,k1},tAM1{j,k1}],[0.4,0.6],'Color',pcols{1},'Linestyle','-','linewidth',2);
                    end
                    if ~isempty(tAM0{j,k2})
                        line([tAM0{j,k2},t{j,k2}(AMspots1{j,k2}(1)-1)],[0.5,0.5],'Color',pcols{2},'Linestyle','-','linewidth',2);
                        line([tAM0{j,k2},tAM0{j,k2}],[0.4,0.6],'Color',pcols{2},'Linestyle','-','linewidth',2);
                        line([t{j,k2}(AMspots1{j,k2}(1)-1),t{j,k2}(AMspots1{j,k2}(1)-1)],[0.4,0.6],'Color',pcols{2},'Linestyle','-','linewidth',2);
                    end
                    if ~isempty(tAM0{j,k3})
                        line([tAM0{j,k3},t{j,k3}(AMspots1{j,k3}(1)-1)],[0.5,0.5],'Color',pcols{3},'Linestyle','-','linewidth',2);
                        line([tAM0{j,k3},tAM0{j,k3}],[0.4,0.6],'Color',pcols{3},'Linestyle','-','linewidth',2);
                        line([t{j,k3}(AMspots1{j,k3}(1)-1),t{j,k3}(AMspots1{j,k3}(1)-1)],[0.4,0.6],'Color',pcols{3},'Linestyle','-','linewidth',2);
                    end
                    
                    AM1maxX=find(AM{j,k1}==max(AM{j,k1}));
                    AM2maxX=find(AM{j,k2}==max(AM{j,k2}));
                    AM3maxX=find(AM{j,k3}==max(AM{j,k3}));
                    plot(t{j,k1}(AM1maxX),max(AM{j,k1}),'k.');
                    plot(t{j,k2}(AM2maxX),max(AM{j,k2}),'k.');
                    plot(t{j,k3}(AM3maxX),max(AM{j,k3}),'k.');


                    hold off
                    if ~isempty(tAM0{j,k1})
                        text(450,0.2,['$',num2str(round(tAM1{j,k1}-tAM0{j,k1},2)),'$'],'interpreter','latex','Color',pcols{1})
                    end
                    if ~isempty(tAM0{j,k2})
                        text(500,0.4,['$',num2str(round(tAM1{j,k2}-tAM0{j,k2},2)),'$'],'interpreter','latex','Color',pcols{2})
                    end
                    if ~isempty(tAM0{j,k3})
                        text(650,0.4,['$',num2str(round(tAM1{j,k3}-tAM0{j,k3},2)),'$'],'interpreter','latex','Color',pcols{3})
                    end
                    text(t{j,3}(AM1maxX),AM{j,k1}(AM1maxX)+0.05,['$',num2str(round(max(AM{j,k1}),2)),'$'],'interpreter','latex','color',pcols{1})
                    text(t{j,4}(AM2maxX),AM{j,k2}(AM2maxX)+0.05,['$',num2str(round(max(AM{j,k2}),2)),'$'],'interpreter','latex','color',pcols{2})
                    text(t{j,4}(AM3maxX),AM{j,k3}(AM3maxX)+0.05,['$',num2str(round(max(AM{j,k3}),2)),'$'],'interpreter','latex','color',pcols{3})
                    ylim([0,1.5]);
                    xlim([250,1800]);
                    figprop(fig_num).xlab = xlabel('$t$ (days since March 10)');
                    figprop(fig_num).ylab = ylabel('Active Cases');
                    figprop(fig_num).leg = legend('$C_c^*/2$','$C_c^*$','$2C_c^*$','location','northeast');
                    figprop(fig_num).ax = gca;
                    
                    fig_num = fig_num + 1;
                    [fig(fig_num),figprop(fig_num)] = new_figure('eta_active_compare2',pos);
                    j = 1;
                    k1 =2;
                    k2 =6;
                    plot(t{j,k1},AM{j,k1},'linewidth',2,'Color',pcols{1});
                    hold on
                    plot(t{j,k2},AM{j,k2},'linewidth',2,'Color',pcols{2});
                    if ~isempty(tAM0{j,k1})
                        line([tAM0{j,k1},tAM1{j,k1}],[0.5,0.5],'Color',pcols{1},'Linestyle','-','linewidth',2);
                        line([tAM0{j,k1},tAM0{j,k1}],[0.4,0.6],'Color',pcols{1},'Linestyle','-','linewidth',2);
                        line([tAM1{j,k1},tAM1{j,k1}],[0.4,0.6],'Color',pcols{1},'Linestyle','-','linewidth',2);
                    end
                    if ~isempty(tAM0{j,k2})
                        line([tAM0{j,k2},t{j,k2}(AMspots1{j,k2}(1)-1)],[0.5,0.5],'Color',pcols{2},'Linestyle','-','linewidth',2);
                        line([tAM0{j,k2},tAM0{j,k2}],[0.4,0.6],'Color',pcols{2},'Linestyle','-','linewidth',2);
                        line([t{j,k2}(AMspots1{j,k2}(1)-1),t{j,k2}(AMspots1{j,k2}(1)-1)],[0.4,0.6],'Color',pcols{2},'Linestyle','-','linewidth',2);
                    end
                    
                    AM1maxX=find(AM{j,k1}==max(AM{j,k1}));
                    AM2maxX=find(AM{j,k2}==max(AM{j,k2}));
                    plot(t{j,k1}(AM1maxX),max(AM{j,k1}),'k.');
                    plot(t{j,k2}(AM2maxX),max(AM{j,k2}),'k.');


                    hold off
                    if ~isempty(tAM0{j,k1})
                        text(226,0.7,['$',num2str(round(tAM1{j,k1}-tAM0{j,k1},2)),'$'],'interpreter','latex','Color',pcols{1})
                    end
                    if ~isempty(tAM0{j,k2})
                        text(1330,0.4,['$',num2str(round(tAM1{j,k2}-tAM0{j,k2},2)),'$'],'interpreter','latex','Color',pcols{2})
                    end
                    text(t{j,3}(AM1maxX),AM{j,k1}(AM1maxX)+0.05,['$',num2str(round(max(AM{j,k1}),2)),'$'],'interpreter','latex','color',pcols{1})
                    text(t{j,4}(AM2maxX),AM{j,k2}(AM2maxX)+0.05,['$',num2str(round(max(AM{j,k2}),2)),'$'],'interpreter','latex','color',pcols{2})
                    ylim([0,1.5]);
                    xlim([500,2300]);
                    figprop(fig_num).xlab = xlabel('$t$ (days since March 10)');
                    figprop(fig_num).ylab = ylabel('Active Cases');
                    figprop(fig_num).leg = legend('$C_c^*/4$','$4C_c^*$','location','northeast');
                    figprop(fig_num).ax = gca;
                    
                case 2
                    eta_names={'1d4';'1d2';'1';'2';'4'};
                    c_names={'$C_c^*/4$';'$C_c^*/2$';'$C_c^*$';'$2C_c^*$';'$4C_c^*$'};
                    for k=1:len_m   


                        [fig(k),figprop(k)] = new_figure(strcat('sick_case_eta_',eta_names{k}),pos);
                        
                        plot(t{1,1},sick{1,1},'linewidth',2','Color',pcols{end});
                        legentry{1} = 'baseline';
                        hold on
                        for j=1:len_m
                            J = (k-1)*len_m+j+1;
                            plot(t{1,J},sick{1,J},'linewidth',2,'Color',pcols{j});
                            legentry{j+1} = c_names{j};
                        end
                        hold off
                        ylim([0,1]);
                        figprop(k).xlab = xlabel('$t$ (days since March 10, 2020)');
                        figprop(k).ylab = ylabel('\% sick');
                        figprop(k).leg = legend(legentry{:},'location','southeast');
                        figprop(k).ax = gca;
                        figprop(k).fnt_size=14;

                    end
                case 3
                    eta_names={'1/4';'1/2';'1';'2';'4'};
                    c_names={'C_c^*/4';'C_c^*/2';'C_c^*';'2C_c^*';'4C_c^*'};
                    [fig(1),figprop(1)] = new_figure('heat_peak_time_eta',pos);
                    heatmap(c_names,eta_names,reshape(peak_time(1,2:end)/peak_time(1,1),len_m,len_m)');
                    colormap(jet);
                    figprop(1).type='heat_map';
                case 4
                    pos = [2,4.5,7,4.5];
                    fig_num = 0;
                    eta_names={'1d4';'1d2';'1';'2';'4'};
                    c_names={'$C_c^*/4$';'$C_c^*/2$';'$C_c^*$';'$2C_c^*$';'$4C_c^*$'};
                    for k=1:len_m   
                        fig_num = fig_num + 1;
                        [fig(fig_num),figprop(fig_num)] = new_figure(strcat('SD1_case_eta_',eta_names{k}),pos);
                        hold on
                        for j=1:len_m
                            J = (k-1)*len_m+j+1;
                            plot(t{1,J},SD1{1,J},'linewidth',2,'Color',pcols{j});
                            legentry{j+1} = c_names{j};
                        end
                        hold off
                        ylim([0,75]);
%                         xlim([0,1200]);
                        figprop(fig_num).xlab = xlabel('$t$ (days since March 10, 2020)');
                        figprop(fig_num).ylab = ylabel('Social Distancing Class 1');
                        figprop(fig_num).leg = legend(legentry{:},'location','northeast');
                        figprop(fig_num).ax = gca;
                        figprop(fig_num).fnt_size=14;
                        
                        fig_num = fig_num + 1;
                        [fig(fig_num),figprop(fig_num)] = new_figure(strcat('SD2_case_eta',eta_names{k}),pos);
                        hold on
                        for j=1:len_m
                            J = (k-1)*len_m+j+1;
                            plot(t{1,J},SD2{1,J},'linewidth',2,'Color',pcols{j});
                            legentry{j+1} = c_names{j};
                        end
                        hold off
                        ylim([0,45]);
%                         xlim([0,1200]);
                        figprop(fig_num).xlab = xlabel('$t$ (days since March 10, 2020)');
                        figprop(fig_num).ylab = ylabel('Social Distancing Class 2');
                        figprop(fig_num).leg = legend(legentry{:},'location','northeast');
                        figprop(fig_num).ax = gca;
                        figprop(fig_num).fnt_size=14;

                    end
            end
        case 9
            q = [0.9;0.69;0.27];
            multiplier = [8;16;32;64;128];
            cmultiplier = [1/4;1/2;1;2;4];
            len_q = length(q);
            len_m = length(multiplier);
            fprintf('Loading Data...\n');
            for k=1:len_q
    %             K = (k-1)*(len_m^2+1)+1;
                file_title=strcat('moreMc-odemodel-3-q-',num2str(floor(100*q(k))),'-case0.mat');
                DATAS = load(strcat(runpath,file_title));
                params{k,1} = DATAS.params;
                t{k,1} = DATAS.t;
                y = DATAS.y;
                S{k,1} = y(:,1);
                S1{k,1} = y(:,2);
                S2{k,1} = y(:,3);
                E{k,1} = y(:,4);
                E1{k,1} = y(:,5);
                E2{k,1} = y(:,6);
                P{k,1} = y(:,7);
                P1{k,1} = y(:,8);
                P2{k,1} = y(:,9);
                PM{k,1} = y(:,10);
                Is{k,1} = y(:,11);
                Is1{k,1} = y(:,12);
                Is2{k,1} = y(:,13);
                IsM{k,1} = y(:,14);
                Ia{k,1} = y(:,15);
                Ia1{k,1} = y(:,16);
                Ia2{k,1} = y(:,17);
                IaM{k,1} = y(:,18);
                Rs{k,1} = y(:,19);
                Rs1{k,1} = y(:,20);
                Rs2{k,1} = y(:,21);
                RsM{k,1} = y(:,22);
                Ra{k,1} = y(:,23);
                Ra1{k,1} = y(:,24);
                Ra2{k,1} = y(:,25);
                RaM{k,1} = y(:,26);
                M{k,1} = y(:,27);
                C{k,1} = y(:,28);
                for j1=1:len_m
                    for j2=1:len_m
                        K2 = 1 + (j1-1)*len_m + j2;
                        file_title=strcat('moreMc-odemodel-3-q-',num2str(floor(100*q(k))),'-case',num2str((j1-1)*len_m+j2),'.mat');
                        DATAS = load(strcat(runpath,file_title));
                        params{k,K2} = DATAS.params;
                        t{k,K2} = DATAS.t;
                        y = DATAS.y;
                        S{k,K2} = y(:,1);
                        S1{k,K2} = y(:,2);
                        S2{k,K2} = y(:,3);
                        E{k,K2} = y(:,4);
                        E1{k,K2} = y(:,5);
                        E2{k,K2} = y(:,6);
                        P{k,K2} = y(:,7);
                        P1{k,K2} = y(:,8);
                        P2{k,K2} = y(:,9);
                        PM{k,K2} = y(:,10);
                        Is{k,K2} = y(:,11);
                        Is1{k,K2} = y(:,12);
                        Is2{k,K2} = y(:,13);
                        IsM{k,K2} = y(:,14);
                        Ia{k,K2} = y(:,15);
                        Ia1{k,K2} = y(:,16);
                        Ia2{k,K2} = y(:,17);
                        IaM{k,K2} = y(:,18);
                        Rs{k,K2} = y(:,19);
                        Rs1{k,K2} = y(:,20);
                        Rs2{k,K2} = y(:,21);
                        RsM{k,K2} = y(:,22);
                        Ra{k,K2} = y(:,23);
                        Ra1{k,K2} = y(:,24);
                        Ra2{k,K2} = y(:,25);
                        RaM{k,K2} = y(:,26);
                        M{k,K2} = y(:,27);
                        C{k,K2} = y(:,28);
                    end
                end
            end
            fprintf('Data Loaded...\n');

%%%%%%%%%%%%% Now do some processing on the data
            omega = 0.5;
            if process_data_9 == 0 %If looping only process the data once
                n_crit = 1/2; %Value for which we surpass health resources
                for j=1:len_q
                    for k=1:len_m^2+1
                        It{j,k} = Is{j,k}+Is1{j,k}+Is2{j,k}+IsM{j,k};
                        IAt{j,k} = Ia{j,k}+Ia1{j,k}+Ia2{j,k}+IaM{j,k};
                        Pt{j,k} = P{j,k}+P1{j,k}+P2{j,k}+PM{j,k};
                        peak_time(j,k) = t{j,k}(It{j,k}==max(It{j,k}));
                        peak_val(j,k) = max(It{j,k});
                        sickS{j,k} = (Is{j,k}+Is1{j,k}+Is2{j,k}+IsM{j,k}+Rs{j,k}+Rs1{j,k}+Rs2{j,k}+RsM{j,k})*params{j,k}.N0/params{j,k}.N;
                        sickA{j,k} = (Ia{j,k}+Ia1{j,k}+Ia2{j,k}+IaM{j,k}+Ra{j,k}+Ra1{j,k}+Ra2{j,k}+RaM{j,k})*params{j,k}.N0/params{j,k}.N;
                        sick{j,k} = (sickS{j,k} + sickA{j,k});

                        KM{j,k} = 1/log(2)*max((params{j,k}.rho0*(Ia{j,k}+Ia1{j,k}+Ia2{j,k}+P{j,k}+P1{j,k}+P2{j,k}) + params{j,k}.rhoI*(Is{j,k}+Is1{j,k}+Is2{j,k}))./M{j,k},0);
                        AM{j,k} = PM{j,k} + IsM{j,k} + IaM{j,k};
                        Kfun{j,k} = max(KM{j,k}-params{j,k}.Kc,0)./(max(KM{j,k}-params{j,k}.Kc,0)+params{j,k}.K0-params{j,k}.Kc);
                        Afun{j,k} = max(AM{j,k}-params{j,k}.Mc,0)./(max(AM{j,k}-params{j,k}.Mc,0)+params{j,k}.M0-params{j,k}.Mc);
                        mu{j,k} = params{j,k}.mumax*Kfun{j,k}.*Afun{j,k};
                        nu{j,k} = params{j,k}.numax*max(C{j,k}-params{j,k}.Cc,0)./(max(C{j,k}-params{j,k}.Cc,0)+params{j,k}.C0-params{j,k}.Cc);

                        %Find spike times for infection
                        donespikes = 0;
                        Itcount(j,k) = 0;
                        sgnIt = 1;
                        while(~donespikes)
                            sgnIt = sgnIt-1+find(sign(It{j,k}(sgnIt:end)-n_crit)==1,1);
                            if isempty(sgnIt)
                                donespikes=1;
                            else
                                Itcount(j,k) = Itcount(j,k)+1;
                                Itspots0{j,k}(Itcount(j,k)) = sgnIt;
                                tIt0{j,k}(Itcount(j,k))= t{j,k}(sgnIt);
                                sgnIt = sgnIt-1+find(sign(It{j,k}(sgnIt:end)-n_crit)==-1,1);

                                if isempty(sgnIt)
                                    Itspots1{j,k}(Itcount(j,k)) = length(It{j,k});
                                    tIt1{j,k}(Itcount(j,k)) = t{j,k}(end);
                                    t_cut_flag(j,k) = 1;
                                else
                                    Itspots1{j,k}(Itcount(j,k)) = sgnIt;
                                    tIt1{j,k}(Itcount(j,k)) = t{j,k}(sgnIt);
                                    t_cut_flag(j,k) = 0;
                                end

                            end
                        end
                        donespikes = 0;
                        AMcount(j,k) = 0;
                        sgnAM = 1;
                        while(~donespikes)
                            sgnAM = sgnAM-1+find(sign(AM{j,k}(sgnAM:end)-n_crit)==1,1);
                            if isempty(sgnAM)
                                donespikes=1;
                            else
                                AMcount(j,k) = AMcount(j,k)+1;
                                AMspots0{j,k}(AMcount(j,k)) = sgnAM;
                                tAM0{j,k}(AMcount(j,k))= t{j,k}(sgnAM);
                                sgnAM = sgnAM-1+find(sign(AM{j,k}(sgnAM:end)-n_crit)==-1,1);

                                if isempty(sgnAM)
                                    AMspots1{j,k}(AMcount(j,k)) = length(AM{j,k});
                                    tAM1{j,k}(AMcount(j,k)) = t{j,k}(end);
                                    t_AMcut_flag(j,k) = 1;
                                else
                                    AMspots1{j,k}(AMcount(j,k)) = sgnAM;
                                    tAM1{j,k}(AMcount(j,k)) = t{j,k}(sgnAM);
                                    t_AMcut_flag(j,k) = 0;
                                end

                            end
                        end

                        health_cost(j,k) = 0;
                        if AMcount(j,k)~=0
                            for k2=1:length(tAM0{j,k})
                                health_cost(j,k) = health_cost(j,k) + trapz(t{j,k}(AMspots0{j,k}(k2):AMspots1{j,k}(k2)),AM{j,k}(AMspots0{j,k}(k2):AMspots1{j,k}(k2)));
                            end
                        end
                        rel_health_cost(j,k) = health_cost(j,k)/health_cost(j,1);
                        cost(j,k) = C{j,k}(end);
                    end
                    max_cost(j) = max(cost(j,:));
                    rel_cost(j,:) = cost(j,:)/max_cost(j);
                end
%                 max_cost = max(max(cost));
%                 rel_cost = cost/max_cost;
                total_cost = omega*rel_health_cost+(1-omega)*rel_cost;
                for k1=2:len_m^2+1
                    for k2=k1:len_m^2+1
                        rel_total_cost(k1-1,k2-1) = total_cost(2,k2)/total_cost(2,k1);
                    end
                end
                process_data_9 = 1;
            end
%%%%%%%%%%%DONE PROCESSING

            %sub_experiment is instead of all plots, go through the ones we
            %want
            % 0 - It graphs plotted with a critical threshold
            % 1 - heatmap graphs of costs
            % 2 - sick people graphs
            % 3 - peak sick time heat map
            

            switch sub_experiment

                case 0
                    pos = [2,4.5,7,4.5];
                    for k=1:(len_m^2+1)
                        [fig(k),figprop(k)] = new_figure(strcat('moreMc-I_t_case_',num2str(k-1)),pos);
                        temp_It{1} = spline(t{1,k},It{1,k}+IAt{1,k}+Pt{1,k},t{2,k});
                        temp_It{2} = spline(t{3,k},It{3,k}+IAt{3,k}+Pt{3,k},t{2,k});

                        err_bar{k} = [max([temp_It{1} It{2,k}+IAt{2,k}+Pt{2,k} temp_It{2}],[],2)'-(It{2,k}+IAt{2,k}+Pt{2,k})';(It{2,k}+IAt{2,k}+Pt{2,k})'];
                        
                        temp_AM{1} = spline(t{1,k},AM{1,k},t{2,k});
                        temp_AM{2} = spline(t{3,k},AM{3,k},t{2,k});
                        
                        err_barAM{k} = [max([temp_AM{1} AM{2,k} temp_AM{2}],[],2)'-AM{2,k}';AM{2,k}'];
                        
                        temp_It0{1} = spline(t{1,1},It{1,1}+IAt{1,1}+Pt{1,1},t{2,1});
                        temp_It0{2} = spline(t{3,1},It{3,1}+IAt{3,1}+Pt{3,1},t{2,1});
                        
                        err_bar0{k} = [max([temp_It0{1} It{2,1}+IAt{2,1}+Pt{2,1} temp_It0{2}],[],2)'-(It{2,1}+IAt{2,1}+Pt{2,1})';(It{2,1}+IAt{2,1}+Pt{2,1})'];

                        yyaxis left
                        
                        if err_bar_flag
                            shadedErrorBar(t{2,k},AM{2,k},err_barAM{k},'lineprops',{'Color',pcols{3},'linewidth',2});
                            hold on
                            shadedErrorBar(t{2,k},It{2,k}+IAt{2,k}+Pt{2,k},err_bar{k},'lineprops',{'Color',pcols{1},'linewidth',2});
                            shadedErrorBar(t{2,1},It{2,1}+IAt{2,1}+Pt{2,1},err_bar0{k},'lineprops',{'LineStyle','-.','Color',pcols{end},'linewidth',2});
                        else
                            plot(t{2,k},AM{2,k},'linewidth',2,'Color',pcols{3});
                            hold on
                            plot(t{2,k},It{2,k}+IAt{2,k}+Pt{2,k},'--','linewidth',2,'Color',pcols{1});
                            plot(t{2,1},It{2,1}+IAt{2,1}+Pt{2,1},'-.','linewidth',2,'Color',pcols{end});
                        end
                        plot(t{2,k},n_crit*ones(length(t{2,k}),1),'k--','linewidth',2);
                        hold off
                        ylim([0,36]);
                        figprop(k).ylab = ylabel('Active Cases');
                        figprop(k).ax = gca;
                        figprop(k).fnt_size =14;
                        set(gca,'ycolor','k');
                        
                        yyaxis right

                        temp_C{1} = spline(t{1,k},C{1,k},t{2,k});
                        temp_C{2} = spline(t{3,k},C{3,k},t{2,k});

                        err_barC{k} = [max([temp_C{1} C{2,k} temp_C{2}],[],2)'-C{2,k}';C{2,k}'-min([temp_C{1} C{2,k} temp_C{2}],[],2)'];
                        
                        if err_bar_flag
                            shadedErrorBar(t{2,k},C{2,k},err_barC{k},'lineprops',{'Color',pcols{2},'linewidth',2});
                        else
                            plot(t{2,k},C{2,k},'linewidth',2,'Color',pcols{2});
                        end
                        ylim([0,250]);

                        figprop(k).ylab2 = ylabel('$C$ (days)');
                        
                        xlim([0,3*365]);
                        figprop(k).xlab = xlabel('$t$ (days since March 10, 2020)');
                        set(gca,'ycolor',pcols{2});
                    end
                case 1
                    M_names={'8M_c^*';'16M_c^*';'32M_c^*';'64M_c^*';'128M_c^*'};
                    c_names={'C_c^*/4';'C_c^*/2';'C_c^*';'2C_c^*';'4C_c^*'};
                    row_names={'M';'c';'health_cost';'cost';'total_cost'};
                    for k=1:len_q
                        for k1=1:len_m
                            heat_health_data{k}(k1,:)=rel_health_cost(k,(k1-1)*len_m+2:(k1-1)*len_m+6);
                            heat_cost_data{k}(k1,:)=rel_cost(k,(k1-1)*len_m+2:(k1-1)*len_m+6);
                            heat_tot_data{k}(k1,:)=total_cost(k,(k1-1)*len_m+2:(k1-1)*len_m+6);

                        end
                        [fig((k-1)*3+1),figprop((k-1)*3+1)] = new_figure(strcat('moreMc-heat_health_j_',num2str(100*q(k))),pos);
                        heatmap(c_names,M_names,heat_health_data{k});
                        colormap(jet);
                        figprop((k-1)*3+1).type='heat_map';
                        [fig((k-1)*3+2),figprop((k-1)*3+2)] = new_figure(strcat('moreMc-heat_cost_j_',num2str(100*q(k))),pos);
                        heatmap(c_names,M_names,heat_cost_data{k});
                        colormap(jet);
                        figprop((k-1)*3+2).type='heat_map';
                        [fig((k-1)*3+3),figprop((k-1)*3+3)] = new_figure(strcat('moreMc-heat_total_j_',num2str(100*q(k)),'-',num2str(100*omega),'-',num2str(100*(1-omega))),pos);
                        heatmap(c_names,M_names,heat_tot_data{k});
                        colormap(jet);
                        figprop((k-1)*3+3).type='heat_map';
                    end
%                     [fig(3*len_q+1),figprop(3*len_q+1)] = new_figure('active_compare',pos);
%                     plot(t{2,3},AM{2,3},'linewidth',2,'Color',pcols{1});
%                     hold on
%                     plot(t{2,4},AM{2,4},'linewidth',2,'Color',pcols{2});
%                     line([tAM0{2,3},tAM1{2,3}],[0.5,0.5],'Color',pcols{1},'Linestyle','-','linewidth',2);
%                     line([tAM0{2,3},tAM0{2,3}],[0.4,0.6],'Color',pcols{1},'Linestyle','-','linewidth',2);
%                     line([tAM1{2,3},tAM1{2,3}],[0.4,0.6],'Color',pcols{1},'Linestyle','-','linewidth',2);
%                     line([tAM0{2,4},t{2,4}(AMspots1{2,4}(1)-1)],[0.5,0.5],'Color',pcols{2},'Linestyle','-','linewidth',2);
%                     line([tAM0{2,4},tAM0{2,4}],[0.4,0.6],'Color',pcols{2},'Linestyle','-','linewidth',2);
%                     line([t{2,4}(AMspots1{2,4}(1)-1),t{2,4}(AMspots1{2,4}(1)-1)],[0.4,0.6],'Color',pcols{2},'Linestyle','-','linewidth',2);
%                     
%                     AM1maxX=find(AM{2,3}==max(AM{2,3}));
%                     AM2maxX=find(AM{2,4}==max(AM{2,4}));
%                     plot(t{2,3}(AM1maxX),max(AM{2,3}),'k.');
%                     plot(t{2,4}(AM2maxX),max(AM{2,4}),'k.');
% 
% 
%                     hold off
%                     text(226,0.7,['$',num2str(round(tAM1{2,3}-tAM0{2,3},2)),'$'],'interpreter','latex','Color',pcols{1})
%                     text(326,0.7,['$',num2str(round(tAM1{2,4}-tAM0{2,4},2)),'$'],'interpreter','latex','Color',pcols{2})
%                     text(t{2,3}(AM1maxX),AM{2,3}(AM1maxX)+0.1,['$',num2str(round(max(AM{2,3}),2)),'$'],'interpreter','latex','color',pcols{1})
%                     text(t{2,4}(AM2maxX),AM{2,4}(AM2maxX)+0.1,['$',num2str(round(max(AM{2,4}),2)),'$'],'interpreter','latex','color',pcols{2})
%                     ylim([0,4.25]);
%                     xlim([175,385]);
%                     figprop(3*len_q+1).xlab = xlabel('$t$ (days since March 10)');
%                     figprop(3*len_q+1).ylab = ylabel('Active Cases');
%                     figprop(3*len_q+1).leg = legend('$C_c^*/2$','$C_c^*$','location','northeast');
%                     figprop(3*len_q+1).ax = gca;
%                     
%                     [fig(3*len_q+2),figprop(3*len_q+2)] = new_figure('active_compare_2',pos);
%                     plot(t{2,2},AM{2,2},'linewidth',2,'Color',pcols{1});
%                     hold on
%                     plot(t{2,22},AM{2,22},'linewidth',2,'Color',pcols{2});
%                     line([tAM0{2,2},tAM1{2,2}],[0.5,0.5],'Color',pcols{1},'Linestyle','-','linewidth',2);
%                     line([tAM0{2,2},tAM0{2,2}],[0.4,0.6],'Color',pcols{1},'Linestyle','-','linewidth',2);
%                     line([tAM1{2,2},tAM1{2,2}],[0.4,0.6],'Color',pcols{1},'Linestyle','-','linewidth',2);
%                     line([tAM0{2,22},t{2,22}(AMspots1{2,22}(1)-1)],[0.5,0.5],'Color',pcols{2},'Linestyle','-','linewidth',2);
%                     line([tAM0{2,22},tAM0{2,22}],[0.4,0.6],'Color',pcols{2},'Linestyle','-','linewidth',2);
%                     line([t{2,22}(AMspots1{2,22}(1)-1),t{2,22}(AMspots1{2,22}(1)-1)],[0.4,0.6],'Color',pcols{2},'Linestyle','-','linewidth',2);
%                     
%                     AM1maxX2=find(AM{2,2}==max(AM{2,2}));
%                     AM2maxX2=find(AM{2,22}==max(AM{2,22}));
%                     plot(t{2,2}(AM1maxX2),max(AM{2,2}),'k.');
%                     plot(t{2,22}(AM2maxX2),max(AM{2,22}),'k.');
% 
% 
%                     hold off
%                     text(262,0.7,['$',num2str(round(tAM1{2,2}-tAM0{2,2},2)),'$'],'interpreter','latex','Color',pcols{1})
%                     text(175,0.7,['$',num2str(round(tAM1{2,22}-tAM0{2,22},2)),'$'],'interpreter','latex','Color',pcols{2})
%                     text(t{2,2}(AM1maxX2),AM{2,2}(AM1maxX2)+0.1,['$',num2str(round(max(AM{2,2}),2)),'$'],'interpreter','latex','color',pcols{1})
%                     text(t{2,22}(AM2maxX2),AM{2,22}(AM2maxX2)+0.1,['$',num2str(round(max(AM{2,22}),2)),'$'],'interpreter','latex','color',pcols{2})
%                     ylim([0,4.25]);
%                     xlim([0,385]);
%                     figprop(3*len_q+1).xlab = xlabel('$t$ (days since March 10)');
%                     figprop(3*len_q+1).ylab = ylabel('Active Cases');
%                     figprop(3*len_q+1).leg = legend('$M_c^*/4$','$4M_c^*$','location','northeast');
%                     figprop(3*len_q+1).ax = gca;
%                     
%                     [fig(3*len_q+3),figprop(3*len_q+3)] = new_figure('active_compare_3',pos);
%                     plot(t{2,16},AM{2,16},'linewidth',2,'Color',pcols{1});
%                     hold on
%                     plot(t{2,21},AM{2,21},'linewidth',2,'Color',pcols{2});
%                     line([tAM0{2,16},tAM1{2,16}],[0.5,0.5],'Color',pcols{1},'Linestyle','-','linewidth',2);
%                     line([tAM0{2,16},tAM0{2,16}],[0.4,0.6],'Color',pcols{1},'Linestyle','-','linewidth',2);
%                     line([tAM1{2,16},tAM1{2,16}],[0.4,0.6],'Color',pcols{1},'Linestyle','-','linewidth',2);
%                     line([tAM0{2,21},t{2,21}(AMspots1{2,21}(1)-1)],[0.5,0.5],'Color',pcols{2},'Linestyle','-','linewidth',2);
%                     line([tAM0{2,21},tAM0{2,21}],[0.4,0.6],'Color',pcols{2},'Linestyle','-','linewidth',2);
%                     line([t{2,21}(AMspots1{2,21}(1)-1),t{2,22}(AMspots1{2,21}(1)-1)],[0.4,0.6],'Color',pcols{2},'Linestyle','-','linewidth',2);
%                     
%                     AM1maxX3=find(AM{2,16}==max(AM{2,16}));
%                     AM2maxX3=find(AM{2,21}==max(AM{2,21}));
%                     plot(t{2,16}(AM1maxX3),max(AM{2,16}),'k.');
%                     plot(t{2,21}(AM2maxX3),max(AM{2,21}),'k.');
% 
% 
%                     hold off
%                     text(655,0.7,['$',num2str(round(tAM1{2,16}-tAM0{2,16},2)),'$'],'interpreter','latex','Color',pcols{1})
%                     text(750,0.7,['$',num2str(round(tAM1{2,21}-tAM0{2,21},2)),'$'],'interpreter','latex','Color',pcols{2})
%                     text(t{2,16}(AM1maxX3)+2,AM{2,16}(AM1maxX3)+0.1,['$',num2str(round(max(AM{2,16}),2)),'$'],'interpreter','latex','color',pcols{1})
%                     text(t{2,21}(AM2maxX3),AM{2,21}(AM2maxX3)+0.1,['$',num2str(round(max(AM{2,21}),2)),'$'],'interpreter','latex','color',pcols{2})
%                     ylim([0,4.25]);
%                     xlim([600,850]);
%                     figprop(3*len_q+1).xlab = xlabel('$t$ (days since March 10)');
%                     figprop(3*len_q+1).ylab = ylabel('Active Cases');
%                     figprop(3*len_q+1).leg = legend('$M_c^*$','$2M_c^*$','location','northeast');
%                     figprop(3*len_q+1).ax = gca;
                case 2
                    M_names={'8M_c';'16M_c';'32M_c';'64M_c';'128M_c'};
                    c_names={'$C_c^*/4$';'$C_c^*/2$';'$C_c^*$';'$2C_c^*$';'$4C_c^*$'};
                    for k=1:len_m   


                        [fig(k),figprop(k)] = new_figure(strcat('moreMc-sick_case_',M_names{k}),pos);
                        temp_sick{1} = spline(t{1,1},sick{1,1},t{2,1});
                        temp_sick{2} = spline(t{3,1},sick{3,1},t{2,1});
                        err_bar_sick0 = [max([temp_sick{1} sick{2,1} temp_sick{2}],[],2)'-sick{2,1}';sick{2,1}'-min([temp_sick{1} sick{2,1} temp_sick{2}],[],2)'];
                        if err_bar_flag
                            shadedErrorBar(t{2,1},sick{2,1},err_bar_sick0,'lineprops',{'Color',pcols{end},'linewidth',2});
                        else
                            plot(t{2,1},sick{2,1},'--','linewidth',2,'Color',pcols{end});
                        end
                        legentry{1} = 'baseline';
                        hold on
                        for j=1:len_m
                            J = (k-1)*len_m+j+1;
                            temp_sick{1} = spline(t{1,J},sick{1,J},t{2,J});
                            temp_sick{2} = spline(t{3,J},sick{3,J},t{2,J});

                            err_bar_sick{j} = [max([temp_sick{1} sick{2,J} temp_sick{2}],[],2)'-sick{2,J}';sick{2,J}'-min([temp_sick{1} sick{2,J} temp_sick{2}],[],2)'];
                            if err_bar_flag
                                shadedErrorBar(t{2,J},sick{2,J},err_bar_sick{j},'lineprops',{'Color',pcols{j},'linewidth',2});
                            else
                                plot(t{2,J},sick{2,J},'linewidth',2,'Color',pcols{j});
                            end
                            legentry{j+1} = c_names{j};
                        end
                        hold off
                        ylim([0,1]);
                        xlim([0,1200]);
                        figprop(k).xlab = xlabel('$t$ (days since March 10, 2020)');
                        figprop(k).ylab = ylabel('\% sick');
                        figprop(k).leg = legend(legentry{:},'location','southeast');
                        figprop(k).ax = gca;
                        figprop(k).fnt_size=14;

                    end
                case 3
                    M_names={'8M_c^*';'16M_c^*';'32M_c^*';'64M_c^*';'128M_c^*'};
                    c_names={'C_c^*/4';'C_c^*/2';'C_c^*';'2C_c^*';'4C_c^*'};
                    for k=1:len_q
                        [fig(k),figprop(k)] = new_figure(strcat('moreMc-heat_peak_time_j_',num2str(100*q(k))),pos);
                        heatmap(c_names,M_names,reshape(peak_time(k,2:end)/peak_time(k,1),len_m,len_m)');
                        colormap(jet);
                        figprop(k).type='heat_map';
                    end
            end
            
        case 10
            lenMC = 5000;
            k = 1;
            fprintf('Loading Data...\n');
            file_title=strcat('testMc-odemodel-3-case0.mat');
            DATAS = load(strcat(runpath,file_title));
            params{k,1} = DATAS.params;
            t{k,1} = DATAS.t;
            y = DATAS.y;
            S{k,1} = y(:,1);
            S1{k,1} = y(:,2);
            S2{k,1} = y(:,3);
            E{k,1} = y(:,4);
            E1{k,1} = y(:,5);
            E2{k,1} = y(:,6);
            P{k,1} = y(:,7);
            P1{k,1} = y(:,8);
            P2{k,1} = y(:,9);
            PM{k,1} = y(:,10);
            Is{k,1} = y(:,11);
            Is1{k,1} = y(:,12);
            Is2{k,1} = y(:,13);
            IsM{k,1} = y(:,14);
            Ia{k,1} = y(:,15);
            Ia1{k,1} = y(:,16);
            Ia2{k,1} = y(:,17);
            IaM{k,1} = y(:,18);
            Rs{k,1} = y(:,19);
            Rs1{k,1} = y(:,20);
            Rs2{k,1} = y(:,21);
            RsM{k,1} = y(:,22);
            Ra{k,1} = y(:,23);
            Ra1{k,1} = y(:,24);
            Ra2{k,1} = y(:,25);
            RaM{k,1} = y(:,26);
            M{k,1} = y(:,27);
            C{k,1} = y(:,28);
            for j1=1:lenMC
                K2=j1+1;
                file_title=strcat('testMc-odemodel-3-case',num2str(j1),'.mat');
                DATAS = load(strcat(runpath,file_title));
                params{k,K2} = DATAS.params;
                t{k,K2} = DATAS.t;
                y = DATAS.y;
                S{k,K2} = y(:,1);
                S1{k,K2} = y(:,2);
                S2{k,K2} = y(:,3);
                E{k,K2} = y(:,4);
                E1{k,K2} = y(:,5);
                E2{k,K2} = y(:,6);
                P{k,K2} = y(:,7);
                P1{k,K2} = y(:,8);
                P2{k,K2} = y(:,9);
                PM{k,K2} = y(:,10);
                Is{k,K2} = y(:,11);
                Is1{k,K2} = y(:,12);
                Is2{k,K2} = y(:,13);
                IsM{k,K2} = y(:,14);
                Ia{k,K2} = y(:,15);
                Ia1{k,K2} = y(:,16);
                Ia2{k,K2} = y(:,17);
                IaM{k,K2} = y(:,18);
                Rs{k,K2} = y(:,19);
                Rs1{k,K2} = y(:,20);
                Rs2{k,K2} = y(:,21);
                RsM{k,K2} = y(:,22);
                Ra{k,K2} = y(:,23);
                Ra1{k,K2} = y(:,24);
                Ra2{k,K2} = y(:,25);
                RaM{k,K2} = y(:,26);
                M{k,K2} = y(:,27);
                C{k,K2} = y(:,28);
            end
            fprintf('Data Loaded...\n');
            
            %%%%%%%%%%%%% Now do some processing on the data
            omega = 0.5;
            n_crit = 1/2;
            if process_data_10 == 0 %If looping only process the data once
                j = 1;
                for k=1:lenMC+1
                    It{j,k} = Is{j,k}+Is1{j,k}+Is2{j,k}+IsM{j,k};
                    IAt{j,k} = Ia{j,k}+Ia1{j,k}+Ia2{j,k}+IaM{j,k};
                    Pt{j,k} = P{j,k}+P1{j,k}+P2{j,k}+PM{j,k};
                    peak_time(j,k) = t{j,k}(It{j,k}==max(It{j,k}));
                    peak_val(j,k) = max(It{j,k});
                    sickS{j,k} = (Is{j,k}+Is1{j,k}+Is2{j,k}+IsM{j,k}+Rs{j,k}+Rs1{j,k}+Rs2{j,k}+RsM{j,k})*params{j,k}.N0/params{j,k}.N;
                    sickA{j,k} = (Ia{j,k}+Ia1{j,k}+Ia2{j,k}+IaM{j,k}+Ra{j,k}+Ra1{j,k}+Ra2{j,k}+RaM{j,k})*params{j,k}.N0/params{j,k}.N;
                    sick{j,k} = (sickS{j,k} + sickA{j,k});

                    KM{j,k} = 1/log(2)*max((params{j,k}.rho0*(Ia{j,k}+Ia1{j,k}+Ia2{j,k}+P{j,k}+P1{j,k}+P2{j,k}) + params{j,k}.rhoI*(Is{j,k}+Is1{j,k}+Is2{j,k}))./M{j,k},0);
                    AM{j,k} = PM{j,k} + IsM{j,k} + IaM{j,k};
                    Kfun{j,k} = max(KM{j,k}-params{j,k}.Kc,0)./(max(KM{j,k}-params{j,k}.Kc,0)+params{j,k}.K0-params{j,k}.Kc);
                    Afun{j,k} = max(AM{j,k}-params{j,k}.Mc,0)./(max(AM{j,k}-params{j,k}.Mc,0)+params{j,k}.M0-params{j,k}.Mc);
                    mu{j,k} = params{j,k}.mumax*Kfun{j,k}.*Afun{j,k};
                    nu{j,k} = params{j,k}.numax*max(C{j,k}-params{j,k}.Cc,0)./(max(C{j,k}-params{j,k}.Cc,0)+params{j,k}.C0-params{j,k}.Cc);

                    %Find spike times for infection
                    donespikes = 0;
                    Itcount(j,k) = 0;
                    sgnIt = 1;
                    while(~donespikes)
                        sgnIt = sgnIt-1+find(sign(It{j,k}(sgnIt:end)-n_crit)==1,1);
                        if isempty(sgnIt)
                            donespikes=1;
                        else
                            Itcount(j,k) = Itcount(j,k)+1;
                            Itspots0{j,k}(Itcount(j,k)) = sgnIt;
                            tIt0{j,k}(Itcount(j,k))= t{j,k}(sgnIt);
                            sgnIt = sgnIt-1+find(sign(It{j,k}(sgnIt:end)-n_crit)==-1,1);

                            if isempty(sgnIt)
                                Itspots1{j,k}(Itcount(j,k)) = length(It{j,k});
                                tIt1{j,k}(Itcount(j,k)) = t{j,k}(end);
                                t_cut_flag(j,k) = 1;
                            else
                                Itspots1{j,k}(Itcount(j,k)) = sgnIt;
                                tIt1{j,k}(Itcount(j,k)) = t{j,k}(sgnIt);
                                t_cut_flag(j,k) = 0;
                            end

                        end
                    end
                    donespikes = 0;
                    AMcount(j,k) = 0;
                    sgnAM = 1;
                    while(~donespikes)
                        sgnAM = sgnAM-1+find(sign(AM{j,k}(sgnAM:end)-n_crit)==1,1);
                        if isempty(sgnAM)
                            donespikes=1;
                        else
                            AMcount(j,k) = AMcount(j,k)+1;
                            AMspots0{j,k}(AMcount(j,k)) = sgnAM;
                            tAM0{j,k}(AMcount(j,k))= t{j,k}(sgnAM);
                            sgnAM = sgnAM-1+find(sign(AM{j,k}(sgnAM:end)-n_crit)==-1,1);

                            if isempty(sgnAM)
                                AMspots1{j,k}(AMcount(j,k)) = length(AM{j,k});
                                tAM1{j,k}(AMcount(j,k)) = t{j,k}(end);
                                t_AMcut_flag(j,k) = 1;
                            else
                                AMspots1{j,k}(AMcount(j,k)) = sgnAM;
                                tAM1{j,k}(AMcount(j,k)) = t{j,k}(sgnAM);
                                t_AMcut_flag(j,k) = 0;
                            end

                        end
                    end

                    health_cost(j,k) = 0;
                    if AMcount(j,k)~=0
                        for k2=1:length(tAM0{j,k})
                            health_cost(j,k) = health_cost(j,k) + trapz(t{j,k}(AMspots0{j,k}(k2):AMspots1{j,k}(k2)),AM{j,k}(AMspots0{j,k}(k2):AMspots1{j,k}(k2)));
                        end
                    end
                    rel_health_cost(j,k) = health_cost(j,k)/health_cost(j,1);
                    cost(j,k) = C{j,k}(end);
                end
                max_cost(j) = max(cost(j,:));
                rel_cost(j,:) = cost(j,:)/max_cost(j);
                total_cost = omega*rel_health_cost+(1-omega)*rel_cost;
                for k=2:lenMC
                    Mc(k) = params{1,k}.Mc*params{1,k}.N_crit/params{1,k}.N;
                end
                process_data_10 = 1;
            end
%%%%%%%%%%%DONE PROCESSING
            [fig(1),figprop(1)] = new_figure('health_cost_compare',pos);
            plot(Mc(2:end)*100,rel_health_cost(3:end),'linewidth',2,'Color',pcols{1});
            hold on
            plot(params{1}.Mc*params{1}.N_crit/params{1}.N*ones(1000,1)*100,linspace(0,1,1000),'k--','linewidth',2);
            hold off
            annotation('textarrow',[0.23,0.135],[0.15,0.15],'String','M_c^*')
            xlim([0,2]);
            ylim([0,1]);
            figprop(1).xlab = xlabel('$M_c$ (\% $N$)');
            figprop(1).ylab = ylabel('$H/H_\infty$');
            figprop(1).ax = gca;
            figprop(1).fnt_size =14;
            
    end
    
    if loop_flag
        if sub_len(experiment+1)>0
            sub_experiment = sub_experiment +1;
                if sub_experiment>sub_len(experiment+1)
                    sub_experiment = 0;
                    experiment = experiment +1;
                    clr_flag = 1;
                end
        else
            experiment = experiment +1;
            clr_flag = 1;
        end
        if clr_flag
            clearvars -except process_data_2 process_data_5 process_data_8 process_data_9 process_data_10 loop_flag pcols curpath newpath plotpath runpath pos exp_len sub_len done experiment sub_experiment fig figprop err_bar_flag
        end
        if experiment > exp_len
            done =1;
        end
    end
    figure_revise(pos,plotpath,fig,figprop);
end
```
