## Supplementary material for "Cost and social distancing dynamics in a mathematical model of COVID-19 with application to Ontario, Canada": Code: CC_PLOT.m

```
%% Plot X (input, LHS matrix) and Y (output) at column s (time point)
%% type: either linear-linear plot ('lin') or linear-log plot '(log')
%% var: labels of the parameters varied in the X (as legend)
%% The Title of the plot is the Pearson correlation coefficient
%% by Simeone Marino, June 5 2007 %%
function CC_PLOT(X,Y,s,type,PRCC_var,y_var)
[A p]=corr(X,Y(s,:)');
[a b]=size(X);
for i=1:b
    a=['[Pearson correlation , p-value] = ' '[' num2str(A(i)) ' , '  num2str(p(i)) '].'];% ' Time point=' num2str(s-1)]
    c=['X(:,',num2str(i),');'];
    if type=='lin'
        figure,plot(eval(c),Y(s,:),'.'),Title(a),legend(PRCC_var{i}),...%
            xlabel(PRCC_var{i}),ylabel(y_var);%eval(c6);
    elseif type=='log'
        figure,semilogy(eval(c),Y(s,:),'.'),Title(a),legend(PRCC_var{i}),...%
            xlabel(PRCC_var{i}),ylabel(y_var);%eval(c6);
    end
end
```
