## Supplementary material for "Cost and social distancing dynamics in a mathematical model of COVID-19 with application to Ontario, Canada": Code: LHS_Call.m

```
function s=LHS_Call(xmin,xmean,xmax,xsd,nsample,distrib,threshold)
% s=latin_hs(xmean,xsd,nsample,nvar)
% LHS from normal distribution, no correlation
% method of Stein
% Stein, M. 1987. Large Sample Properties of Simulations Using Latin Hypercube Sampling. 
%                 Technometrics 29:143-151

if nsample==1
    s=xmean;
    return
end
if nargin<7
    threshold=1e20;
end

[sample,nvar]=size(xmean);
if distrib == 'norm'   % you only need to specify xmean & xsd
    ran=rand(nsample,nvar);
    s=zeros(nsample,nvar);
    %method of Stein
    for j=1: nvar
        idx=randperm(nsample);
        P=(idx'-ran(:,j))/nsample;       % probability of the cdf
        s(:,j) = xmean(j) + ltqnorm(P).* xsd(j); % this can be replaced by any inverse distribution function
    end
end


if distrib == 'unif'  % you only need to specify xmin & xmax
    if xmin==0
        xmin=1e-300;
    end
    nvar=length(xmin);
    ran=rand(nsample,nvar);
    s=zeros(nsample,nvar);
    for j=1: nvar
        idx=randperm(nsample);
        P =(idx'-ran(:,j))/nsample;
        xmax(j);
        xmin(j);
        xmax(j)/xmin(j);
        if (xmax(j)<1 & xmin(j)<1) || (xmax(j)>1 & xmin(j)>1)
            'SAME RANGE';
            if (xmax(j)/xmin(j))<threshold %% It uses the log scale if the order of magnitude of [xmax-xmin] is bigger than threshold
                '<1e3: LINEAR SCALE';
                s(:,j) = xmin(j) + P.* (xmax(j)-xmin(j));
            else
                '>=1e3: LOG SCALE';
                s(:,j) = log(xmin(j)) + P.*abs(abs(log(xmax(j)))-abs(log(xmin(j))));
                s(:,j) = exp(s(:,j));
            end
        else
            'e- to e+';
            if (xmax(j)/xmin(j))<threshold %% It uses the log scale if the order of magnitude of [xmax-xmin] is bigger than threshold
                '<1e3: LINEAR SCALE';
                s(:,j) = xmin(j) + P.* (xmax(j)-xmin(j));
            else
                '>=1e3: LOG SCALE';
                s(:,j) = log(xmin(j)) + P.*abs(log(xmax(j))-log(xmin(j)));
                s(:,j) = exp(s(:,j));
            end
        end
    end
end
%hist(s)   % plots the histogram of the pdf
```
