## Supplementary material for "Cost and social distancing dynamics in a mathematical model of COVID-19 with application to Ontario, Canada": Code: Model_LHS.m

```
%% The results should be compared to the PRCC results section in
%% Supplementary Material D and Table D.1 for different N (specified by
%% "runs" in the script below
clear variables;
close all;

if exist('Model_LHS.mat','file')~=0
    resp = input('This file already exists, rerun? Y/N\n','s');
    if strcmpi(resp,'Y')
        run_sims=1;
    else
        run_sims=0;
    end
else
    run_sims = 1;
end

if run_sims

    %% Sample size N
    runs=10000;

    %% LHS MATRIX  %%
    Parameter_settings_LHS;

    fprintf('Setting random parameters...\n');
    alpha_LHS=LHS_Call(0.1, params.alpha, 0.9, 0 ,runs,'unif'); 
    sigma_LHS=LHS_Call(1/7, params.sigma, 10, 0 ,runs,'unif');
    phi_LHS=LHS_Call(1/12, params.phi, 2, 0, runs,'unif'); 
    gamma_LHS=LHS_Call(1/26.3,params.gamma,1/3.6, 0 ,runs,'unif');
    q_LHS=LHS_Call(0.2, params.q, 0.98, 0, runs,'unif');
    q1_LHS= LHS_Call(0.01 , params.q1 , 1 , 0 , runs , 'unif');
    q2_LHS=LHS_Call(0.01,params.q2,1, 0 ,runs,'unif'); 
    muI_LHS=LHS_Call(0.001,params.muI,1, 0 ,runs,'unif');
    qI_LHS=LHS_Call(0.01,params.qI,1, 0 ,runs,'unif'); 
    rho0_LHS=LHS_Call(params.rho0/100,params.rho0,params.rho0*100, 0 ,runs,'unif');
    rhoI_LHS=LHS_Call(params.rhoI/100,params.rhoI,params.rhoI*100, 0 ,runs,'unif');
    fprintf('Random parameters set...\n');

    %% LHS MATRIX and PARAMETER LABELS
    LHSmatrix=[alpha_LHS sigma_LHS phi_LHS gamma_LHS q_LHS ...
                  q1_LHS q2_LHS muI_LHS  qI_LHS...
                  rho0_LHS rhoI_LHS];
    tic
    parfor x=1:runs %Run solution x times choosing different values
        f=@ODE_LHS;
        fprintf('Running simulation for parameter set %i...\n',x);
    %     LHSmatrix(x,:)
%         y0 = [params.N-LHSmatrix(x,end);0;0;0;0;0;0;0;0;0;LHSmatrix(x,end);0;0;0;0;0;0;0;0;0;0;0;0;0;0;0;0;0];
%         y0 = y0/params.N0;
        odeopts = odeset('NonNegative',1,'RelTol',1e-8,'AbsTol',1e-9);
        [t,y]=ode23t(@(t,y)f(t,y,LHSmatrix,x,runs),tspan,y0,odeopts);
        It=sum(y(:,11:14)')';
        max_spot=find(It==max(It));
        peak_val(x) = It(max_spot);
        peak_time(x) = t(max_spot);
        A=[t y]; % [time y]
        %% Save the outputs at ALL time points [tspan]
        %T_lhs(:,x)=Anew(:,1);
        %CD4_lhs(:,x)=Anew(:,2);
        %T1_lhs(:,x)=Anew(:,3);
        %T2_lhs(:,x)=Anew(:,4);
        %V_lhs(:,x)=Anew(:,5);

        %% Save only the outputs at the time points of interest [time_points]:
        %% MORE EFFICIENT
        time(:,x) = A(time_points+1,1);
        S(:,x) = A(time_points+1,2);
        S1(:,x) = A(time_points+1,3);
        S2(:,x) = A(time_points+1,4);
        E(:,x) = A(time_points+1,5);
        E1(:,x) = A(time_points+1,6);
        E2(:,x) = A(time_points+1,7);
        P(:,x) = A(time_points+1,8);
        P1(:,x) = A(time_points+1,9);
        P2(:,x) = A(time_points+1,10);
        PM(:,x) = A(time_points+1,11);
        Is(:,x) = A(time_points+1,12);
        Is1(:,x) = A(time_points+1,13);
        Is2(:,x) = A(time_points+1,14);
        IsM(:,x) = A(time_points+1,15);
        Ia(:,x) = A(time_points+1,16);
        Ia1(:,x) = A(time_points+1,17);
        Ia2(:,x) = A(time_points+1,18);
        IaM(:,x) = A(time_points+1,19);
        Rs(:,x) = A(time_points+1,20);
        Rs1(:,x) = A(time_points+1,21);
        Rs2(:,x) = A(time_points+1,22);
        RsM(:,x) = A(time_points+1,23);
        Ra(:,x) = A(time_points+1,24);
        Ra1(:,x) = A(time_points+1,25);
        Ra2(:,x) = A(time_points+1,26);
        RaM(:,x) = A(time_points+1,27);
        M(:,x) = A(time_points+1,28);
        C(:,x) = A(time_points+1,29);


    end
    time_end = toc;
    fprintf('Simulations completed in %3.2fs...\n',time_end);
    %% Save the workspace
    save Model_LHS.mat;
end
```
