## Supplementary material for "Cost and social distancing dynamics in a mathematical model of COVID-19 with application to Ontario, Canada": Code: ODE_LHS.m

```
function dy=covidmodel(t,y,LHSmatrix,x,runs)
%% PARAMETERS %%
Parameter_settings_LHS;

params.alpha=LHSmatrix(x,1);
params.sigma=LHSmatrix(x,2);
params.phi=LHSmatrix(x,3);
params.gamma=LHSmatrix(x,4);
params.q=LHSmatrix(x,5);
params.q1=LHSmatrix(x,6);
params.q2=LHSmatrix(x,7);
params.muI=LHSmatrix(x,8);
params.qI=LHSmatrix(x,9);
params.rho0=LHSmatrix(x,10);
params.rhoI=LHSmatrix(x,11);

params.beta = params.R0*params.phi*params.gamma/(params.alpha*(params.gamma+params.phi)+(1-params.alpha)*params.q*params.phi);

dy = zeros(22,1);
        
S = y(1);
S1 = y(2);
S2 = y(3);
E = y(4);
E1 = y(5);
E2 = y(6);
P = y(7);
P1 = y(8);
P2 = y(9);
PM = y(10);
Is = y(11);
Is1 = y(12);
Is2 = y(13);
IsM = y(14);
Ia = y(15);
Ia1 = y(16);
Ia2 = y(17);
IaM = y(18);
Rs = y(19);
Rs1 = y(20);
Rs2 = y(21);
RsM = y(22);
Ra = y(23);
Ra1 = y(24);
Ra2 = y(25);
RaM = y(26);
M = y(27);
C = y(28);

 F_S = params.N0/params.N*params.beta*(params.alpha*P*S+params.alpha*Ia*S+Is*S...
            +params.alpha*params.delta*P1*S+params.alpha*params.delta*Ia1*S+params.delta*Is1*S);
F_S1 = params.N0/params.N*params.beta*(params.alpha*params.delta*P*S1+params.alpha*params.delta*Ia*S1+params.delta*Is*S1...
            +params.alpha*params.delta^2*P1*S1+params.alpha*params.delta^2*Ia1*S1+params.delta^2*Is1*S1);

KM = min(1/log(2)*max((params.rho0*(Ia+Ia1+Ia2+P+P1+P2) + params.rhoI*(Is+Is1+Is2))/M,0),1000);
AM = PM+IsM+IaM;

mu = params.mumax*max(KM-params.Kc,0)/(max(KM-params.Kc,0)+params.K0-params.Kc)*max(AM-params.Mc,0)/(max(AM-params.Mc,0)+params.M0-params.Mc);
mu1 = mu/2;

nu2 = params.numax*max(C-params.Cc,0)/(max(C-params.Cc,0)+params.C0-params.Cc);
nu = nu2/2;

        

        
        dy(1) = -F_S - mu*S + nu*S1 + (1-params.q2)*nu2*S2;
        dy(2) = -F_S1 - mu1*S1 + params.q1*mu*S - nu*S1 +params.q2*nu2*S2;
        dy(3) = (1-params.q1)*mu*S + mu1*S1 - nu2*S2;
        dy(4) = F_S - mu*E + nu*E1 + (1-params.q1)*nu2*E2 -params.sigma*E;
        dy(5) = F_S1 - mu1*E1 + params.q1*mu*E - nu*E1 + params.q1*nu2*E2 - params.sigma*E1;
        dy(6) = (1-params.q1)*mu*E + mu1*E1 - nu2*E2 - params.sigma*E2;
        dy(7) = params.sigma*E -mu*P + nu*P1 + (1-params.q1)*nu2*P2 - params.phi*P - params.rho0*P;
        dy(8) = params.sigma*E1 -mu1*P1 + params.q1*mu*P - nu*P1 + params.q1*nu2*P2 - params.phi*P1 - params.rho0*P1 ;
        dy(9) = params.sigma*E2 + (1-params.q1)*mu*P + mu1*P1 - nu2*P2 - params.phi*P2 - params.rho0*P2;
        dy(10) = params.rho0*(P+P1+P2) - params.phi*PM;
        dy(11) = params.q*params.phi*P - params.muI*Is - params.gamma*Is - params.rhoI*Is;
        dy(12) = params.q*params.phi*P1 + params.qI*params.muI*Is - params.gamma*Is1 - params.rhoI*Is1;
        dy(13) = params.q*params.phi*P2 + (1-params.qI)*params.muI*Is - params.gamma*Is2 - params.rhoI*Is2;
        dy(14) = params.rhoI*(Is+Is1+Is2) + params.q*params.phi*PM - params.gamma*IsM;
        dy(15) = (1-params.q)*params.phi*P - mu*Ia +nu*Ia1 + (1-params.q1)*nu2*Ia2 - params.gamma*Ia - params.rho0*Ia;
        dy(16) = (1-params.q)*params.phi*P1 - mu1*Ia1 + params.q1*mu*Ia - nu*Ia1 + params.q1*nu2*Ia2 - params.gamma*Ia1 - params.rho0*Ia1;
        dy(17) = (1-params.q)*params.phi*P2 + (1-params.q1)*mu*Ia + mu1*Ia1 - nu2*Ia2 - params.gamma*Ia2 - params.rho0*Ia2;
        dy(18) = params.rho0*(Ia+Ia1+Ia2) +(1-params.q)*params.phi*PM - params.gamma*IaM;
        dy(19) = params.gamma*Is;
        dy(20) = params.gamma*Is1;
        dy(21) = params.gamma*Is2;
        dy(22) = params.gamma*IsM;
        dy(23) = params.gamma*Ia;
        dy(24) = params.gamma*Ia1;
        dy(25) = params.gamma*Ia2;
        dy(26) = params.gamma*IsM;
        dy(27) = params.rho0*(Ia+Ia1+Ia2+P+P1+P2) + params.rhoI*(Is+Is1+Is2);
        dy(28) = params.n*((S2+E2)+(1-params.delta)*(S1+E1)) - params.muC*C;
end
```
