## Supplementary material for "Cost and social distancing dynamics in a mathematical model of COVID-19 with application to Ontario, Canada": Code: Parameter_settings_LHS.m

```
% PARAMETER BASELINE VALUES
params.alpha = 1/2;
params.sigma = 2;
params.phi = 1/4.6;
params.gamma = 1/10;
params.q = 0.69;
params.q1 = 0.9;
params.q2 = 0.6;
params.muI = 0.01;
params.qI = 0.6;
params.rho0 = 8.7E-3;
params.rhoI = 4*params.rho0;

params.N = 13448494; 
params.N_crit = 10000/0.123; 
params.N0=params.N_crit;
params.n = params.N0/params.N;
params.R0 = 2.4;
params.delta = 0.25;
params.mumax = 1;
params.numax = params.mumax;
params.Mc = 2090/params.N0;
params.M0 = 2*params.Mc;
params.Kc = 1/16.24;
params.K0 = 4*params.Kc;
params.Cc = 50;
params.C0 = 2*params.Cc;
params.muC = 0;


% Parameter Labels 
PRCC_var={'\alpha', '\sigma', '\phi', ...
    '\gamma','q', 'q_1','q_2', '\mu_I','q_I',...
    '\rho_0', '\rho_I'};% 

%% TIME SPAN OF THE SIMULATION
t_end=3*365; % length of the simulations
tspan=(0:1:t_end);   % time points where the output is calculated
time_points=[round(t_end/2) t_end]; % time points of interest for the US analysis

% INITIAL CONDITION FOR THE ODE MODEL
infected_initial = 2E-4*params.N;
        
y0 = [params.N-infected_initial;0;0;0;0;0;0;0;0;0;infected_initial;0;0;0;0;0;0;0;0;0;0;0;0;0;0;0;0;0];
y0 = y0/params.N0;

% Variables Labels
y_var_label={'S';'S_1';'S_2';'E';'E_1';'E_2';'P';'P_1';'P_2';'P_M';'I_S';'I_{S1}';'I_{S2}';'I_{SM}';'I_A';'I_{A1}';'I_{A2}';'I_{AM}';'R_S';'R_{S1}';'R_{S2}';'R_{SM}';'R_A';'R_{A1}';'R_{A2}';'R_{AM}';'M';'C'};
```
