## Supplementary material for "Cost and social distancing dynamics in a mathematical model of COVID-19 with application to Ontario, Canada": Code: PRCC.m

```
%% It calculates PRCCs and their significances
%% (uncorrected p-value, Bonferroni correction and 
%% Benjamini and Hochberg False Discovery Rate correction)
%% LHSmatrix: LHS matrix (N x k) %%
%% Y: output matrix (time x N) %%
%% s: time points to test (row vector) %%
%% PRCC_var: vector of strings {'p1','p2',...,'pk'}
%%            to label all the parameters varied in the LHS
%% by Simeone Marino, May 29 2007 %%
%% N.B.: this version uses ONE output at a time

function [prcc,sign,sign_label]=PRCC(LHSmatrix,Y,s,PRCC_var,alpha)

%LHSmatrix=LHS; % Define LHS matrix
Y=Y(s,:)';% Define the output. Comment out if the Y is already 
          % a subset of all the time points and it already comprises
          % ONLY the s rows of interest
[a,k]=size(LHSmatrix); % Define the size of LHS matrix
[b,out]=size(Y);
for i=1:k  % Loop for the whole submatrices
    c=['LHStemp=LHSmatrix;LHStemp(:,',num2str(i),')=[];Z',num2str(i),'=LHStemp;LHStemp=[];'];
    eval(c);
    % Loop to calculate PRCCs and significances
    c1=['[LHSmatrix(:,',num2str(i),'),Y];'];
    c2=['Z',num2str(i)];
    [rho,p]=partialcorr(eval(c1),eval(c2),'type','Spearman');
    for j=1:out
        c3=['prcc_',num2str(i),'(',num2str(j),')=rho(1,',num2str(j+1),');'];
        c4=['prcc_sign_',num2str(i),'(',num2str(j),')=p(1,',num2str(j+1),');'];
        eval(c3);
        eval(c4);
    end
    c5=['clear Z',num2str(i),';'];
    eval(c5);
end
prcc=[];
prcc_sign=[];
for i=1:k
    d1=['prcc=[prcc ; prcc_',num2str(i),'];'];
    eval(d1);
    d2=['prcc_sign=[prcc_sign ; prcc_sign_',num2str(i),'];'];
    eval(d2);
end
[length(s) k out];
PRCCs=prcc';
uncorrected_sign=prcc_sign';
prcc=PRCCs;
sign=uncorrected_sign;

%% Multiple tests correction: Bonferroni and FDR
%tests=length(s)*k; % # of tests performed
%correction_factor=tests;
%Bonf_sign=uncorrected_sign*tests;
%sign_new=[];
%for i=1:length(s)
%    sign_new=[sign_new;(1:k)',ones(k,1)*s(i),sign(i,:)'];
%end
%sign_new=sortrows(sign_new,3);
%for j=2:k
%    sign_new(j,3)=sign_new(j,3)*(tests/(tests-j+1));
%end
%sign_new=sortrows(sign_new,[2 1]); % FDR correction
%sign_new=sign_new(:,3)';
%for i=1:length(s)
%    FDRsign(i,:)=[sign_new(1+k*(i-1):i*k)];
%end
%uncorrected_sign; % uncorrected p-value
%Bonf_sign;  % Bonferroni correction
%FDRsign; % FDR correction

sign_label_struct=struct;
sign_label_struct.uncorrected_sign=uncorrected_sign;
%sign_label_struct.value=prcc;

%figure
for r=1:length(s)
    c1=['PRCCs at time = ' num2str(s(r))];
    a=find(uncorrected_sign(r,:)<alpha);
    ['Significant PRCCs'];
    a;
    PRCC_var(a);
    prcc(r,a);
    b=num2str(prcc(r,a));
    sign_label_struct.index{r}=a;
    sign_label_struct.label{r}=PRCC_var(a);
    sign_label_struct.value{r}=b;
    %sign_label_struct.value(r)=b;
    %% Plots of PRCCs and their p-values
    %subplot(1,2,1),bar(PRCCs(r,:)),Title(c1);%,set(gca,'XTickLabel',PRCC_var,'XTick',[1:k]),Title('PRCCs');
    %subplot(1,2,2),bar(uncorrected_sign(r,:)),...
    %    set(gca,'YLim',[0,.1]),Title('P values');%'XTickLabel',PRCC_var,'XTick',[1:k],
end
sign_label=sign_label_struct;
```
