## Supplementary material for "Cost and social distancing dynamics in a mathematical model of COVID-19 with application to Ontario, Canada": Code: PRCC_PLOT.m

```
%% Plot the residual of the partial regression of X (input - LHS matrix) and Y (output)
%% at column s (time points saved). PCC Coefficients are calculated on these
%% var: labels of the parameters varied in the X (as legend)
%% The Title of the plot is the Pearson correlation coefficient of the
%% transformed data, that is  the PRCC calculated on the original data.
%% The p-value is also showed in the title
%% by Simeone Marino, June 5 2007 %%

function PRCC_PLOT(X,Y,s,PRCC_var,y_var)

Y=Y(s,:);
[a k]=size(X); % Define the size of LHS matrix
Xranked=rankingN(X);
Yranked=ranking1(Y);
for i=1:k  % Loop for the whole submatrices, Zi
    c1=['LHStemp=Xranked;LHStemp(:,',num2str(i),')=[];Z',num2str(i),'=[ones(a,1) LHStemp];LHStemp=[];'];
    eval(c1);
end
for i=1:k
    c2=['[b',num2str(i),',bint',num2str(i),',r',num2str(i),']= regress(Yranked,Z',num2str(i),');'];
    c3=['[b',num2str(i),',bint',num2str(i),',rx',num2str(i),']= regress(Xranked(:,',num2str(i),'),Z',num2str(i),');'];
    eval(c2);
    eval(c3);
end
for i=1:k
    c4=['r',num2str(i)];
    c5=['rx',num2str(i)];
    [r p]=corr(eval(c4),eval(c5));
    a=['[PRCC , p-value] = ' '[' num2str(r) ' , '  num2str(p) '].'];% ' Time point=' num2str(s-1)];
    figure,plot((eval(c4)),(eval(c5)),'.'),Title(a),...
            legend(PRCC_var{i}),xlabel(PRCC_var{i}),ylabel(y_var);%eval(c6);

end
```
