## Supplementary material for "Cost and social distancing dynamics in a mathematical model of COVID-19 with application to Ontario, Canada": Code: ranking1.m

```
function [r,i]=ranking(x)   
% [r,i]=ranking(x)   
% Ranking of a vector  
% input:
%   x   : vector (nrow,1)  
% output:
% r : rank of the vector  (nrow,1)                                                
% i : index vector from the sort routine  (nrow,1)                                  
%                                                                               
n=length(x);
[s,i]=sort(x);                                                
r(i,1)=[1:n]';
```
