## Supplementary material for "Cost and social distancing dynamics in a mathematical model of COVID-19 with application to Ontario, Canada": Code: rankingN.m

```
function [r,i]=ranking(x)   
% [r,i]=ranking(x)   
% Ranking of a matrix  
% input:
%   x   : vector (nrow,N columns)  
% output:
% r : rank of the Matrix  (nrow,N columns)                                                
% i : index vector from the sort routine  (nrow,1)                                  
%                                                                               

%n=length(x)                                                                         
[a b]=size(x);
for j=1:b
    [s,i]=sort(x(:,j));
    r(i,j)=[1:a]';
end
```
