## Supplementary material for "Cost and social distancing dynamics in a mathematical model of COVID-19 with application to Ontario, Canada": Code: RCC_PLOT.m

```
%% Plot the ranked input (X - LHS matrix) vs the ranked output (Y) at column s (time point)
%% type: either linear-linear plot ('lin') or linear-log plot '(log')
%% RCC_var: labels of the parameters varied in the X (as legend)
%% y_var: label of the output variable
%% The Title of the plot is the Spearman correlation coefficient with the 
%% correspondent p-value and the time point chosen (s)
%% by Simeone Marino, June 5 2007 %%

function rcc_plot=RCC_PLOT(X,Y,s,type,RCC_var,y_var)
[A p]=corr(X,Y(s,:)','type','Spearman');
[a b]=size(X);
for i=1:b
    a=['[Spearman correlation , p-value] = ' '[' num2str(A(i)) ' , '  num2str(p(i)) '].'];% ' Time point=' num2str(s)]
    c=['ranking1(X(:,',num2str(i),'));'];
    %c2=['legend(var{i})'];
    if type=='log'
        figure,semilogy(eval(c),ranking1(Y(s,:)),'.'),Title(a),legend(RCC_var{i}),...
            xlabel(PRCC_var{i}),ylabel(y_var);
    elseif type=='lin'
        figure,plot(eval(c),ranking1(Y(s,:)),'.'),Title(a),legend(RCC_var{i}),...
            xlabel(RCC_var{i}),ylabel(y_var);
    end
end
```
