## Supplementary material for "Cost and social distancing dynamics in a mathematical model of COVID-19 with application to Ontario, Canada": Code: run_prcc.m

```
close all
clear variables

file_to_load = 0;
%0 - 3000 data point file on Aug 21

alpha = 0.05;

switch file_to_load
    case 0
        file_title='prcc_Model_LHS_10000_Aug2120';
        load([file_title(6:end),'.mat']);
        
end
sickS = Is+Is1+Is2+IsM+Rs+Rs1+Rs2+RsM;
sickA = Ia+Ia1+Ia2+IaM+Ra+Ra1+Ra2+RaM;
sick = sickS+sickA;
% CALCULATE PRCC for sick symptomatic
[prcc{1},sign{1},sign_label{1}]=PRCC(LHSmatrix,sickS,1:length(time_points),PRCC_var,alpha);
% CALCULATE PRCC for susceptibles
[prcc{2},sign{2},sign_label{2}]=PRCC(LHSmatrix,S+S1+S2,1:length(time_points),PRCC_var,alpha);
% CALCULATE PRCC for peak value
[prcc{3},sign{3},sign_label{3}]=PRCC(LHSmatrix,peak_val,1,PRCC_var,alpha);
% CALCULATE PRCC for peak time
[prcc{4},sign{4},sign_label{4}]=PRCC(LHSmatrix,peak_time,1,PRCC_var,alpha);
% CALCULATE PRCC for sick asymptomatic
[prcc{5},sign{5},sign_label{5}]=PRCC(LHSmatrix,sickA,1:length(time_points),PRCC_var,alpha);

% file_title=strcat(file_title,'-',strrep(strrep(datestr(clock),':','-'),' ','-'),'.mat');
save(file_title,'prcc','sign','sign_label','PRCC_var');
```
