## Supplementary material for "Cost and social distancing dynamics in a mathematical model of COVID-19 with application to Ontario, Canada": Code: new_figure.m

```
function [fig,figprops] = new_figure(title,pos)
if nargin<2
    pos = [2,4.5,6,4.5];
end

fig = figure;
set(fig,'units','inches');
set(fig,'position',pos);
set(fig,'name',title);

drawnow;

figprops.ax='';
figprops.title='';
figprops.xlab='';
figprops.ylab='';
figprops.ylab2='';
figprops.leg='';
figprops.ypos='';
figprops.ypos2='';
figprops.ax2='';
figprops.cb='';
figprops.type='figure';
figprops.fnt_size = 16;
end
```
