## Supplementary material for "Cost and social distancing dynamics in a mathematical model of COVID-19 with application to Ontario, Canada": Code: ODEf.m

```
function dy=ODEf(t,y,p,flags)

%if (t>180 && t<220) || (t>240 && t<250) || (t>260 && t<270)
% if (t>190 && t<210) || (t>237 && t<247) || (t>271 && t<284) %|| (t>280 && t<300)
if (t>180 && t<220) || (t>245 && t<255)  || (t>270 && t<280) %|| (t>280 && t<300)
% p.Kc=p.Kc/100;
p.Kc = 0;
p.K0 = 0.070122974212569;
% p.K0=150*p.Kc;

% p.Kc = 0;
% p.K0 = 0.073301177290017;
end


switch flags.model
        
    case 1
        dy = zeros(7,1);
        
        S = y(1);
        E = y(2);
        P = y(3);
        Is = y(4);
        Ia = y(5);
        Rs = y(6);
        Ra = y(7);
        
        F_S = p.alpha*p.beta*P*S+p.alpha*p.beta*Ia*S+p.beta*Is*S;
        
        dy(1) = -F_S;
        dy(2) = F_S-p.sigma*E;
        dy(3) = p.sigma*E - p.phi*P;
        dy(4) = p.q*p.phi*P - p.gamma*Is;
        dy(5) = (1-p.q)*p.phi*P - p.gamma*Ia;
        dy(6) = p.gamma*Is;
        dy(7) = p.gamma*Ia;
        
    case 2
        dy = zeros(22,1);
        
        S = y(1);
        S1 = y(2);
        S2 = y(3);
        E = y(4);
        E1 = y(5);
        E2 = y(6);
        P = y(7);
        P1 = y(8);
        P2 = y(9);
        Is = y(10);
        Is1 = y(11);
        Is2 = y(12);
        Ia = y(13);
        Ia1 = y(14);
        Ia2 = y(15);
        Rs = y(16);
        Rs1 = y(17);
        Rs2 = y(18);
        Ra = y(19);
        Ra1 = y(20);
        Ra2 = y(21);
        M = y(22);
        
        
        
        F_S = p.N0/p.N*p.beta*(p.alpha*P*S+p.alpha*Ia*S+Is*S...
            +p.alpha*p.delta*P1*S+p.alpha*p.delta*Ia1*S+p.delta*Is1*S);
        F_S1 = p.N0/p.N*p.beta*(p.alpha*p.delta*P*S1+p.alpha*p.delta*Ia*S1+p.delta*Is*S1...
            +p.alpha*p.delta^2*P1*S1+p.alpha*p.delta^2*Ia1*S1+p.delta^2*Is1*S1);
        

        mu = p.mumax*tanh(atanh(0.9)/(p.M1-p.M0)*max(M-p.M0,0));

        mu1 = 0;%0.2*mu;

        nu = -(p.numax-p.numin)*tanh(atanh(1-0.1*p.numin/(p.numax-p.numin))/(p.M1-p.M0)*max(M-p.M0,0))+p.numax;
        nu2 = nu;%1/2*nu;
        

        
        dy(1) = -F_S - mu*S + nu*S1 + (1-p.q2)*nu2*S2;
        dy(2) = -F_S1 - mu1*S1 + p.q1*mu*S - nu*S1 +p.q2*nu2*S2;
        dy(3) = (1-p.q1)*mu*S + mu1*S1 - nu2*S2;
        dy(4) = F_S - mu*E + nu*E1 + (1-p.q1)*nu2*E2 -p.sigma*E;
        dy(5) = F_S1 - mu1*E1 + p.q1*mu*E - nu*E1 + p.q1*nu2*E2 - p.sigma*E1;
        dy(6) = (1-p.q1)*mu*E + mu1*E1 - nu2*E2 - p.sigma*E2;
        dy(7) = p.sigma*E -mu*P + nu*P1 + (1-p.q1)*nu2*P2 - p.phi*P;
        dy(8) = p.sigma*E1 -mu1*P1 + p.q1*mu*P - nu*P1 + p.q1*nu2*P2 - p.phi*P1;
        dy(9) = p.sigma*E2 + (1-p.q1)*mu*P + mu1*P1 - nu2*P2 - p.phi*P2;
        dy(10) = p.q*p.phi*P - p.muI*Is - p.gamma*Is;
        dy(11) = p.q*p.phi*P1 + p.qI*p.muI*Is - p.gamma*Is1;
        dy(12) = p.q*p.phi*P2 + (1-p.qI)*p.muI*Is - p.gamma*Is2;
        dy(13) = (1-p.q)*p.phi*P - mu*Ia +nu*Ia1 + (1-p.q1)*nu2*Ia2 - p.gamma*Ia;
        dy(14) = (1-p.q)*p.phi*P1 - mu1*Ia1 + p.q1*mu*Ia - nu*Ia1 + p.q1*nu2*Ia2 - p.gamma*Ia1;
        dy(15) = (1-p.q)*p.phi*P2 + (1-p.q1)*mu*Ia + mu1*Ia1 - nu2*Ia2 - p.gamma*Ia2;
        dy(16) = p.gamma*Is;
        dy(17) = p.gamma*Is1;
        dy(18) = p.gamma*Is2;
        dy(19) = p.gamma*Ia;
        dy(20) = p.gamma*Ia1;
        dy(21) = p.gamma*Ia2;
        dy(22) = p.rho1*p.q*p.phi*(P+P1+P2) - p.muM*M;
        
    case 3
        dy = zeros(22,1);
        
        S = y(1);
        S1 = y(2);
        S2 = y(3);
        E = y(4);
        E1 = y(5);
        E2 = y(6);
        P = y(7);
        P1 = y(8);
        P2 = y(9);
        PM = y(10);
        Is = y(11);
        Is1 = y(12);
        Is2 = y(13);
        IsM = y(14);
        Ia = y(15);
        Ia1 = y(16);
        Ia2 = y(17);
        IaM = y(18);
        Rs = y(19);
        Rs1 = y(20);
        Rs2 = y(21);
        RsM = y(22);
        Ra = y(23);
        Ra1 = y(24);
        Ra2 = y(25);
        RaM = y(26);
        M = y(27);
        C = y(28);
        
        
        
        F_S = p.N0/p.N*p.beta*(p.alpha*P*S+p.alpha*Ia*S+Is*S...
            +p.alpha*p.delta*P1*S+p.alpha*p.delta*Ia1*S+p.delta*Is1*S);
        F_S1 = p.N0/p.N*p.beta*(p.alpha*p.delta*P*S1+p.alpha*p.delta*Ia*S1+p.delta*Is*S1...
            +p.alpha*p.delta^2*P1*S1+p.alpha*p.delta^2*Ia1*S1+p.delta^2*Is1*S1);
        
            KM = min(1/log(2)*max((p.rho0*(Ia+Ia1+Ia2+P+P1+P2) + p.rhoI*(Is+Is1+Is2))/M,0),1000);
            AM = PM+IsM+IaM;
%             AM = p.rho0*(P+P1+P2)+p.rhoI*(Is+Is1+Is2)+p.rho0*(Ia+Ia1+Ia2);

            mu = p.mumax*max(KM-p.Kc,0)/(max(KM-p.Kc,0)+p.K0-p.Kc)*max(AM-p.Mc,0)/(max(AM-p.Mc,0)+p.M0-p.Mc);
            mu1 = mu/2;
            
            if flags.cost_control
                nu2 = p.numax*max(C-p.Cc,0)/(max(C-p.Cc,0)+p.C0-p.Cc)*max(p.eta*p.Mc-AM,0);%/p.eta/p.Mc;
            else
                nu2 = p.numax*max(C-p.Cc,0)/(max(C-p.Cc,0)+p.C0-p.Cc);
            end
            nu = nu2/2;
            
        

        
        dy(1) = -F_S - mu*S + nu*S1 + (1-p.q2)*nu2*S2;
        dy(2) = -F_S1 - mu1*S1 + p.q1*mu*S - nu*S1 +p.q2*nu2*S2;
        dy(3) = (1-p.q1)*mu*S + mu1*S1 - nu2*S2;
        dy(4) = F_S - mu*E + nu*E1 + (1-p.q1)*nu2*E2 -p.sigma*E;
        dy(5) = F_S1 - mu1*E1 + p.q1*mu*E - nu*E1 + p.q1*nu2*E2 - p.sigma*E1;
        dy(6) = (1-p.q1)*mu*E + mu1*E1 - nu2*E2 - p.sigma*E2;
        dy(7) = p.sigma*E -mu*P + nu*P1 + (1-p.q1)*nu2*P2 - p.phi*P - p.rho0*P;
        dy(8) = p.sigma*E1 -mu1*P1 + p.q1*mu*P - nu*P1 + p.q1*nu2*P2 - p.phi*P1 - p.rho0*P1 ;
        dy(9) = p.sigma*E2 + (1-p.q1)*mu*P + mu1*P1 - nu2*P2 - p.phi*P2 - p.rho0*P2;
        dy(10) = p.rho0*(P+P1+P2) - p.phi*PM;
        dy(11) = p.q*p.phi*P - p.muI*Is - p.gamma*Is - p.rhoI*Is;
        dy(12) = p.q*p.phi*P1 + p.qI*p.muI*Is - p.gamma*Is1 - p.rhoI*Is1;
        dy(13) = p.q*p.phi*P2 + (1-p.qI)*p.muI*Is - p.gamma*Is2 - p.rhoI*Is2;
        dy(14) = p.rhoI*(Is+Is1+Is2) + p.q*p.phi*PM - p.gamma*IsM;
        dy(15) = (1-p.q)*p.phi*P - mu*Ia +nu*Ia1 + (1-p.q1)*nu2*Ia2 - p.gamma*Ia - p.rho0*Ia;
        dy(16) = (1-p.q)*p.phi*P1 - mu1*Ia1 + p.q1*mu*Ia - nu*Ia1 + p.q1*nu2*Ia2 - p.gamma*Ia1 - p.rho0*Ia1;
        dy(17) = (1-p.q)*p.phi*P2 + (1-p.q1)*mu*Ia + mu1*Ia1 - nu2*Ia2 - p.gamma*Ia2 - p.rho0*Ia2;
        dy(18) = p.rho0*(Ia+Ia1+Ia2) +(1-p.q)*p.phi*PM - p.gamma*IaM;
        dy(19) = p.gamma*Is;
        dy(20) = p.gamma*Is1;
        dy(21) = p.gamma*Is2;
        dy(22) = p.gamma*IsM;
        dy(23) = p.gamma*Ia;
        dy(24) = p.gamma*Ia1;
        dy(25) = p.gamma*Ia2;
        dy(26) = p.gamma*IsM;
        dy(27) = p.rho0*(Ia+Ia1+Ia2+P+P1+P2) + p.rhoI*(Is+Is1+Is2);
        dy(28) = p.n*((S2+E2)+(1-p.delta)*(S1+E1)) - p.muC*C;
        
        
end
```
