## Supplementary material for "Cost and social distancing dynamics in a mathematical model of COVID-19 with application to Ontario, Canada": Code: odesim.m

```
close all
clear variables


curpath = pwd;
newpath = strcat(curpath,'\runs\');
flags.save = 0; %Choose to save run or not
flags.case_name = 0; %Whether to use date stamping or case naming
title_case = 2;
flags.social_distance = 1; %Decide if social distancing is enabled (0 sets all motion rates to 0)
flags.cost_control = 0; %Decide if relaxation is controlled with cases


%SET MODEL
flags.model = 3; 
%*****************************************1 -SEPIR model
%S-susceptible, no social distancing
%E- exposed, no symptoms, not infectious
%P – Presymptomatic, infectious
%IS – infectious and symptomatic
%IA – infectious, asymptomatic
%Rs – recovered/removed from Is
%Ra – recovered/removed from Ia
%*****************************************2 -SEPIR model with social
%distancing
%S-susceptible, no social distancing
%S1-susceptible, some social distancing (3/4 reduction)
%S2-susceptible, complete social distancing (1 reduction)
%E- exposed, no symptoms, not infectious
%P – Presymptomatic, infectious
%IS – infectious and symptomatic
%IA – infectious, asymptomatic
%Rs – recovered/removed from Is
%Ra – recovered/removed from Ia
%1 and 2 subscripts for all above
%M - media reports (a proxy for testing results)
%*****************************************3 -SEPIR model with social
%distancing and cost. People who are tested are removed is positive
%S-susceptible, no social distancing
%S1-susceptible, some social distancing (3/4 reduction)
%S2-susceptible, complete social distancing (1 reduction)
%E- exposed, no symptoms, not infectious
%P – Presymptomatic, infectious
%IS – infectious and symptomatic
%IA – infectious, asymptomatic
%Rs – recovered/removed from Is
%Ra – recovered/removed from Ia
%1 and 2 subscripts for all above M for testing removal
%M - media reports (a proxy for testing results)
%C - cost of social distancing
%Note, we scale this model by N_crit instead of N

fprintf('Running model %i\n',flags.model);


%SET PARAMS
switch flags.model
    case 1
        params.R0 = 2.4;
        params.alpha = 1/2;
        params.sigma = 2;
        params.phi = 1/4.6;
        params.q = 0.35;
        params.gamma = 1/18.3;
        
        params.beta = params.R0*params.phi*params.gamma/(params.alpha*(params.gamma+params.phi)+(1-params.alpha)*params.q*params.phi);
        
        y0 = [0.999;0;0;0.001;0;0;0];
        t0 = [1,365];
        var_names = {'S';'E';'P';'I_S';'I_A';'R_S';'R_A'};
    case 2
        params.N0 = params.N;
        params.R0 = 2.4;
        
        params.alpha = 1/2;
        params.sigma = 1/4.6;
        params.phi = 2;
        params.q = 0.22;
        params.gamma = 1/10;
        
        params.beta = params.R0*params.phi*params.gamma/(params.alpha*(params.gamma+params.phi)+(1-params.alpha)*params.q*params.phi);
        params.delta = 0.25;
        

        params.numax = 24;
        params.numin = 1/7;
        params.q1 = 0.9;
        params.q2 = 0;
        params.mumax = 1;
        params.M0 = 0.1;%*N_crit/N;
        params.M1 = 0.75;%*N_crit/N;
        params.muI = 0.8*1/3;
        params.qI = 0.9;
        params.muM = 1;%1/10;%1/14;
        
        params.rho1 = 0.6*1/3;
        
        y0 = [1-3E-5;0;0;0;0;0;0;0;0;3E-5;0;0;0;0;0;0;0;0;0;0;0;0];
        y0 = y0*params.N/params.N0;
        t0 = [1,2*365];
        var_names = {'S';'S_1';'S_2';'E';'E_1';'E_2';'P';'P_1';'P_2';'I_S';'I_{S1}';'I_{S2}';'I_A';'I_{A1}';'I_{A2}';'R_S';'R_{S1}';'R_{S2}';'R_A';'R_{A1}';'R_{A2}';'M'};
        
    case 3
        run_params;
        
        if flags.social_distance==0
%             params.muI = 0;
            params.rho0 = 0;
            params.rhoI = 0;
            params.mumax = 0;
            params.numax = 0;
        end
        
       
        t0 = [1,7*365];
%         t0 = [0,161];
        var_names = {'S';'S_1';'S_2';'E';'E_1';'E_2';'P';'P_1';'P_2';'P_M';'I_S';'I_{S1}';'I_{S2}';'I_{SM}';'I_A';'I_{A1}';'I_{A2}';'I_{AM}';'R_S';'R_{S1}';'R_{S2}';'R_{SM}';'R_A';'R_{A1}';'R_{A2}';'R_{AM}';'M';'C'};
        
end

% odeopts = odeset('NonNegative',1,'RelTol',1e-8,'AbsTol',1e-9);
odeopts = odeset('NonNegative',1);
[t,y] = ode23t(@ODEf,t0,y0,odeopts,params,flags);

switch flags.model
    case 1
        sick = sum(y(:,4:7),2);
    case 2
        sick = sum(y(:,10:21),2);
        sickS = sum(y(:,10:12),2)+sum(y(:,16:18),2);
        sickA = sum(y(:,13:15),2)+sum(y(:,19:21),2);
    case 3
        sick = sum(y(:,11:26),2);
        sickS = sum(y(:,11:14),2)+sum(y(:,19:22),2);
        sickA = sum(y(:,15:18),2)+sum(y(:,23:26),2);
end

% t = t/365*12;


switch flags.model
    case 2
        for k=1:5
            figure
            hold on
            for j=1:3
                if k==1
                    plot(t,y(:,(k-1)*3+j)*params.n,'linewidth',2);
                else
                    plot(t,y(:,(k-1)*3+j),'linewidth',2);
                end
            end
            legend(var_names{(k-1)*3+1:(k-1)*3+3},'location','best');
            if k==4
                plot(t,ones(length(t),1)*params.N_crit/params.N0,'k--','linewidth',2);
                plot(t,y(:,10)+y(:,11)+y(:,12),'k','linewidth',2);
            end
            hold off
        end
        figure
        plot(t,y(:,22),'linewidth',2);
%         hold on
%         plot(t,ones(length(t),1)*params.N_crit/params.N0,'k--','linewidth',2);
%         hold off
        legend(var_names{22},'location','best');
        
        figure
        plot(t,y(:,23),'linewidth',2);
        legend(var_names{23},'location','best');
    case 3
        for k=1:2
            figure
            hold on
            for j=1:3
%                 if k==1
%                     plot(t,y(:,(k-1)*3+j)*params.n,'linewidth',2);
%                 else
                    plot(t,y(:,(k-1)*3+j),'linewidth',2);
%                 end
            end
            legend(var_names{(k-1)*3+1:(k-1)*3+3},'location','best');
            hold off
        end
        for k = 3:5
            figure
            hold on
            for j=1:4
                plot(t,y(:,6+(k-3)*4+j),'linewidth',2);
            end
            legend(var_names{6+(k-3)*4+1:6+(k-3)*4+4},'location','best');
            if k==4
                plot(t,ones(length(t),1)*params.N_crit/params.N0,'k--','linewidth',2);
                plot(t,y(:,11)+y(:,12)+y(:,13)+y(:,14),'k','linewidth',2);
            end
            hold off
        end
        figure
        yyaxis left
        plot(t,y(:,27),'linewidth',2);
        hold on
        yyaxis right
        plot(t,y(:,10)+y(:,14)+y(:,18),'linewidth',2);
        legend({var_names{27},'M_A'},'location','best');
        
        figure
        plot(t,y(:,28),'linewidth',2);
        legend(var_names{28},'location','best');
end

figure 
switch flags.model
    case 1
        plot(t,sick,'linewidth',2);
    case {2,3}
        hold on
        plot(t,sickS*params.N0/params.N,'linewidth',2);
        plot(t,sickA*params.N0/params.N,'linewidth',2);
        plot(t,sick*params.N0/params.N,'linewidth',2);
        legend({'S','A','total'},'location','best');
end


switch flags.model
    case 3
        S = y(:,1);
        S1 = y(:,2);
        S2 = y(:,3);
        E = y(:,4);
        E1 = y(:,5);
        E2 = y(:,6);
        P = y(:,7);
        P1 = y(:,8);
        P2 = y(:,9);
        PM = y(:,10);
        Is = y(:,11);
        Is1 = y(:,12);
        Is2 = y(:,13);
        IsM = y(:,14);
        Ia = y(:,15);
        Ia1 = y(:,16);
        Ia2 = y(:,17);
        IaM = y(:,18);
        M = y(:,27);
        C = y(:,28);
        
        Q = S+E+P+Is+Ia;
        Q1 = S1+E1+P1+Is1+Ia1;
        Q2 = S2+E2+P2+Is2+Ia2;
end
switch flags.model
    case 2
        mu = params.mumax*tanh(atanh(0.9)/(params.M1-params.M0)*max(M-params.M0,0));
        nu = -(params.numax-params.numin)*tanh(atanh(1-0.1*params.numin/(params.numax-params.numin))/(params.M1-params.M0)*max(M-params.M0,0))+params.numax;

        figure
        plot(t,mu,'linewidth',2);

        figure
        plot(t,nu,'linewidth',2);
    case 3
        KM = 1/log(2)*max((params.rho0*(Ia+Ia1+Ia2+P+P1+P2) + params.rhoI*(Is+Is1+Is2))./M,0);
        AM = PM + IsM + IaM;
        Kfun = max(KM-params.Kc,0)./(max(KM-params.Kc,0)+params.K0-params.Kc);
        Afun = max(AM-params.Mc,0)./(max(AM-params.Mc,0)+params.M0-params.Mc);
        mu = params.mumax*Kfun.*Afun;
        if flags.cost_control
            nu = params.numax*max(C-params.Cc,0)./(max(C-params.Cc,0)+params.C0-params.Cc).*max(params.eta*params.Mc-AM,0);%/params.eta/params.Mc;
        else
            nu = params.numax*max(C-params.Cc,0)./(max(C-params.Cc,0)+params.C0-params.Cc);
        end
        
        It = Is+Is1+Is2+IsM;
        AT = P+P1+P2+PM+It+Ia+Ia1+Ia2+IaM;
        
        figure
        plot(t,mu,'linewidth',2);
        hold on
        plot(t,nu,'linewidth',2);
        hold off
        legend('mu','nu','location','best');
        
        figure
        plot(t,Kfun,'linewidth',2);
        hold on
        plot(t,Afun,'linewidth',2);
        hold off
        legend('K','A','location','best');
        
        %Find spike times for social distancing
        donespikes = 0;
        mucount = 0;
        sgnmu = 1;
        while(~donespikes)
            sgnmu = sgnmu-1+find(sign(mu(sgnmu:end))==1,1);
            if isempty(sgnmu)
                donespikes=1;
            else
                mucount = mucount+1;
                muspots0(mucount) = sgnmu;
                tmu0(mucount) = t(sgnmu);
                sgnmu = sgnmu-1+find(sign(mu(sgnmu:end))==0,1);
                muspots1(mucount) = sgnmu;
                tmu1(mucount) = t(sgnmu);
            end
        end
        
        %Find spike times for infection
        donespikes = 0;
        Itcount = 0;
        sgnIt = 1;
        while(~donespikes)
            sgnIt = sgnIt-1+find(sign(It(sgnIt:end)-1)==1,1);
            if isempty(sgnIt)
                donespikes=1;
            else
                Itcount = Itcount+1;
                Itspots0(Itcount) = sgnIt;
                tIt0(Itcount)= t(sgnIt);
                sgnIt = sgnIt-1+find(sign(It(sgnIt:end)-1)==-1,1);
                Itspots1(Itcount) = sgnIt;
                tIt1(Itcount) = t(sgnIt);
            end
        end
        
        
        
        health_cost = 0;
        if Itcount~=0
            for k=1:length(tIt0)
                health_cost = health_cost + trapz(t(Itspots0(k):Itspots1(k)),It(Itspots0(k):Itspots1(k)));
            end
        end
        
end

if flags.save
    file_title=strcat('odemodel-',num2str(flags.model),'-q-',num2str(floor(100*params.q)));
    if flags.case_name
        file_title=strcat(file_title,'-case',num2str(title_case),'.mat');
    else
        file_title=strcat(file_title,'-',strrep(strrep(datestr(clock),':','-'),' ','-'),'.mat');
    end
    save(strcat(newpath,file_title),'t','y','params');
end

data_compare_plot
```
