## Supplementary material for "Cost and social distancing dynamics in a mathematical model of COVID-19 with application to Ontario, Canada": Code: README.txt

Parameters are set in "run_parameters".

With parameters set run "odesim" to simulate the model listed in "ODEf".

"run_cases" generates a series of case files for simulations used in experiment. These are stored in the folder "runs". The file "case_inf.xlsx" lists what each case number means in terms of parameters.

"Model_LHS" (in the LHS-PRCC folder) runs the sensitivity analysis based on setting parameters in "Parameter_settings_LHS". "run_prcc" then uses the data generated to produce the correlation coefficient results. These coefficients "prcc_Model_LHS_n_DATE" are then copied to the main folder to be used in data fitting.

The file "generate_media" produces all of the plots of the manusript reading data in "runs" and storing plots in the folder "plots"

Data used to fit the model is stored in "DATA-Aug18.mat"

Other functions not specifically mentioned are supporting functions to the above mentioned codes. Furthermore, some codes/plots are depreciated and do not appear in the manuscript

* Note we do not include the run files necessary to generate the figures. You must run_cases for each of the experiments to generate the runs needed. To do this deploy "run_cases" with loop_flag set to 1 and all of the data will be generated. You need to create a folder called "runs" in the main directory.
