## Supplementary material for "Cost and social distancing dynamics in a mathematical model of COVID-19 with application to Ontario, Canada": Code: run_cases.m

```
close all
clear variables

curpath = pwd;
newpath = strcat(curpath,'\runs\');

flags.model = 3;
flags.cost_control = 0;

loop_flag = 0;

% t0 = [1,10*365];
t0 = [1:1:10*365];
var_names = {'S';'S_1';'S_2';'E';'E_1';'E_2';'P';'P_1';'P_2';'P_M';'I_S';'I_{S1}';'I_{S2}';'I_{SM}';'I_A';'I_{A1}';'I_{A2}';'I_{AM}';'R_S';'R_{S1}';'R_{S2}';'R_{SM}';'R_A';'R_{A1}';'R_{A2}';'R_{AM}';'M';'C'};
odeopts = odeset('NonNegative',1);

exp_len = 5;

experiment = 5;
%0 is the runs for cost analysis
%1 is the runs for varying q to see peak
%2 is the runs for cost analysis with fixing q and varying testing
%3 is the runs for cost analysis with modified cost formula
%4 is the same as 0 but with other Mc values
%5 vary Mc to see social distancing effects

if loop_flag
    experiment = 0;
end
done = 0;
while ~done
    run_params;
    Mc = params.Mc;
    Cc = params.Cc;
    rho0 = params.rho0;
    rhoI = 4*params.rho0;
    eta = params.eta;
    if ~loop_flag
        done = 1;
    end
    fprintf('Running experiment %i...\n',experiment);
    switch experiment
        case 0

            q = [0.9;0.69;0.27];

            multiplier = [1/4;1/2;1;2;4];

            tic;

            for K = 0:1
                flags.social_distance = K;
                if flags.social_distance
                    params.rho0 = rho0;
                    params.rhoI = rhoI;
                    params.mumax = 1;
                    params.numax = params.mumax;
                else
%                     params.rho0 = 0;
%                     params.rhoI = 0;
                    params.mumax = 0;
                    params.numax = 0;
                end
                for k = 1:length(q)
                    params.q = q(k);
                    params.beta = params.R0*params.phi*params.gamma/(params.alpha*(params.gamma+params.phi)+(1-params.alpha)*params.q*params.phi);
                    if flags.social_distance
                        for j1 = 1:length(multiplier)
                            params.Mc = multiplier(j1)*Mc;
                            params.M0 = 2*params.Mc;
                            for j2=1:length(multiplier)
                                title_case = (j1-1)*length(multiplier)+j2;
                                fprintf('Running case %i q=%i...\n',title_case,floor(100*params.q));
                                params.Cc = multiplier(j2)*Cc;
                                params.C0 = 2*params.Cc;
                                [t,y] = ode23t(@ODEf,t0,y0,odeopts,params,flags);
                                file_title=strcat('odemodel-',num2str(flags.model),'-q-',num2str(floor(100*params.q)),'-case',num2str(title_case),'.mat');
                                save(strcat(newpath,file_title),'t','y','params');
                            end
                        end
                    else
                        fprintf('Running case 0 q=%i...\n',floor(100*params.q));
                        params.Mc = Mc;
                        params.M0 = 2*params.Kc;
                        params.Cc = Cc;
                        params.C0 = 2*params.Cc;
                        [t,y] = ode23t(@ODEf,t0,y0,odeopts,params,flags);
                        file_title=strcat('odemodel-',num2str(flags.model),'-q-',num2str(floor(100*params.q)),'-case0.mat');
                        save(strcat(newpath,file_title),'t','y','params');
                    end
                end
            end
            end_time=toc;

            fprintf('Simulations completed, running time was %3.2f...\n',end_time);
        case 1
            tic;
            q = linspace(0,1,100);
%             params.rho0 = 0;
%             params.rhoI = 0;
            params.mumax = 0;
            params.numax = 0;
            for k = 1:length(q)
                params.q=q(k);
                params.beta = params.R0*params.phi*params.gamma/(params.alpha*(params.gamma+params.phi)+(1-params.alpha)*params.q*params.phi);
                fprintf('Running case %i...\n',k);
                [t,y] = ode23t(@ODEf,t0,y0,odeopts,params,flags);
                file_title=strcat('qstudy-odemodel-',num2str(flags.model),'-q-',num2str(floor(100*params.q)),'.mat');
                save(strcat(newpath,file_title),'t','y','params');
            end
            end_time=toc;
            fprintf('Simulations completed, running time was %3.2f...\n',end_time);
        case 2

            rho_multiplier = [1/2;1;2];
            rho_label = {'0p5','1','2'};
            multiplier = [1/4;1/2;1;2;4];
            params.beta = params.R0*params.phi*params.gamma/(params.alpha*(params.gamma+params.phi)+(1-params.alpha)*params.q*params.phi);

            tic;

%             params.rho0 = 0;
%             params.rhoI = 0;
            params.mumax = 0;
            params.numax = 0;

            fprintf('Running base case...\n');
            [t,y] = ode23t(@ODEf,t0,y0,odeopts,params,flags);
            file_title=strcat('odemodel-',num2str(flags.model),'-rho_baseline','.mat');
            save(strcat(newpath,file_title),'t','y','params');


            params.mumax = 1;
            params.numax = params.mumax;

            for k = 1:length(rho_multiplier)
                params.rho0 = rho_multiplier(k)*rho0;
                params.rhoI = rho_multiplier(k)*rhoI;
                for j1 = 1:length(multiplier)
                    params.Mc = multiplier(j1)*Mc;
                    params.M0 = 2*params.Kc;
                    for j2=1:length(multiplier)
                        title_case = (j1-1)*length(multiplier)+j2;
                        fprintf('Running case %i rho_mult=%i...\n',title_case,k);
                        params.Cc = multiplier(j2)*Cc;
                        params.C0 = 2*params.Cc;
                        [t,y] = ode23t(@ODEf,t0,y0,odeopts,params,flags);
                        file_title=strcat('odemodel-',num2str(flags.model),'-rho_mult-',rho_label{k},'-case',num2str(title_case),'.mat');
                        save(strcat(newpath,file_title),'t','y','params');
                    end
                end
            end
            end_time=toc;

            fprintf('Simulations completed, running time was %3.2f...\n',end_time);
        case 3
            multiplier = [1/4;1/2;1;2;4];
            flags.cost_control = 1;

            tic;

            for K = 0:1
                flags.social_distance = K;
                if flags.social_distance
                    params.rho0 = rho0;
                    params.rhoI = rhoI;
                    params.mumax = 1;
                    params.numax = params.mumax;
                else
%                     params.rho0 = 0;
%                     params.rhoI = 0;
                    params.mumax = 0;
                    params.numax = 0;
                end
                
                if flags.social_distance
                    for j1 = 1:length(multiplier)
                       params.eta = multiplier(j1)*eta;
                        for j2=1:length(multiplier)
                            title_case = (j1-1)*length(multiplier)+j2;
                            fprintf('Running case %i q=%i...\n',title_case,floor(100*params.q));
                            params.Cc = multiplier(j2)*Cc;
                            params.C0 = 2*params.Cc;
                            [t,y] = ode23t(@ODEf,t0,y0,odeopts,params,flags);
                            file_title=strcat('odemodel-',num2str(flags.model),'-eta-case',num2str(title_case),'.mat');
                            save(strcat(newpath,file_title),'t','y','params');
                        end
                    end
                else
                    fprintf('Running case 0 q=%i...\n',floor(100*params.q));
                    params.eta = eta;
                    params.Cc = Cc;
                    params.C0 = 2*params.Cc;
                    [t,y] = ode23t(@ODEf,t0,y0,odeopts,params,flags);
                    file_title=strcat('odemodel-',num2str(flags.model),'-eta-case0.mat');
                    save(strcat(newpath,file_title),'t','y','params');
                end
            end
            end_time=toc;

            fprintf('Simulations completed, running time was %3.2f...\n',end_time);
        case 4
            q = [0.9;0.69;0.27];

            multiplier = [8;16;32;64;128];
            cmultiplier = [1/4;1/2;1;2;4];

            tic;

            for K = 0:1
                flags.social_distance = K;
                if flags.social_distance
                    params.rho0 = rho0;
                    params.rhoI = rhoI;
                    params.mumax = 1;
                    params.numax = params.mumax;
                else
%                     params.rho0 = 0;
%                     params.rhoI = 0;
                    params.mumax = 0;
                    params.numax = 0;
                end
                for k = 1:length(q)
                    params.q = q(k);
                    params.beta = params.R0*params.phi*params.gamma/(params.alpha*(params.gamma+params.phi)+(1-params.alpha)*params.q*params.phi);
                    if flags.social_distance
                        for j1 = 1:length(multiplier)
                            params.Mc = multiplier(j1)*Mc;
                            params.M0 = 2*params.Mc;
                            for j2=1:length(multiplier)
                                title_case = (j1-1)*length(multiplier)+j2;
                                fprintf('Running case %i q=%i...\n',title_case,floor(100*params.q));
                                params.Cc = cmultiplier(j2)*Cc;
                                params.C0 = 2*params.Cc;
                                [t,y] = ode23t(@ODEf,t0,y0,odeopts,params,flags);
                                file_title=strcat('moreMc-odemodel-',num2str(flags.model),'-q-',num2str(floor(100*params.q)),'-case',num2str(title_case),'.mat');
                                save(strcat(newpath,file_title),'t','y','params');
                            end
                        end
                    else
                        fprintf('Running case 0 q=%i...\n',floor(100*params.q));
                        params.Mc = Mc;
                        params.M0 = 2*params.Kc;
                        params.Cc = Cc;
                        params.C0 = 2*params.Cc;
                        [t,y] = ode23t(@ODEf,t0,y0,odeopts,params,flags);
                        file_title=strcat('MoreMc-odemodel-',num2str(flags.model),'-q-',num2str(floor(100*params.q)),'-case0.mat');
                        save(strcat(newpath,file_title),'t','y','params');
                    end
                end
            end
            end_time=toc;

            fprintf('Simulations completed, running time was %3.2f...\n',end_time);
        case 5
            params.q = 0.69;
            params.Cc = Cc;
            params.C0 = 2*params.Cc;
            
            Mcvals = linspace(0,0.02,5000)*params.N/params.N_crit;

            tic;
            
            params.mumax = 0;
            params.numax = 0;
            fprintf('Running case 0\n');
            params.Mc = Mc;
            params.M0 = 2*params.Kc;
            [t,y] = ode23t(@ODEf,t0,y0,odeopts,params,flags);
            file_title=strcat('testMc-odemodel-',num2str(flags.model),'-case0.mat');
            save(strcat(newpath,file_title),'t','y','params');
            
            params.rho0 = rho0;
            params.rhoI = rhoI;
            params.mumax = 1;
            params.numax = params.mumax;          
            params.beta = params.R0*params.phi*params.gamma/(params.alpha*(params.gamma+params.phi)+(1-params.alpha)*params.q*params.phi);
            for k=1:length(Mcvals)
                params.Mc = Mcvals(k);
                params.M0 = 2*params.Mc;
                if mod(k,100)==0
                    fprintf('Running case %i \n',k);
                end
                [t,y] = ode23t(@ODEf,t0,y0,odeopts,params,flags);
                file_title=strcat('testMc-odemodel-',num2str(flags.model),'-case',num2str(k),'.mat');
                save(strcat(newpath,file_title),'t','y','params');
            end
            end_time=toc;

            fprintf('Simulations completed, running time was %3.2f...\n',end_time);
    end
    experiment = experiment +1;
    if experiment > exp_len
        done = 1;
    end
end
```
