## Supplementary material for "Cost and social distancing dynamics in a mathematical model of COVID-19 with application to Ontario, Canada": Code: run_params.m

```
params.N = 13448494; %population
params.N_crit = 10000/0.123; %critical hospital capacity 12.3% hospitalization from ontario website

params.N0=params.N_crit;
params.n = params.N0/params.N;
params.R0 = 2.4;

params.alpha = 1/2;
params.sigma = 2;%1/4.6;
params.phi = 1/4.6;%2;
params.gamma = 1/10;%1/9.14

params.q = 0.69; %0.9,0.69,0.27


params.beta = params.R0*params.phi*params.gamma/(params.alpha*(params.gamma+params.phi)+(1-params.alpha)*params.q*params.phi);
params.delta = 0.25;


params.q1 = 0.9;
params.q2 = 0.6;

params.mumax = 1;
params.numax = params.mumax;


params.muI = 0.01*params.mumax;
params.qI = 0.6;

params.rho0 = 8.7E-3;%6E-4;%0.03*1/100;
params.rhoI = 4*params.rho0;%13E-4;%0.12*1/100;


params.Mc = 2090/params.N0; %100/params.N0;
params.M0 = 2*params.Mc;
params.Kc = 1/16.24;%1/100;
params.K0 = 4*params.Kc;
params.Cc = 50;
params.C0 = 2*params.Cc;
params.muC = 0;

params.eta = 1;

infected_initial = 2E-4*params.N;%3E-3*params.N;

y0 = [params.N-infected_initial;0;0;0;0;0;0;0;0;0;infected_initial;0;0;0;0;0;0;0;0;0;0;0;0;0;0;0;0;0];
y0 = y0/params.N0;
```
