## Supplementary material for "Cost and social distancing dynamics in a mathematical model of COVID-19 with application to Ontario, Canada": Code: shadedErrorBar.m

```
function varargout=shadedErrorBar(x,y,errBar,varargin)
% generate continuous error bar area around a line plot
%
% function H=shadedErrorBar(x,y,errBar, ...)
%
% Purpose 
% Makes a 2-d line plot with a pretty shaded error bar made
% using patch. Error bar color is chosen automatically.
%
%
% Inputs (required)
% x - vector of x values [optional, can be left empty]
% y - vector of y values or a matrix of n observations by m cases
%     where m has length(x);
% errBar - if a vector we draw symmetric errorbars. If it has a size
%          of [2,length(x)] then we draw asymmetric error bars with
%          row 1 being the upper bar and row 2 being the lower bar
%          (with respect to y -- see demo). ** alternatively ** 
%          errBar can be a cellArray of two function handles. The 
%          first defines statistic the line should be and the second 
%          defines the error bar.
%
% Inputs (optional, param/value pairs)
% 'lineProps' - ['-k' by default] defines the properties of
%             the data line. e.g.:    
%             'or-', or {'-or','markerfacecolor',[1,0.2,0.2]}
% 'transparent' - [true  by default] if true, the shaded error
%               bar is made transparent. However, for a transparent
%               vector image you will need to save as PDF, not EPS,
%               and set the figure renderer to "painters". An EPS 
%               will only be transparent if you set the renderer 
%               to OpenGL, however this makes a raster image.
% 'patchSaturation'- [0.2 by default] The saturation of the patch color.
%
%
%
% Outputs
% H - a structure of handles to the generated plot objects.
%
%
% Examples:
% y=randn(30,80); 
% x=1:size(y,2);
%
% 1)
% shadedErrorBar(x,mean(y,1),std(y),'lineProps','g');
%
% 2)
% shadedErrorBar(x,y,{@median,@std},'lineProps',{'r-o','markerfacecolor','r'});
%
% 3)
% shadedErrorBar([],y,{@median,@(x) std(x)*1.96},'lineProps',{'r-o','markerfacecolor','k'});
%
% 4)
% Overlay two transparent lines:
% clf
% y=randn(30,80)*10; 
% x=(1:size(y,2))-40;
% shadedErrorBar(x,y,{@mean,@std},'lineProps','-r','transparent',1);
% hold on
% y=ones(30,1)*x; y=y+0.06*y.^2+randn(size(y))*10;
% shadedErrorBar(x,y,{@mean,@std},'lineProps','-b','transparent',1);
% hold off
%
%
% Rob Campbell - November 2009
%%%%%%%%%%%%%%%%%%%%%%%%%%%%%%%%%%%%%%%%%%%%%%%%%%%%%%%%%%%%%
% Parse input arguments
narginchk(3,inf)
params = inputParser;
params.CaseSensitive = false;
params.addParameter('lineProps', '-k', @(x) ischar(x) | iscell(x));
if (sum( size(ver('MATLAB'))) > 0  )
  params.addParameter('transparent', true, @(x) islogical(x) || x==0 || x==1);
elseif (sum( size(ver('Octave'))) > 0  )
  params.addParameter('transparent', false, @(x) islogical(x) || x==0 || x==1);
end
params.addParameter('patchSaturation', 0.2, @(x) isnumeric(x) && x>=0 && x<=1);
params.parse(varargin{:});
%Extract values from the inputParser
lineProps =  params.Results.lineProps;
transparent =  params.Results.transparent;
patchSaturation = params.Results.patchSaturation;
if ~iscell(lineProps), lineProps={lineProps}; end
%Process y using function handles if needed to make the error bar dynamically
if iscell(errBar) 
    fun1=errBar{1};
    fun2=errBar{2};
    errBar=fun2(y);
    y=fun1(y);
else
    y=y(:).';
end
if isempty(x)
    x=1:length(y);
elseif sum( size(ver('MATLAB'))) > 0 
    x=x(:).';
end
%Make upper and lower error bars if only one was specified
if length(errBar)==length(errBar(:))
    errBar=repmat(errBar(:)',2,1);
else
    s=size(errBar);
    f=find(s==2);
    if isempty(f), error('errBar has the wrong size'), end
    if f==2, errBar=errBar'; end
end
% Check for correct x, errbar formats
x_size = size(x);
if (length(x) ~= length(errBar) && sum( size(ver('MATLAB'))) > 0 )
    error('length(x) must equal length(errBar)')
elseif( ( length(x) ~= length(errBar) && checkOctave_datestr(x) == false ) ...
            && sum( size(ver('Octave'))) > 0  )
    error('length(x) must equal length(errBar) or x must have valid datestr')
end
 
%Log the hold status so we don't change
initialHoldStatus=ishold;
if ~initialHoldStatus, hold on,  end
H = makePlot(x,y,errBar,lineProps,transparent,patchSaturation);
if ~initialHoldStatus, hold off, end
if nargout==1
    varargout{1}=H;
end
function H = makePlot(x,y,errBar,lineProps,transparent,patchSaturation)
    %%%%%%%%%%%%%%%%%%%%%%%%%%%%%%%%%%%%%%%%%%%%%%%%%%%%%%%%%%%%%
    % Determine host application
    if (sum( size(ver('MATLAB'))) > 0  )
      hostName = 'MATLAB';
    elseif (sum(size(ver('Octave'))) > 0)
      hostName = 'Octave';
    end % if
    
    % Plot to get the parameters of the line
    if hostName == 'MATLAB'
      H.mainLine=plot(x,y,lineProps{:});
      
    elseif hostName == 'Octave'
      boolxDatestr = checkOctave_datestr(x);
      if boolxDatestr
        x = datenum(x);
        x = x(:).';
        H.mainLine=plot(x,y,lineProps{:});
        datetick(gca);
      else
        H.mainLine=plot(x,y,lineProps{:});
      end
    end
    % Tag the line so we can easily access it
    H.mainLine.Tag = 'shadedErrorBar_mainLine';
    % Work out the color of the shaded region and associated lines.
    % Here we have the option of choosing alpha or a de-saturated
    % solid colour for the patch surface.
    mainLineColor=get(H.mainLine,'color');
    edgeColor=mainLineColor+(1-mainLineColor)*0.55;
    if transparent
        faceAlpha=patchSaturation;
        patchColor=mainLineColor;
    else
        faceAlpha=1;
        patchColor=mainLineColor+(1-mainLineColor)*(1-patchSaturation);
    end
    %Calculate the error bars
    uE=y+errBar(1,:);
    lE=y-errBar(2,:);
    %Make the patch (the shaded error bar)
    yP=[lE,fliplr(uE)];
    xP=[x,fliplr(x)];
    %remove nans otherwise patch won't work
    xP(isnan(yP))=[];
    yP(isnan(yP))=[];
    
    if isdatetime(x) && strcmp(hostName,'MATLAB')
      H.patch=patch(datenum(xP),yP,1);
    else
      H.patch=patch(xP,yP,1);
    end
    set(H.patch,'facecolor',patchColor, ...
        'edgecolor','none', ...
        'facealpha',faceAlpha, ...
        'HandleVisibility', 'off', ...
        'Tag', 'shadedErrorBar_patch')
    %Make pretty edges around the patch. 
    H.edge(1)=plot(x,lE,'-');
    H.edge(2)=plot(x,uE,'-');
    set([H.edge], 'color',edgeColor, ...
      'HandleVisibility','off', ...
      'Tag', 'shadedErrorBar_edge')
    % Ensure the main line of the plot is above the other plot elements
    if hostName == 'MATLAB'
      if strcmp(get(gca,'YAxisLocation'),'left') %Because re-ordering plot elements with yy plot is a disaster
        uistack(H.mainLine,'top')
      end
    elseif hostName == 'Octave'
      % create the struct from scratch by temp.
      H = struct('mainLine', H.mainLine, ...
      'patch', H.patch, ...
      'edge', H.edge);
    end
function boolDate = checkOctave_datestr(x)
  %% Simple try/catch for casting datenums, requireing valid datestr
  boolDate = true;
  try
    datenum(x)
  catch
    boolDate = false;
  end
```
